# Evaluation of statistical methods used in the analysis of interrupted time series studies: a simulation study

**DOI:** 10.1101/2020.10.12.20211706

**Authors:** Simon L Turner, Andrew B Forbes, Amalia Karahalios, Monica Taljaard, Joanne E McKenzie

**Author notes:** Corresponding Author: Joanne McKenzie, mail: Level 4, 553 St. Kilda Road, Melbourne, 3004, Australia.

## Abstract

Interrupted time series (ITS) studies are frequently used to evaluate the effects of population-level interventions or exposures. To our knowledge, no studies have compared the performance of different statistical methods for this design. We simulated data to compare the performance of a set of statistical methods under a range of scenarios which included different level and slope changes, varying lengths of series and magnitudes of autocorrelation. We also examined the performance of the Durbin-Watson (DW) test for detecting autocorrelation. All methods yielded unbiased estimates of the level and slope changes over all scenarios. The magnitude of autocorrelation was underestimated by all methods, however, restricted maximum likelihood (REML) yielded the least biased estimates. Underestimation of autocorrelation led to standard errors that were too small and coverage less than the nominal 95%. All methods performed better with longer time series, except for ordinary least squares (OLS) in the presence of autocorrelation and Newey-West for high values of autocorrelation. The DW test for the presence of autocorrelation performed poorly except for long series and large autocorrelation. From the methods evaluated, OLS was the preferred method in series with fewer than 12 points, while in longer series, REML was preferred. The DW test should not be relied upon to detect autocorrelation, except when the series is long. Care is needed when interpreting results from all methods, given confidence intervals will generally be too narrow. Further research is required to develop better performing methods for ITS, especially for short series.

## 1 Introduction

Interrupted time series (ITS) studies are frequently used to evaluate the impact of interventions or exposures that occur at a particular point in time^1-4^. Although randomised trials are the gold standard study design, randomisation may be infeasible or undesirable in the case of policy evaluation or interventions that are implemented at a population level. Randomization also is not an option for retrospective evaluation of interventions or exposures such as natural disasters or pandemics. The use of an ITS design may be considered in these situations, as they are one of the strongest non-randomised experimental designs^2 5 6^.

In an ITS study, observations are collected at regular time points before and after an interruption, and often analysed in aggregate using a summary statistic (e.g. mean, proportion) within a time interval (e.g. weekly, monthly, or annually). A key feature of the design is that data from the pre-interruption interval can be used to estimate the underlying secular trend. When this trend is modelled correctly, it can be projected into the post-interruption interval, providing a counterfactual for what would have occurred in the absence of the interruption. From this counterfactual, a range of effect measures can be constructed that characterise the impact of the interruption. Two commonly used measures include the ‘change in level’ – which represents the change immediately after the interruption, and the ‘change in slope’ – which represents the difference in trends before and after the interruption.

A key feature of time series data is that there is the potential for non-independence of consecutive data points (serial autocorrelation)^7^. In the presence of positive autocorrelation, statistical methods that do not account for this correlation will give spuriously small standard errors (SEs)^8^. Several statistical methods are available to account for autocorrelation, such as Prais-Winsten generalised least squares or the Newey-West correction to SEs, or to directly model the error, such as autoregressive integrated moving averages (ARIMA). Further, several methods are available for testing for the presence of autocorrelation, with the Durbin-Watson test being the most commonly used^4 6^. While the performance of some of these methods has been examined for time series data^9 10^, their performance in the context of ITS studies has received relatively little attention.

In this study, we therefore aimed to examine the performance of a range of statistical methods for analysing uncontrolled ITS studies using segmented linear models. We restrict our evaluation to ITS designs where there is a single interruption, with an equal number of time points pre and post interruption, and with first order autoregressive errors. The structure of the paper is as follows: In Section 2, we begin by introducing a motivating example for this research. In Section 3, we describe the statistical model and estimation methods used in our simulation study. In Sections 4 and 5, we present the methods and results from the statistical simulation study. In Section 6, we return to our motivating example and demonstrate the impact of applying the methods outlined in Section 3. Finally, in Section 7 we present key findings and implications for practice.

## 2 Motivating example

Healthcare-associated infections (HAIs) are a common complication affecting patients in hospitals. *Clostridium difficile (C difficile)* infection is an example of one such infection that can cause serious gastrointestinal disease. As such, many countries require mandatory surveillance of *C difficile* infection rates in hospitals. When outbreaks of *C difficile* occur, the cleaning and disinfection regimes in hospitals are often changed in an attempt to reduce the infection rate. The routine collection of data in this context has led to many retrospective investigations of the effects of different interventions (e.g. novel disinfectants) to reduce *C difficile* infection using ITS data^11^. Hacek et al^12^ provides an example of such a study, where they examined the effect of terminal room cleaning with dilute bleach (Figure 1) on the rate of patients (per 1000 patient days) with a positive test for *C difficile*. Data were aggregated at monthly intervals. The series was relatively short – a scenario which is not atypical of these studies – with 10 data points pre and 24 post the intervention^11^. In the context of HAIs, there is a tendency for consecutive data points to be more similar to each other, manifesting as ‘clusters’ of data points in time (Figure 1). Fitting a segmented linear regression model to the data shows an apparent immediate decrease in the infection rate (level change), as well as a decrease in the trend (slope change). In the following section, we outline different statistical methods to estimate the model parameters and return to this example in Section 6, where we apply these methods and compare the results.

**Figure 1:**
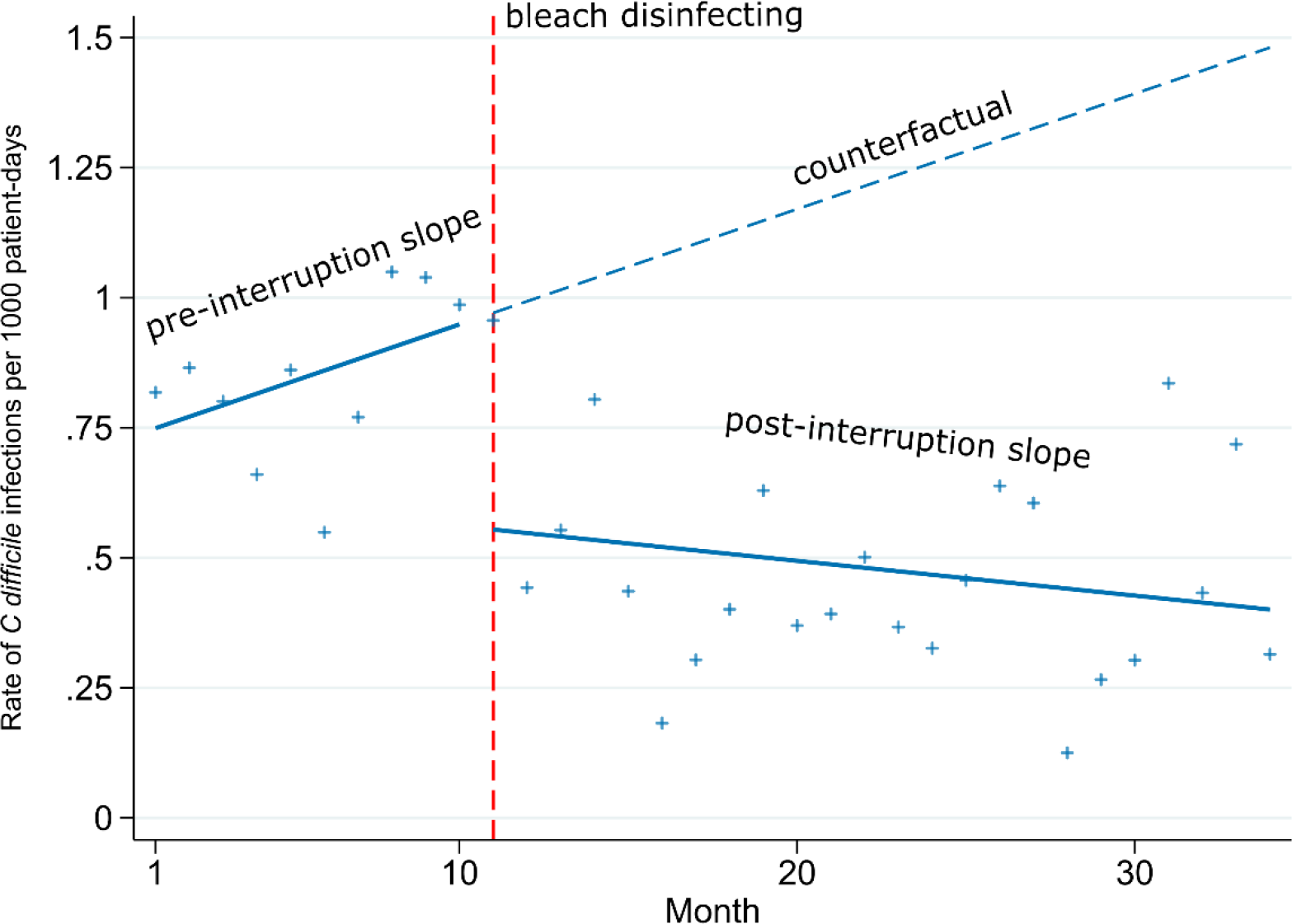
Rate of Clostridium difficile infections (per 1000 patient-days) pre and post bleach disinfection intervention per month.

## 3 Interrupted time series (ITS): model and estimation methods

We begin by describing the statistical model and parameters used in our simulation study followed by a brief description of some common statistical estimation methods and the Durbin-Watson test for autocorrelation.

### 3.1 Statistical model

We use a segmented linear regression model with a single interruption, which can be written using the parameterisation proposed by Huitema and McKean^13^ as:

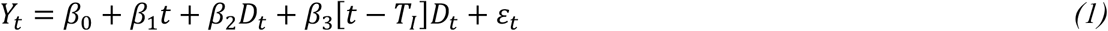

where *Y*_*t*_ represents the outcome at time point *t* of *N* time points. *D*_*t*_ is an indicator variable that represents the post-interruption interval (i.e. *D*_*t*=_1 (*t* ≥*T*_*I*_) where *T*_*I*_ represents the time of the interruption). The model parameters, *β*_0_, *β*_1_, *β*_2_ and *β*_3_ represent the intercept (e.g. baseline rate), slope in the pre-interruption interval, the change in level and the change in slope, respectively. The error term, *ε*_t_, represents deviations from the fitted model, which are constructed as:

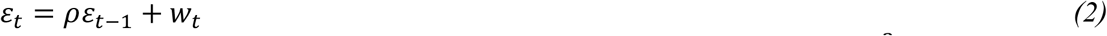

where *w*_*t*_ represents “white noise” that is normally distributed (_*t*_*∼N*(0,*σ*^2^), and *ρ* is the lag-1 autocorrelation of the errors which can range from -1 to +1. A lag-1 error means that the influence of errors on the current error is restricted to the value immediately prior. Longer lags are possible but in this paper we confine attention to lag-1 only (AR(1) errors).

### 3.2 Estimation methods

A range of statistical estimation methods are available for estimating the model parameters. These methods account for autocorrelation in different ways and are briefly described below. We focus on statistical methods that have been more commonly used (Ordinary Least Square (OLS), Generalised Least Squares (GLS), Newey-West (NW), Autoregressive Integrated Moving Average (ARIMA))^2-4 6^. In addition, we have included Restricted Maximum Likelihood (REML) (with and without the Satterthwaite adjustment), which although is not a method in common use, is included because of its potential for reduced bias in the estimation of the autocorrelation parameter, as has been discussed for general (non-interrupted) time series^14^. Further details and equations can be found in Appendix 1.

#### 3.2.1 Ordinary Least Squares

Estimates of the regression parameters and their variances from model (1) can be obtained from fitting a segmented linear regression model using OLS (Appendix 1.1). In the presence of autocorrelation, the OLS estimators for the regression parameters are unbiased; however, the SEs will be incorrect^15^.

#### 3.2.2 Newey-West

The NW estimator of the variance of the regression parameters estimated using OLS accommodates autocorrelation and heteroskedasticity of the error terms in the regression model (1)^16^ (Appendix 1.2).

#### 3.2.3 Generalised least squares

Two common GLS methods for estimating the regression parameters and their variances are Cochrane-Orcutt (CO) and Prais-Winsten (PW). For both methods, a regression model is first fitted using OLS and an estimate of the autocorrelation is calculated from the residuals. This estimate is then used to transform the data and remove the autocorrelation from the errors, upon which the regression parameters are then estimated from the transformed data. If there is still some residual autocorrelation these steps are iterated until a criterion is met (e.g., the estimated value for autocorrelation has converged^17^). The CO method applies the transformation from the second observation onwards (t=2, 3, … n). The PW method is a modification to the CO method in which a transformed value is used for the first observation (Appendix 1.3). The PW method is therefore likely to be more efficient in small series since it does not discard the first observation. The sampling properties of the estimators of the regression parameters are likely to be adversely affected when the series length is small due to poor estimation of the autocorrelation.

#### 3.2.4 Restricted maximum likelihood

It is well known that maximum likelihood estimators of variance components are biased in small samples due to not accounting for the degrees of freedom (d.f.) used when estimating the fixed effect regression parameters^18^. Restricted maximum likelihood is a variant of maximum likelihood estimation and attempts to address the bias by separating the log-likelihood into two terms; one that involves the mean and variance parameters, and one which is only dependent on the variance parameters. By maximising the latter term first with the appropriate number of d.f., an estimate of the variance parameter can be obtained which can be used when maximising the former, thus correctly accounting for the d.f.^14 19^.

For small samples, there is greater uncertainty in the estimation of the SE of the regression parameters. To account for this uncertainty in making inferences about the regression parameters, the Satterthwaite adjustment can be used to adjust the t-distribution d.f. used in hypothesis testing and calculation of confidence limits^20^.

#### 3.2.5 Autoregressive integrated moving average

In an ARIMA model, information from past values, including lagged values of the dependent variable and errors, are explicitly modelled. This is achieved by including regression coefficients for these variables in the ARIMA model. The lagged values can be from a range of previous time points, extending beyond lag-1 models. By explicitly modelling the influence of data from previous time points, their impact at subsequent times is quantified and estimates of the magnitude of autocorrelation can be obtained along with regression parameter estimates. Here we consider ARIMA models with only a first order autoregressive term (an ARIMA(1,0,0) model) estimated via maximum likelihood. ARIMA models have been shown to not perform well with fewer than fifty points^21^. Further details about the method can be found in Appendix 1.4, Nelson^21^ and Box et al^22^.

### 3.3 Durbin-Watson test for autocorrelation

The Durbin-Watson (DW) test is commonly used for detecting autocorrelation in time series. Often, the test is used as part of a two-stage analysis strategy to determine whether to use a method that adjusts for autocorrelation or use OLS (which does not adjust for autocorrelation). The null hypothesis is that there is no autocorrelation (*H*_0_:*ρ*=0) against the alternative that autocorrelation is present (*H*_1_:*ρ*≠0). The DW-statistic can range between zero and four, with values close to two indicating no autocorrelation. The DW-statistic is compared to critical values to determine whether there is evidence of autocorrelation, no autocorrelation, or the test is inconclusive. The critical values differ by series length, significance level and the d.f. in the regression model. Further details are available in Appendix 1.5, Kutner et al^15^ and Durbin and Watson^23^.

## 4 Simulation study methods

We undertook a numerical simulation study, examining the performance of a set of statistical methods under a range of scenarios which included different level and slope changes, varying lengths of series and magnitudes of autocorrelation. Design parameter values were combined using a fully factorial design with 10,000 data sets generated per combination. A range of criteria were used to evaluate the performance of the statistical methods. We now describe the methods of the simulation study using the ADEMP (defining aims, data-generating mechanisms, estimands, methods and performance measures) structure^24^.

### 4.1 Data Generating Mechanisms

We simulated data from ITS studies by randomly sampling from a parametric model (equation 1), with a single interruption at the midpoint, and first order autoregressive errors (examples shown in Supplementary 1.1). We multiplied the first error term, *ε*, by 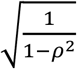 so that the variance of the error term was constant at all time points.

We created a range of simulation scenarios including different values of the model parameters and different numbers of data points per series (Table 1). These values were informed by our review of ITS studies^4^, where we reanalysed available data sets to estimate level and slope changes (standardised by the residual standard deviation), and autocorrelation. We found a median standardised level change of 1.5 (inter-quartile range (IQR): 0.6 to 3.0), n=190), median standardised slope change of 0.13 (IQR: 0.06 to 0.27, n=190) and median autocorrelation 0.2 (IQR: 0 to 0.6, n=180). We therefore constructed models with level changes (*β*_2_) of 0, 0.5, 1 and 2, and slope changes (*β*_3_) of 0 and 0.1. We did not examine negative level or slope changes since we did not expect this to influence the performance metrics. Autocorrelation was varied between 0 and 0.8 in increments of 0.2 to cover the full range of autocorrelations observed in the ITS studies included in the review. The number of data points per series was varied from 6 to 100, equally divided before and after the interruption, informed by the number of data points observed in the ITS studies (median 48, IQR: 30 to 100, n=230). The increment size was varied; initially it was small (i.e. 2) so as to detect changes in the performance metrics that were expected to arise with smaller sample sizes and was increased to 4 and then 20.

**Table 1:** Simulation parameter

| Parameter | Symbol | Parameter Values |
| --- | --- | --- |
| Intercept | $\beta_0$ | 0 |
| Pre-interruption slope | $\beta_1$ | 0 |
| Level change | $\beta_2$ | 0, 0.5, 1, 2 |
| Change in slope post-interruption | $\beta_3$ | 0, 0.1 |
| Autocorrelation coefficient | $\rho$ | 0, 0.2, 0.4, 0.6, 0.8 |
| Variance of white noise error component | $\sigma^2$ | 1 |
| Number of data points |  | 6, 8, 10, 12, 14, 16, 18, 20<br>24, 28, 32, 36, 40, 44, 48, 52, 56<br>60, 80, 100 |

All combinations of the factors in Table 1 were simulated, leading to 800 different simulation scenarios (Table 1, Figure 2).

**Figure 2:**
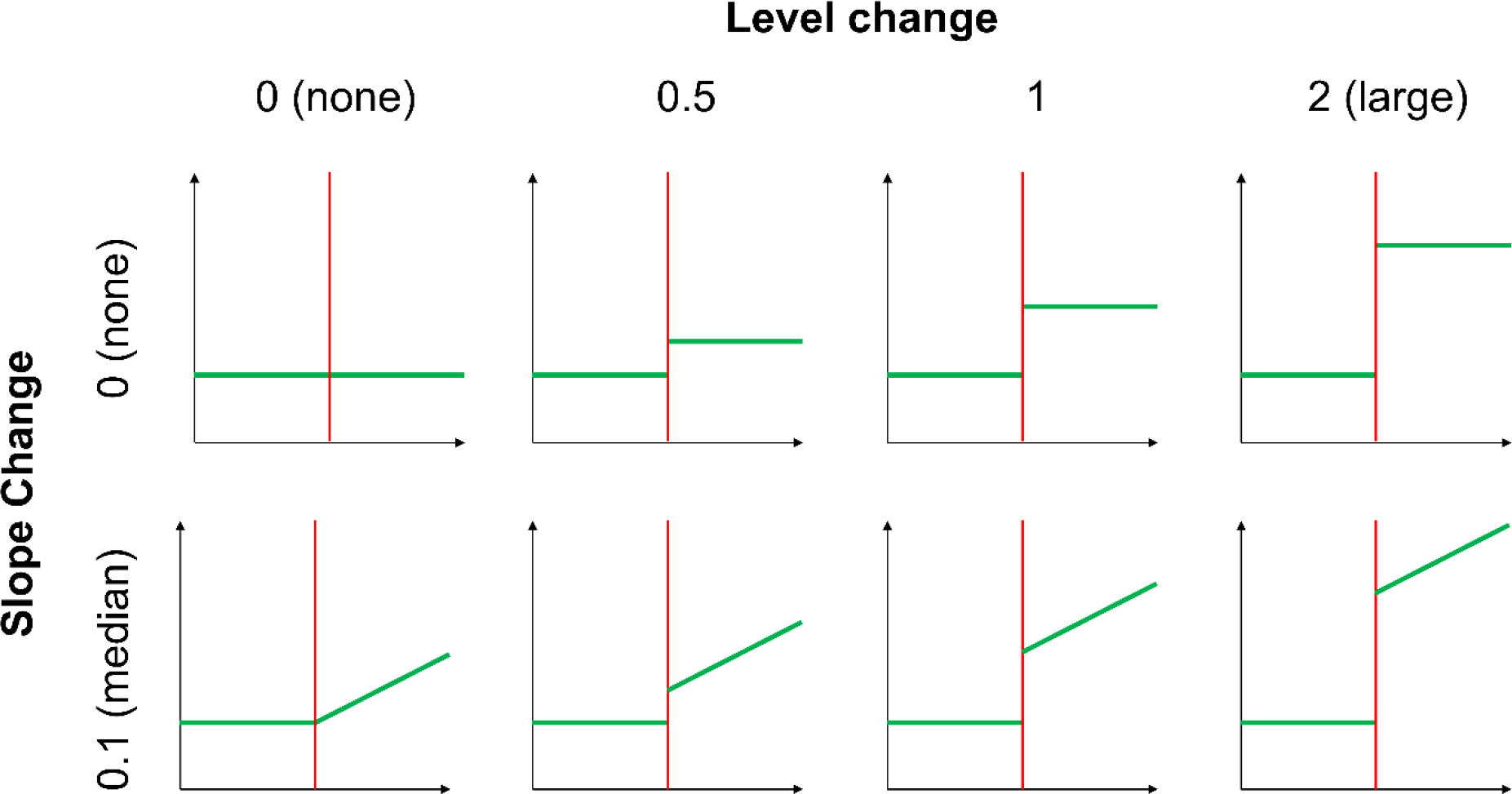
Structure of the eight models constructed from different combinations of the model input parameters (Table 1).

### 4.2 Estimands and other targets

The primary estimands of the simulation study are the parameters of the model, *β*_2_ (level change) and *β*_3_ (slope change) (Equation 1). These were chosen as they are commonly reported effect measures^4^. We also examined the autocorrelation coefficient, *ρ*, and the value of the Durbin Watson statistic.

### 4.3 Statistical Methods to analyse ITS studies

Segmented linear regression models were fitted using the estimation methods described in Section 2.2. We evaluated estimation methods designed to estimate the model parameters under lag-1 autocorrelation (see Table 2 for details). For GLS, we restricted our investigation to the PW method, because it was expected to have better performance than the CO method (on which PW is based) given the PW method utilises all data points. For REML with the Satterthwaite adjustment, we substituted d.f. of 2 when the computed d.f. were less than 2, to avoid overly conservative confidence limits and hypothesis tests. We also investigated the commonly used Durbin-Watson method for detecting autocorrelation at a significance level of 0.05^23^.

**Table 2:**
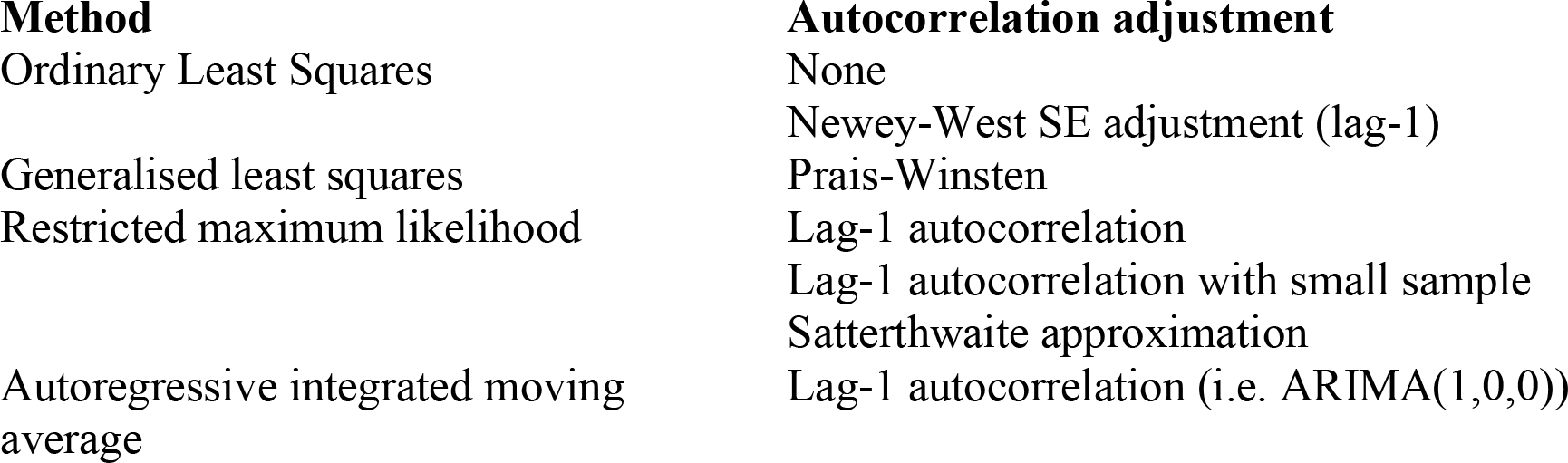
Statistical methods and adjustments for autocorrelation.

Table 2 summarises the methods and model variations used to adjust for autocorrelation. Details of the Stata code used for generating the simulated data and the analysis code can be found in Data Files 1-7.

### 4.4 Performance Measures

The performance of the methods was evaluated by examining bias, empirical SE, model-based SE, 95% confidence interval coverage and power (see Appendix 2 for formulae). Confidence intervals were calculated using the simsum package^25^ with t-distribution critical values. For each simulation scenario, we used 10,000 repetitions in order to keep the Monte Carlo Standard Error (MCSE) below 0.5% for all potential values of coverage and type I error rate. Model non-convergence was recorded and tabulated.

### 4.5 Coding and Execution

The statistical software Stata version 15^26^ was used for the generation of the simulated data. A random seed was set at the beginning of the process and the individual random state was recorded for each repetition of the simulated data sets. Each dataset was independently simulated, using consecutive randomly generated numbers from the starting seed. We used a “burn in” period between each dataset of 300 random number generations so that any lag effects specific to the computer-generated series had time to dissipate^8^.

Prior to running the simulations, we undertook initial checks to confirm that the data generation mechanism was working as expected. This involved fitting series of length 100,000 to check the estimated *β* parameters matched the input parameters. A larger sample of 1,000 datasets was then simulated and checked using summary statistics and graphs. When we were satisfied that the simulations were operating as expected, the full number of datasets were simulated.

### 4.6 Analysis of the simulated datasets

Analyses were performed using Stata version 15^26^. A range of visual displays were constructed to compare the performance of the statistical methods. Frequency distributions were plotted to visualise the level- and slope-change estimates, autocorrelation coefficient estimates, and the results of the Durbin-Watson test for autocorrelation. Scatter plots were used to display the mean values for empirical and model-based SEs, coverage, power and autocorrelation coefficient estimates. Line plots were used to show confidence intervals for the level and slope change estimates. Results and summaries of the analyses were summarised (using the simsum package^25^) and graphed using Stata version 15^26^.

## 5 Results of the simulation study

### 5.1 Bias of level and slope change estimates

All methods yielded approximately unbiased estimates of level change and slope change across all simulation scenarios. Figure 3 presents level change estimates specific to the scenario of a level change of 2 and a slope change of 0.1 (Supplementary Figure S2 shows slope change estimates), but the other 7 combinations of level and slope changes were virtually identical (Supplementary 1.3.1 for level change, Supplementary 1.3.2 for slope change). Note that the Satterthwaite and NW adjustments do not impact the parameter estimates of level or slope change, hence distributions of these parameter estimates are not shown in Figures 3 and S2.

**Figure 3:**
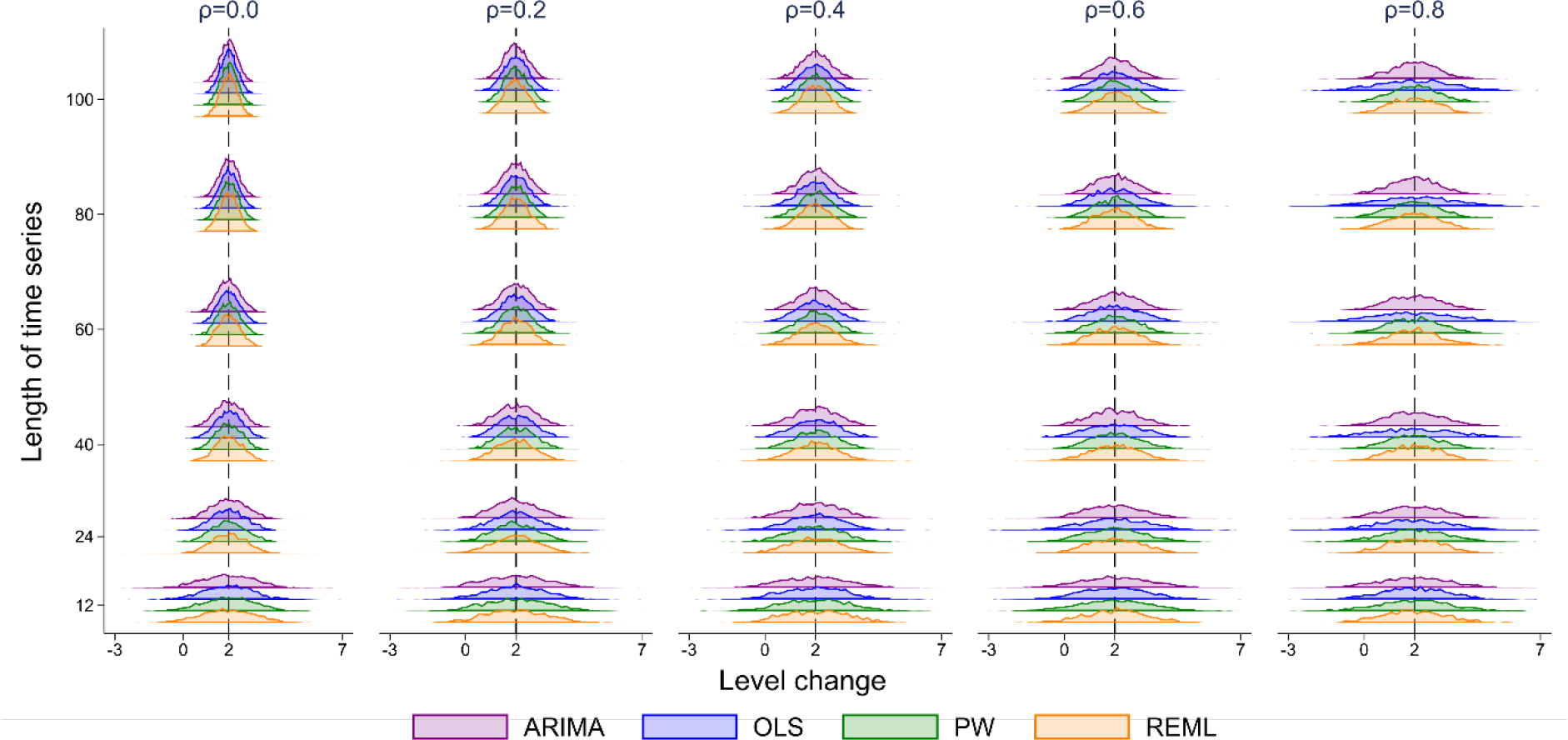
Distributions of level change estimates calculated from four statistical methods, from top to bottom: autoregressive integrated moving average (ARIMA) (purple), ordinary least squares regression (OLS) (blue), Prais-Winsten (PW) (green) and restricted maximum likelihood (REML) (orange). The vertical axis shows the length of the time series. The five vertical columns display the results for different values of autocorrelation. The vertical black line represents the true parameter value (β2). Each subset of four curves shows the distribution from a different analysis method for a given combination of time series length and autocorrelation. The simulation combination presented is for a level change of 2 and slope change of 0.1; however, other structures give similar results. The Satterthwaite adjustment to the REML method and the Newey-West adjustment to the OLS method do not impact the estimate of level or slope change, hence these parameter estimates are not shown.

### 5.2 Standard errors of level and slope change estimates

#### 5.2.1 Empirical standard errors

Figure 3 and Supplementary Figure S2 visually indicate the precision of the estimators in terms of the spread of the distributions therein. To enable a direct quantitative assessment, we plotted the empirical SE of the level and slope changes for each method against selected series lengths and autocorrelation parameter sizes for a level change of 2 and slope change of 0.1 (Figure 4 and Figure 5). The size of the empirical SE of the level change was dependent on the underlying autocorrelation, length of the series and statistical method (Figure 4). Of note, the estimates obtained from the ARIMA and PW models yield almost identical empirical SEs. For each magnitude of autocorrelation, the empirical SE decreased as the length of the time series increased, as would be expected. An exception to this occurred for the OLS estimator (and to a lesser extent ARIMA) which exhibited unusual behaviour for an autocorrelation of 0.8, with the SE initially increasing with an increasing number of points in the series, and then decreasing. Additional simulations were undertaken to examine the behaviour of the OLS estimator for surrounding correlations (0.7 and 0.9), which showed a similar pattern of increasing SEs with an increasing number of points (Supplementary 1.4). The relationship between autocorrelation and the empirical SE was modified by the length of series. For small series (< 10 data points), the empirical SE decreased with increasing autocorrelation, while for longer series (≥ 10 data points) this relationship was reversed, with SEs increasing with increasing autocorrelation.

**Figure 4:**
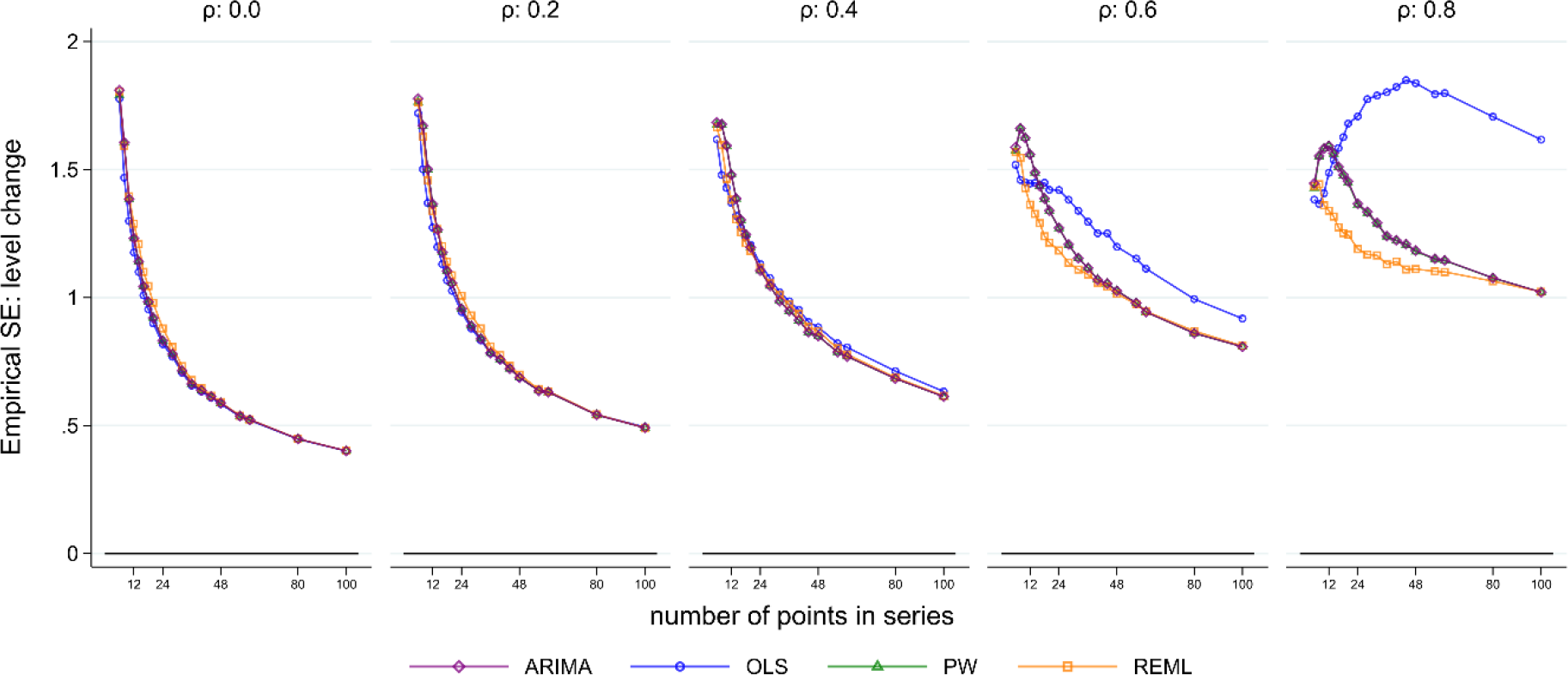
Empirical standard error (SE) of the level change. The horizontal axis shows the length of the time series, the vertical axis shows the empirical SE. The five vertical columns display the results for different values of autocorrelation. The simulation combination presented is for a level change of 2 and slope change of 0.1; however, other combinations give similar results. Abbreviations: ARIMA, autoregressive integrated moving average; OLS, ordinary least squares; PW, Prais-Winsten; REML, restricted maximum likelihood.

**Figure 5:**
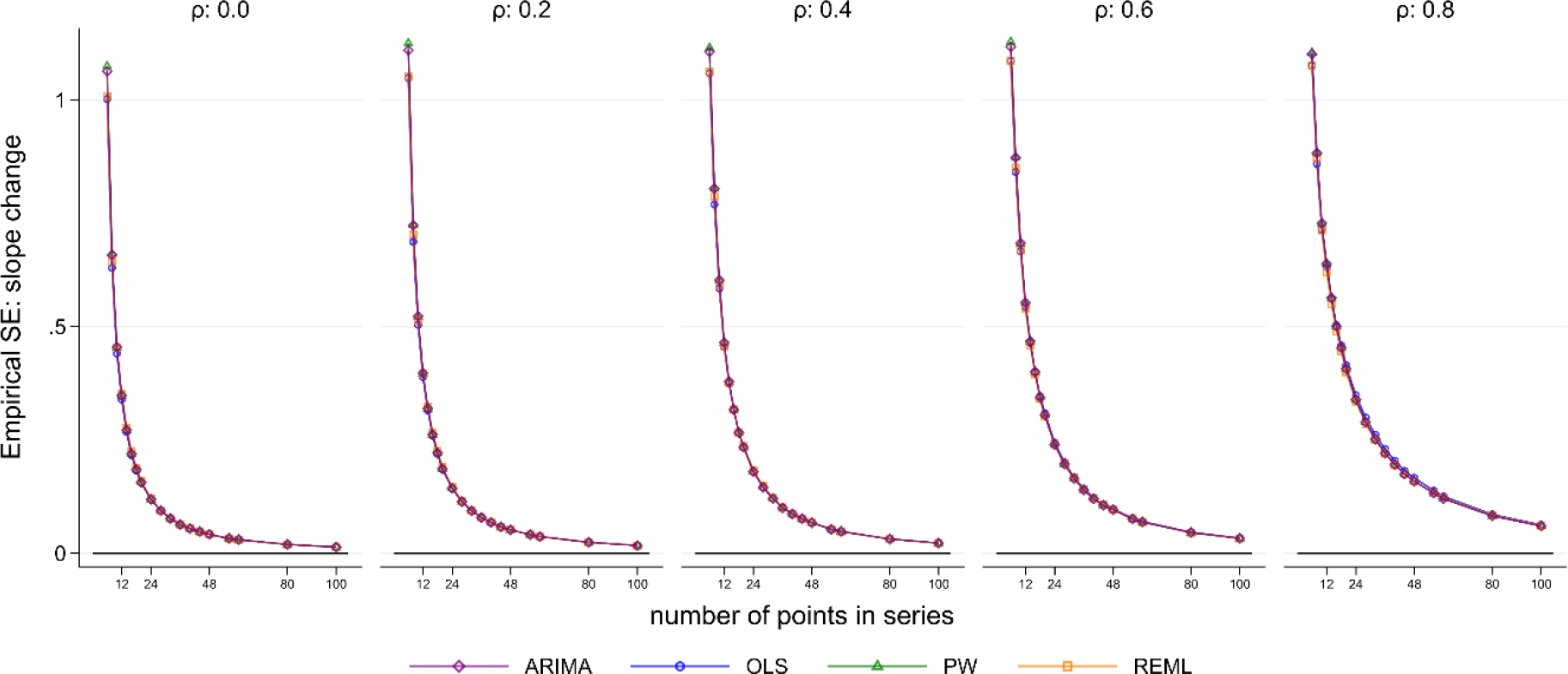
Empirical standard error (SE) of the slope change. The horizontal axis shows the length of the time series, the vertical axis shows the empirical SE. The five vertical columns display the results for different values of autocorrelation. The simulation combination presented is for a level change of 2 and slope change of 0.1; however, other combinations give similar results. Abbreviations: ARIMA, autoregressive integrated moving average; OLS, ordinary least squares; PW, Prais-Winsten; REML, restricted maximum likelihood.

The size of the empirical SE for slope change was dependent on the underlying autocorrelation and length of the series (Supplementary Figure S2 and Figure 5). The empirical SE decreased with increasing series length, but increased with increasing autocorrelation, as would be expected. In contrast to the level change, there were no important differences in the empirical SEs across the statistical methods, even when the autocorrelation was large. The observed patterns did not differ for any of the eight level and slope change combinations (Supplementary 1.3.3 for level change, Supplementary 1.3.4 for slope change).

#### 5.2.2 Comparison between empirical and model-based standard errors

To enable appropriate confidence interval coverage and size of significance tests, the model-based SE needs to be similar to the empirical SE^24^. In this section we present the comparison between the empirical and model-based SEs; results for the model-based SEs alone can be found in Supplementary 1.5.

For the level change parameter (*β*_2_) estimated by OLS, the ratio of model-based to empirical SEs were close to one (indicating the empirical and model-based SEs were similar) for all series lengths when there was no underlying autocorrelation (Figure 6). However, as autocorrelation increased, as expected, the OLS model-based SEs became increasingly smaller relative to the empirical SEs, indicating the model-based SEs is are downwardly biased. The NW method performed only slightly better than the OLS (except when the autocorrelation was zero); however, the NW model-based SEs were still downwardly biased across all scenarios, were worse than OLS for small series lengths, and only marginally better than OLS for large series lengths. Although the empirical SEs of the ARIMA and PW methods were similar, they had quite different model-based SEs. The PW model-based SEs were smaller than the empirical SEs for all magnitudes of autocorrelation, though the model-based SEs approached the empirical SEs with increasing series length. The ARIMA model-based SEs were larger than the empirical SEs for small series (fewer than 24 points) at small underlying values of autocorrelation (*ρ* < 0.4) and also for larger series (more than 24 points) at higher magnitudes of autocorrelation (*ρ* > 0.4). Aside from these scenarios, the ARIMA model-based SEs were approximately equal to the empirical SEs. The REML method behaved similarly to the PW method but, relatively, did not underestimate the SEs to the same extent. For small values of underlying autocorrelation (*ρ* < 0.4) and series greater than 30 points, the model-based SEs were similar to the empirical SEs.

**Figure 6:**
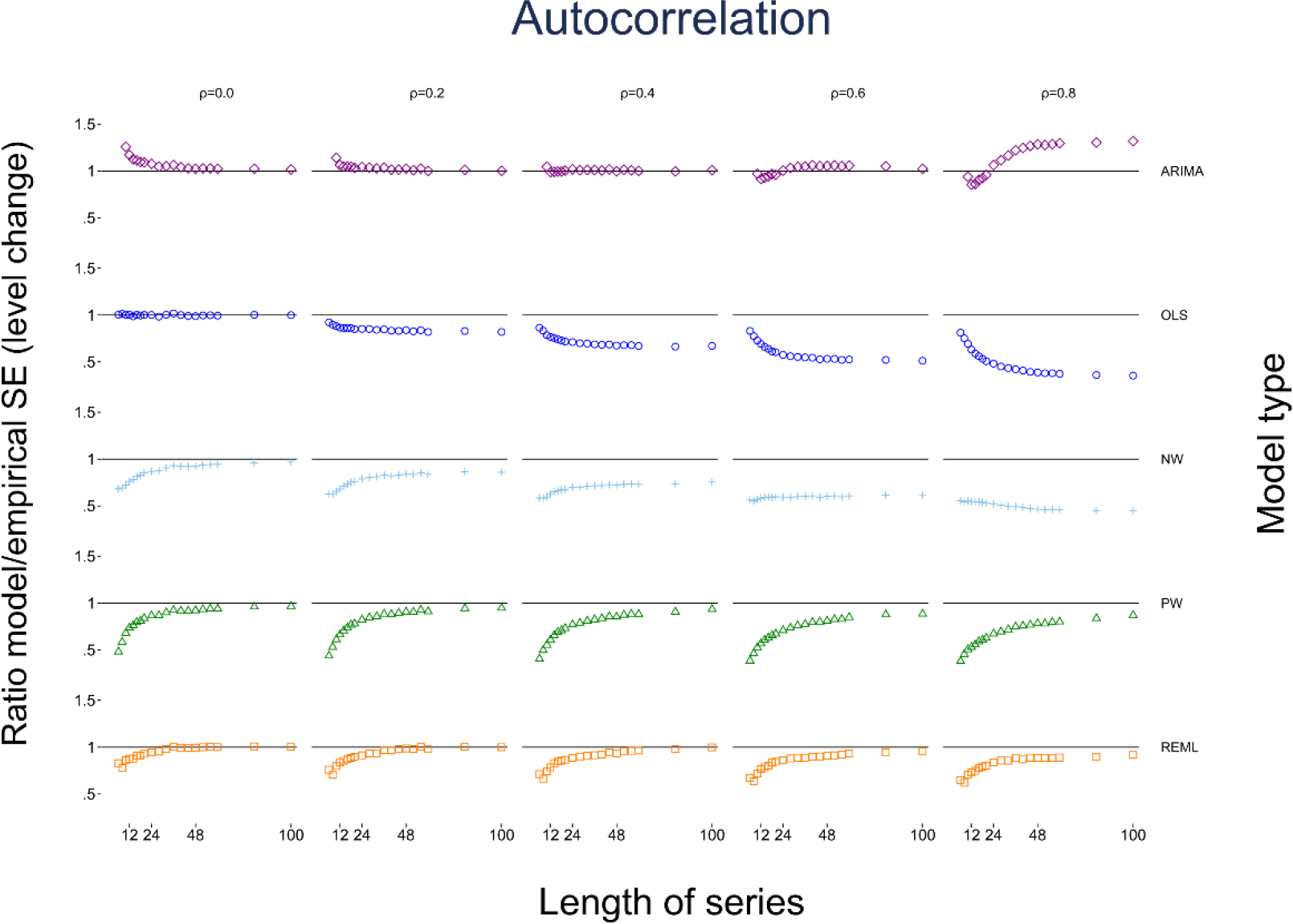
Scatter plots of the ratio of model-based standard error (SE) to the empirical SE for the level change parameter with different levels of autocorrelation and series length. The horizontal axis represents the number of points in the time series, the vertical axis shows the ratio of model-based to empirical SE. The five vertical columns display the results for different values of autocorrelation. The simulation combination presented is for a level change of 2 and slope change of 0.1; however, other combinations give similar results. The first two series lengths are not shown for the ARIMA method due to extreme values. The Satterthwaite adjustment to the REML does not impact the estimate of SE, hence details of this method are not shown. Abbreviations: ARIMA, autoregressive integrated moving average; OLS, ordinary least squares; NW, Newey-West; PW, Prais-Winsten; REML, restricted maximum likelihood.

For the slope change parameter (*β*_3_), the ratios of model-based to empirical SEs followed similar patterns as for the level change parameter (*β*_2_). For any given series length, as the magnitude of autocorrelation increased, model-based SEs became increasingly smaller compared with the empirical SEs for most statistical methods (Supplementary 1.6). Model-based and empirical SEs tended towards equivalence as series lengths increased, with the exception of OLS and NW at high values of autocorrelation (*ρ* > 0.6). For REML and ARIMA, the pattern of ratios of model-based to empirical SEs for *β*_3_ slightly differed compared with *β*_2_. Specifically, the REML model-based SEs were smaller than the empirical SEs for small series, and then increased to be slightly larger as the number of points increased. For ARIMA, the model-based SEs were smaller than the empirical SEs for large underlying values of autocorrelation (*ρ* ≥ 0.6) for small to moderate length series. The observed patterns did not differ for any of the eight level and slope change combinations (Supplementary 1.3.5 for level change, Supplementary 1.3.6 for slope change).

### 5.3 Confidence interval coverage

For all combinations of level change, slope change, number of time points and autocorrelation, most methods had coverage (percentage of 95% confidence intervals including the true parameter) that was less than the nominal 95% level for both level and slope change (Figure 7 for level change and Supplementary 1.7 for slope change, both with a level change of 2 and slope change of 0.1, Supplementary 1.3.7 for level change and Supplementary 1.3.8 for slope change for other parameter combinations). The exceptions were OLS when there was no underlying autocorrelation, and REML with the Satterthwaite adjustment for moderate to large length series. In general, mean values of coverage decreased with increasing autocorrelation and increased with increasing series length. However, coverage of the OLS method decreased with increasing autocorrelation as well as with increasing series length (with the exception of the zero autocorrelation scenario). The NW method exhibited a similar pattern to OLS, but generally had better coverage (except for small autocorrelations), although coverage was often poor (under 90% for all but the longest series with low autocorrelation, *ρ* < 0.4). REML with the Satterthwaite small sample adjustment yielded coverage greater than the nominal 95% level when the number of data points was greater than 30 in the presence of autocorrelation. Confidence interval coverage patterns generally reflected those observed with the comparisons between the model-based and empirical SE.

**Figure 7:**
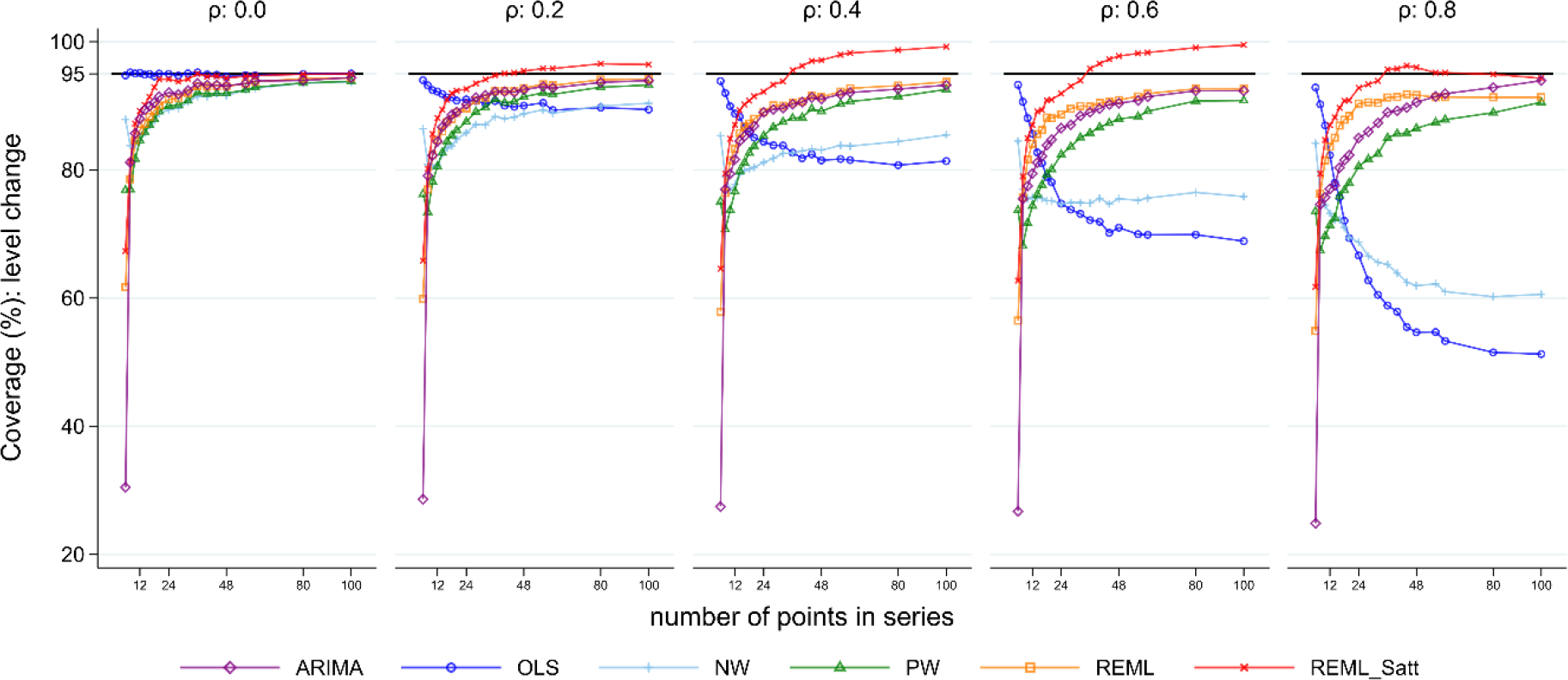
Coverage for the level change parameter. Each point is the proportion of the 10,000 simulations in which the 95% confidence interval included the true value of the parameter. The solid black line depicts the nominal 95% coverage level. The simulation combination presented is for a level change of 2 and slope change of 0.1; however, other combinations give similar results. Abbreviations: ARIMA, autoregressive integrated moving average; OLS, ordinary least squares; NW, Newey-West; PW, Prais-Winsten; REML, restricted maximum likelihood; Satt, Satterthwaite.

### 5.4 Power

Coverage was less than the nominal 95% level in the majority of scenarios (except for the OLS model in the absence of autocorrelation and some scenarios involving the REML method with Satterthwaite adjustment). In scenarios where coverage is less than 95%, examining power is misleading. Due to there being only a very small number of configurations in Figure 7 and Supplementary 1.7 in which 95% coverage was achieved, we adopt a more liberal approach and consider configurations in which the coverage was at least 90%. As such, the results presented below should be viewed as approximate power only and will generally be lower than the value observed if coverage was at least 95%.

For scenarios with a level change of two, power was low for series with a small number of points, but predictably, increased as the number of points increased for all methods, except the REML method with Satterthwaite adjustment (Figure 8). As the magnitude of autocorrelation increased its power decreased, to a point where it became lower than for other methods. This was due to the REML method with Satterthwaite adjustment having greater than 95% coverage in these situations and hence substantially lower than 5% Type I error rates. For smaller values of the level change parameter, predictably, power decreased (Supplementary 1.8.1). Similar patterns were observed for slope change (Supplementary 1.8.2).

**Figure 8:**
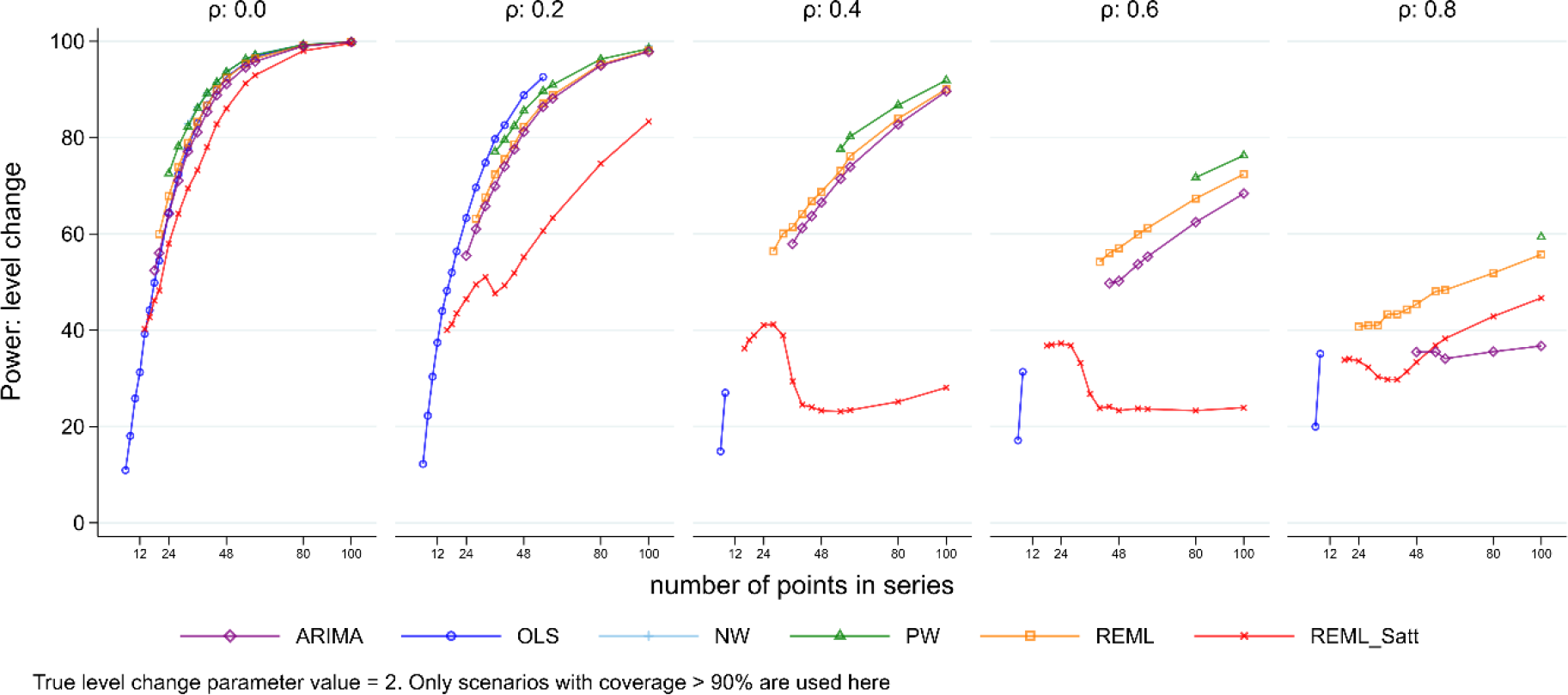
Power for level change. Each point is the mean number of times the 95% confidence interval of the estimate did not include zero from 10,000 simulations. The simulation combination presented is for a level change of 2 and slope change of 0.1. Power for other model combinations is available in Supplementary 1.8.1. Abbreviations: ARIMA, autoregressive integrated moving average; OLS, ordinary least squares; NW, Newey-West; PW, Prais-Winsten; REML, restricted maximum likelihood; Satt, Satterthwaite; NW, Newey-West.

### 5.5 Autocorrelation Coefficient

Most of the statistical methods yield an estimate of the autocorrelation coefficient. All methods underestimated the autocorrelation for series with a small number of points (Figure 9 and Figure 10 show parameter values of 2 for level change and 0.1 for slope change). However, underestimation was most pronounced for scenarios with small series and large underlying autocorrelation. The REML method always yielded estimated autocorrelations closer to the true underlying autocorrelation compared with the other methods. The empirical SEs for autocorrelation generally decreased as the series length increased for all methods (except for small series with fewer than 20 points) (Supplementary 1.9). The observed patterns did not differ for any of the eight level and slope change combinations (Supplementary 1.3.9).

**Figure 9:**
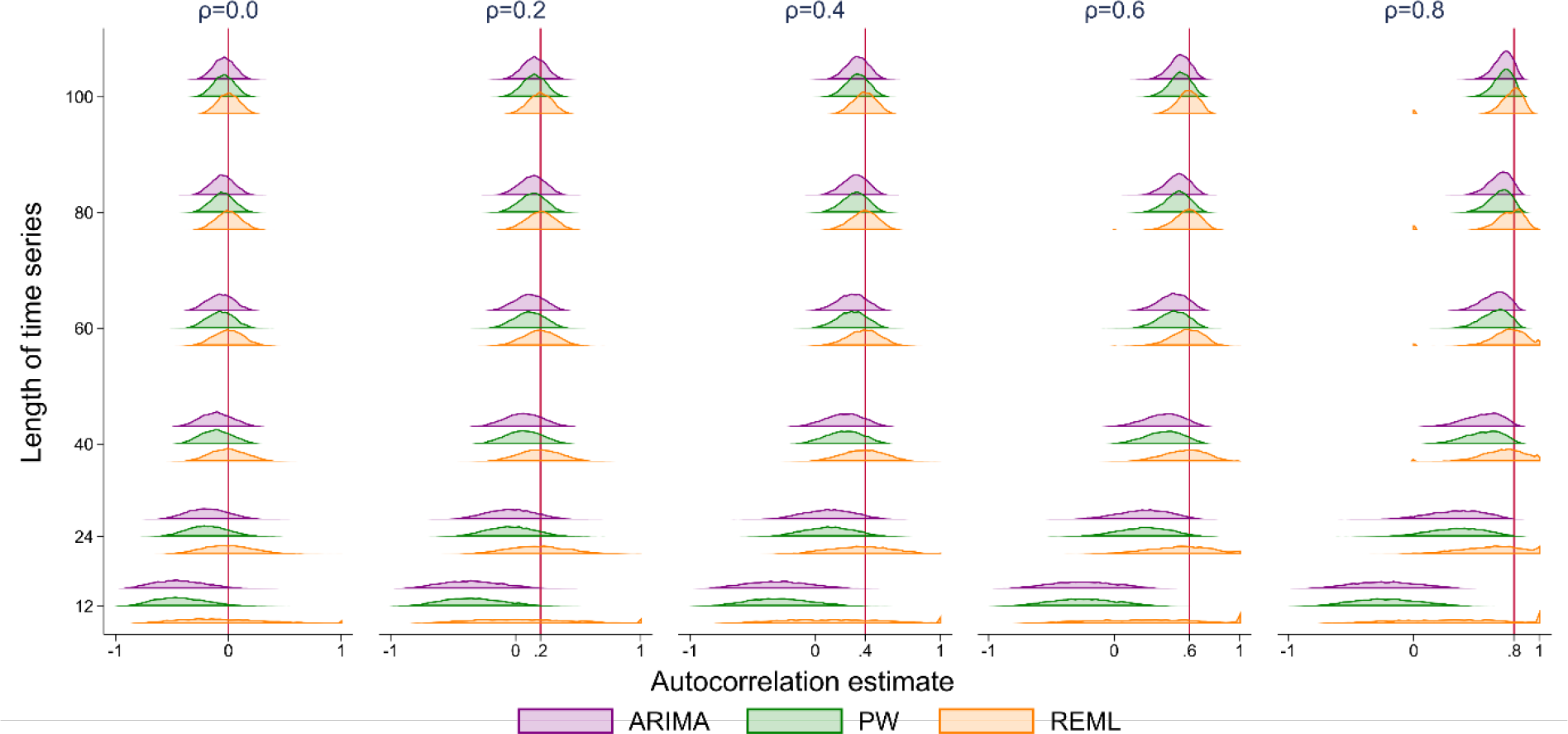
Autocorrelation coefficient estimates. The horizontal axis shows the estimate of autocorrelation coefficient. The vertical axis shows the length of the time series. The five vertical columns display the results for different values of autocorrelation ranging from 0 to 0.8 (the value of autocorrelation is shown by a vertical red line). Each coloured curve shows the distribution of autocorrelation coefficient estimates from 10,000 simulations. Each subset of four curves shows the results from a different analysis method for a given combination of time series length and autocorrelation. The simulation combination presented is for a level change of 2 and slope change of 0.1; however, other combinations give similar results. From top to bottom the methods are autoregressive integrated moving average (ARIMA) (purple), Prais-Winsten (PW) (green) and restricted maximum likelihood (REML) (orange).

**Figure 10:**
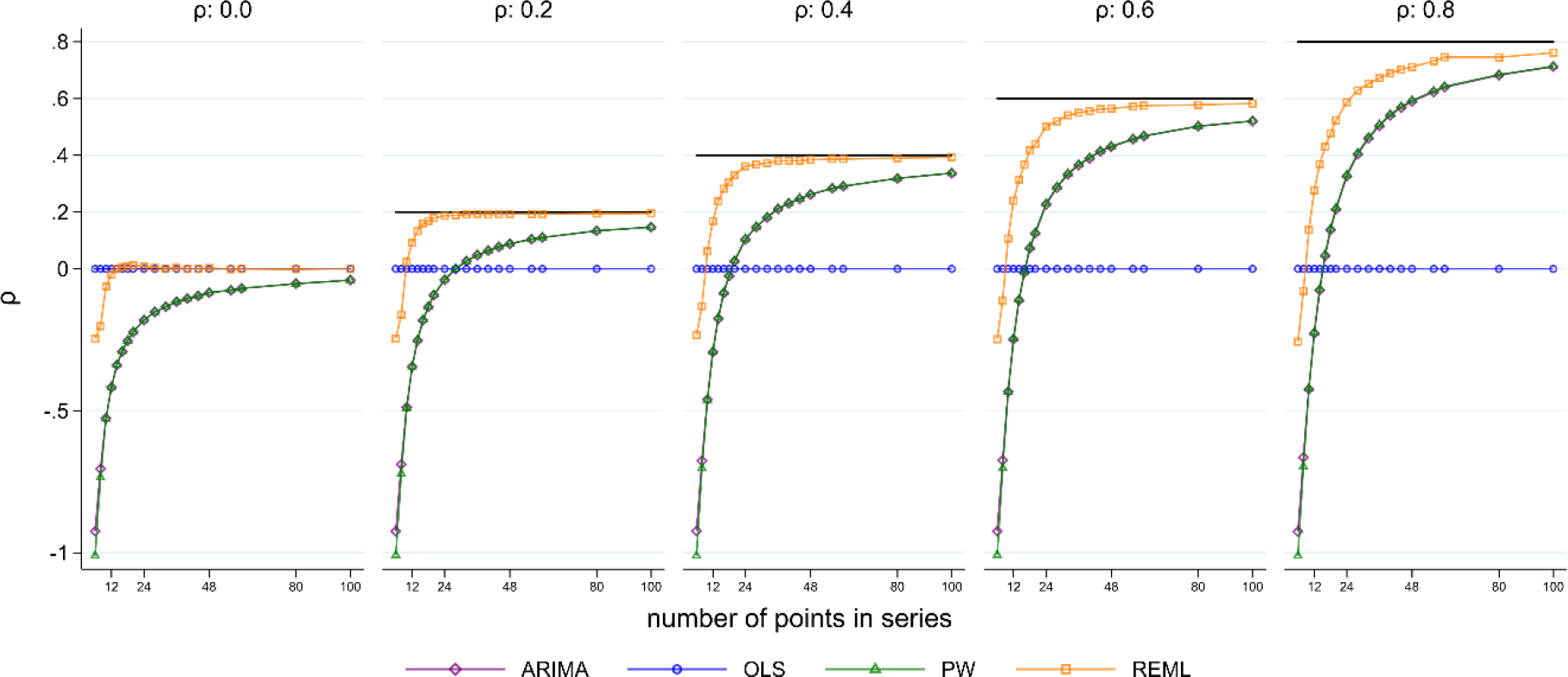
Autocorrelation coefficient estimates. The horizontal axis shows the length of the time series. The vertical axis shows the mean estimate of the autocorrelation coefficient across 10,000 simulations. The five plots display the results for different values of autocorrelation ranging from 0 to 0.8 (the true value of autocorrelation is shown by a horizontal black line). Each coloured point shows the mean autocorrelation estimate for a given combination of true autocorrelation coefficient and number of points in the data series. The simulation combination presented is for a level change of 2 and slope change of 0.1; however, other combinations give similar results. Abbreviations: ARIMA, autoregressive integrated moving average; OLS, ordinary least squares; PW, Prais-Winsten; REML, restricted maximum likelihood.

#### 5.5.1 Durbin-Watson test for autocorrelation

The DW test for detecting autocorrelation performed poorly except for long data series and large underlying values of autocorrelation (Figure 11). For series of moderate length (i.e. 48 points), with an underlying autocorrelation of 0.2, the DW test gave an “inconclusive” result in 30% of the simulations, incorrectly gave a value of no autocorrelation in 63% of the simulations, and only correctly identified that there was autocorrelation in 7% of the simulations. For shorter length series the percentage of simulations in which autocorrelation was correctly identified decreased (for a series length of 24 even at extreme magnitudes of autocorrelation (i.e. 0.8) positive autocorrelation was reported in only 26% of the simulations). For very short length series (fewer than 12 data points) the DW test gave an “inconclusive” result in over 75% of the simulations for all values of autocorrelation and always failed to identify that autocorrelation was present.

**Figure 11:**
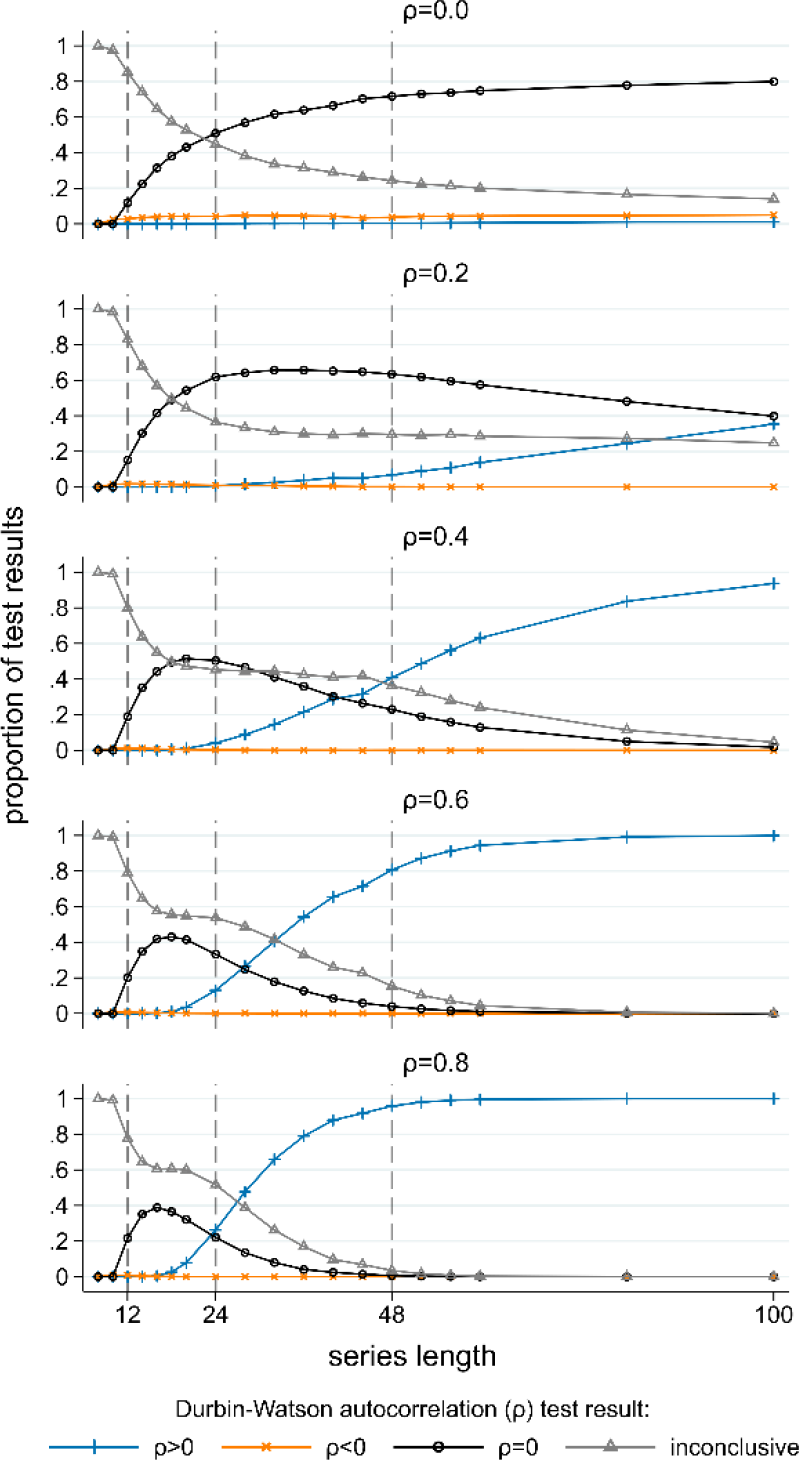
Durbin-Watson tests for autocorrelation. For each combination of length of data series and true magnitude of autocorrelation the Durbin Watson test results from 10,000 simulated data sets are summarised. The horizontal axis is the length of the data series, the vertical axis is the proportion of results indicating: ρ > 0 (blue), ρ < 0, (orange) ρ = 0 (black) and an inconclusive test (grey). Each graph shows results for a different magnitude of autocorrelation. The simulation combination presented is for a level change of 2 and slope change of 0.1; however, other combinations give similar results.

### 5.6 Convergence of estimation methods

The number of the 10,000 simulations in which the estimation methods converged is presented in Supplementary 1.10. Most methods had no numerical convergence issues. The PW model failed to converge a small number of times (less than 7% of simulations) when there were only three data points pre- and post-interruption. The REML model regularly failed to converge (approximately 70% convergence) for short data series (fewer than 12 data points) at all values of autocorrelation, however convergence improved substantially as the number of points in the series increased. In addition, convergence issues for REML occurred more frequently for higher values of autocorrelation, unless the series length was large.

## 6 Analysis of motivating example

We re-analysed the ITS study (introduced in Section 2) using each of the statistical methods evaluated in the simulation study to estimate the effect of terminal room cleaning with dilute bleach on *C difficile* rates. Estimates of level and slope change (along with their confidence intervals and p-values) and autocorrelation are presented in Table 4. The point estimates for level and slope change are similar across methods, but notably, the width of the confidence intervals vary considerably. The confidence intervals are narrower for OLS, NW and PW, but wider for REML (with and without the Satterthwaite adjustment) and ARIMA. For level change, this led to corresponding p-values that ranged from 0.002 to 0.095; and for the slope change, p-values ranging from 0.069 to 0.531. Estimates of autocorrelation also varied, with REML yielding an estimate of 0.23, while ARIMA and PW yielded much lower estimates of 0.07. The DW statistic was 1.86, indicating no autocorrelation. Such differences in confidence interval width and p-values may impact on the interpretation of the results.

**Table 4:**
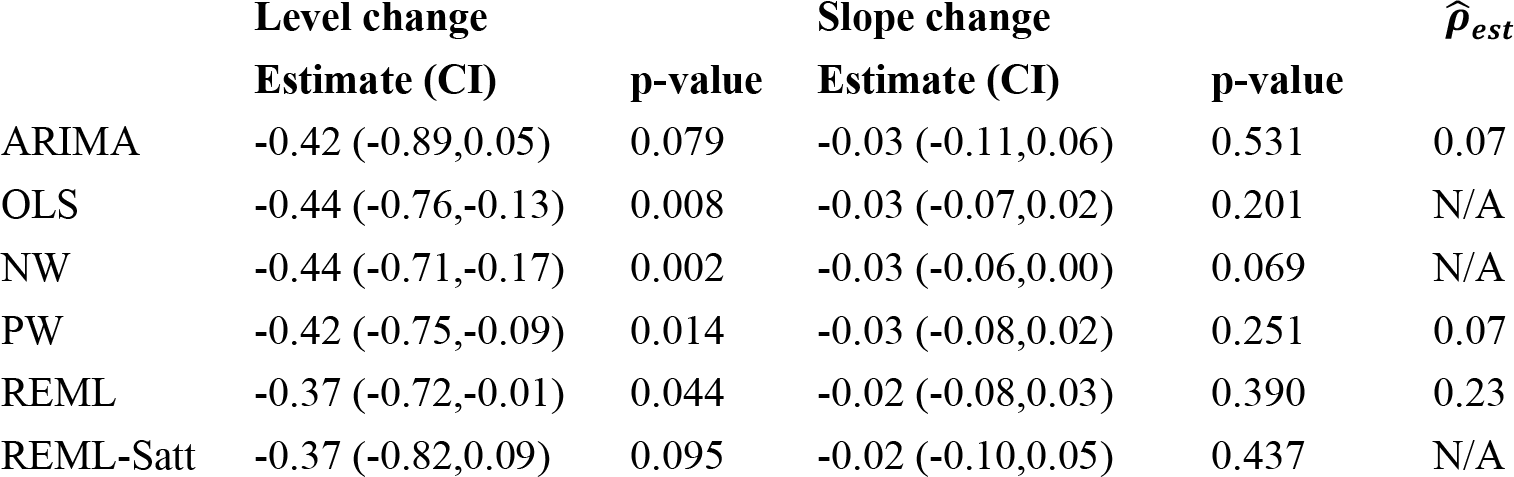
Level- and slope-change point estimates with 95% confidence intervals (CIs), p-values and estimate of magnitude of lag-1 autocorrelation 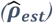 from C difficile infection data using a range of statistical methods. Abbreviations: ARIMA, autoregressive integrated moving average; OLS, ordinary least squares; NW, Newey-West; PW, Prais-Winsten; REML, restricted maximum likelihood; Satt, Satterthwaite.

| | Level change | | Slope change | | $\hat{\rho}_{est}$ |
| --- | --- | --- | --- | --- | --- |
|  | Estimate (CI) | p-value | Estimate (CI) | p-value |  |
| ARIMA | -0.42 (-0.89,0.05) | 0.079 | -0.03 (-0.11,0.06) | 0.531 | 0.07 |
| OLS | -0.44 (-0.76,-0.13) | 0.008 | -0.03 (-0.07,0.02) | 0.201 | N/A |
| NW | -0.44 (-0.71,-0.17) | 0.002 | -0.03 (-0.06,0.00) | 0.069 | N/A |
| PW | -0.42 (-0.75,-0.09) | 0.014 | -0.03 (-0.08,0.02) | 0.251 | 0.07 |
| REML | -0.37 (-0.72,-0.01) | 0.044 | -0.02 (-0.08,0.03) | 0.390 | 0.23 |
| REML-Satt | -0.37 (-0.82,0.09) | 0.095 | -0.02 (-0.10,0.05) | 0.437 | N/A |

## 7 Discussion

### 7.1 Summary and discussion of key findings

Interrupted time series studies are commonly used to evaluate the effects of interventions or exposures. The results of our simulation study provide insight into how a set of statistical methods perform under a range of scenarios which included different level and slope changes, varying lengths of series and magnitudes of autocorrelation. We chose to examine statistical methods that are commonly used in practice for interrupted time series studies^1-4 6^, and those performing well in the general, non-interrupted, time series literature^10 14^.

Not surprisingly, we found that the statistical methods all yielded unbiased estimates of both level and slope change for all values of model shape, length of series and autocorrelation. Confidence interval coverage, however, was generally below the nominal 95% level, except in particular circumstances for specific methods. The REML method with and without the Satterthwaite adjustment had improved confidence interval coverage compared with the other statistical methods. An exception to this was for very small series (fewer than 12 points), where the OLS method had better coverage than the other methods, even in the presence of large underlying autocorrelation. Coverage improved for most methods with increasing series length (with the exception of OLS and NW in some circumstances). REML with the Satterthwaite adjustment to the d.f. was the only method that yielded at least the nominal level of confidence interval coverage, however it was overly conservative in some scenarios, with a resultant reduction in power compared with other methods.

Autocorrelation was systematically underestimated by all statistical methods, with estimates of autocorrelation being particularly biased (and often negative) for small time series (fewer than 24 points). This underestimation of autocorrelation had a detrimental impact on the estimates of SE, which were too small, and in turn, this led to confidence interval coverage that was less than the nominal 95% level. This can be seen in Figure 12 (level change) and Supplementary 1.11 (slope change), where a relationship between the magnitude of bias in the estimates of autocorrelation and confidence interval coverage is clearly evident. Ideally the confidence interval coverage should be at the nominal 95% level with no bias in autocorrelation (the intersection of the dashed lines in Figure 12). For short time series, the severe underestimation of autocorrelation led to poorer confidence interval coverage than had autocorrelation been ignored, as is the case with OLS.

**Figure 12:**
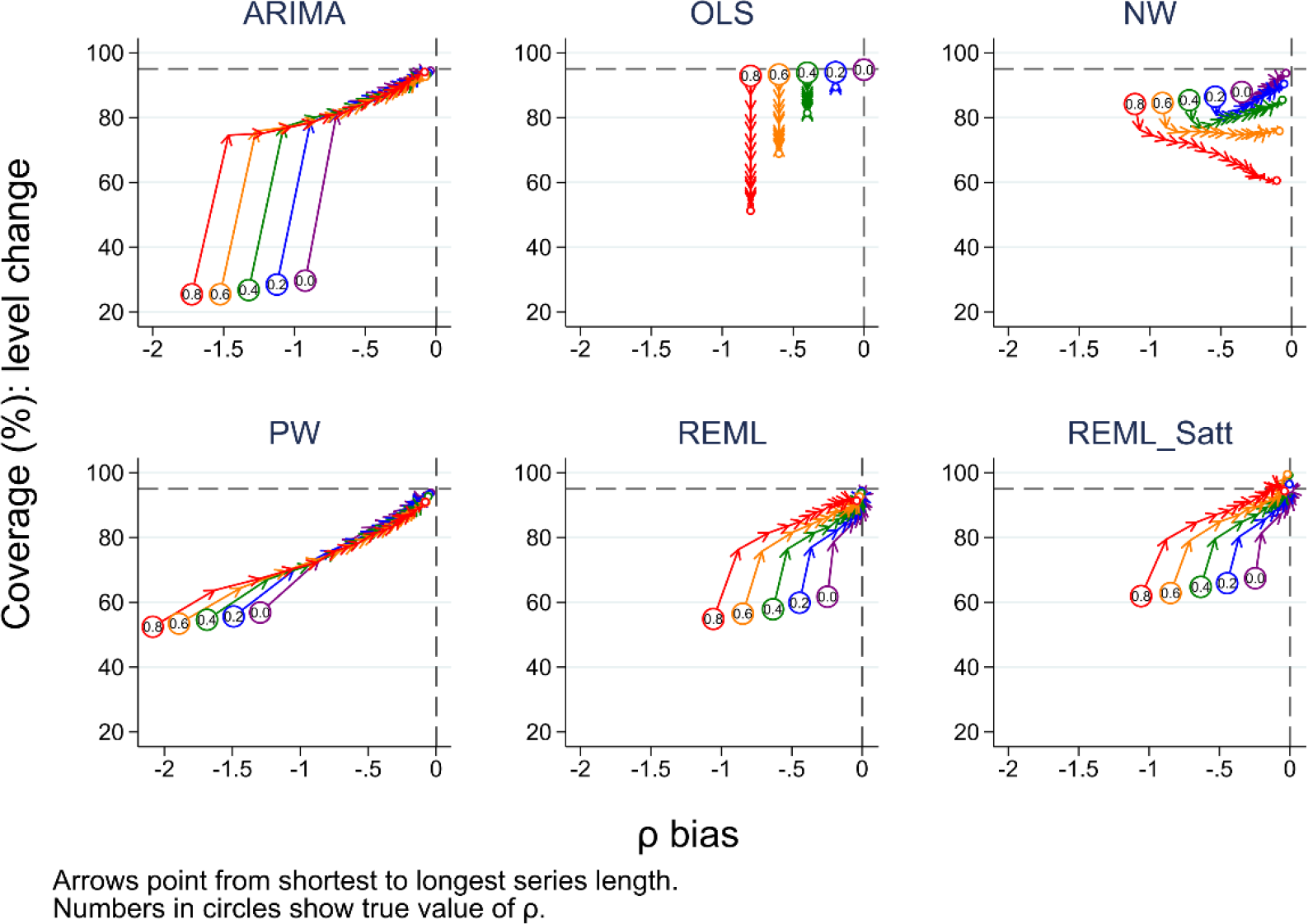
Bias in autocorrelation estimate versus coverage for level change. The horizontal axis shows the bias in the autocorrelation estimate. The vertical axis shows the percentage coverage. The horizontal dashed line indicates 95% coverage, the vertical dashed line indicates no bias in the estimate of autocorrelation. Each colour represents a different value of underlying autocorrelation, ranging from zero (purple) to 0.8 (red), with each value displayed in a circle at the smallest series length (six points). The arrows point from shortest to longest series length, with the small circles at the end of each line showing coverage at a series length of 100 data points. Each data point shows the mean value from 10,000 simulations for a given combination of autocorrelation coefficient and number of points in the series. The simulation combination presented is for a level change of 2 and slope change of 0.1; however, other combinations give similar results. Abbreviations: ARIMA, autoregressive integrated moving average; OLS, ordinary least squares; NW, Newey-West; PW, Prais-Winsten; REML, restricted maximum likelihood; Satt, Satterthwaite.

We included REML due to its potential to reduce bias in the variance parameters compared with maximum likelihood. Although the ARIMA model fitted in our simulations used maximum likelihood estimation, the model-based SEs were generally more similar to the empirical SEs for the ARIMA method compared with the REML method (where the model-based SEs were generally smaller than the empirical SEs), confidence interval coverage was generally better with REML. Further, the REML method yielded less biased estimates of autocorrelation than the other methods, even for small series lengths.

The only method to yield overly conservative confidence intervals was the REML with SW adjustment to the t-distribution d.f.. When deciding whether to use the Satterthwaite adjustment, consideration therefore needs to be made between the trade-off in the risk of type I and type II errors. A further issue we identified with the Satterthwaite adjustment was that the adjusted d.f. were very small in some series, leading to nonsensible confidence intervals. To limit this issue we set a minimum value of 2 for the d.f., but other choices could be adopted.

The DW test is the most commonly used test to identify autocorrelation and is often used when series are short^4 6^. Some authors use the test as part of a two-stage analysis strategy where they first test for autocorrelation, and depending on the result of the test, either use a method that attempts to adjust for autocorrelation or not. This type of two-stage approach is used in other contexts, such as testing for carryover in crossover trials. The findings of our simulation study underscore why such two stage approaches fail and are discouraged; namely, due to their failure to detect the presence of a statistic when it exists (i.e., their high type II error rate). In our case, we found that for short series (fewer than 12 data points), the DW test failed to identify autocorrelation when it was present, and for moderate length series (i.e. 48 points), with an underlying autocorrelation of 0.2, autocorrelation was only detected in 7% of the simulations.

### 7.2 Comparisons with other studies

We are not aware of other simulation studies that have examined the performance of statistical methods for interrupted time series studies. However, other simulation studies have investigated the performance of methods for general time series, and our findings align with these. Alpargu and Dutilleul^10^ concluded from their simulation study examining the performance of REML, PW and OLS for lag(1) time series data over a range of series lengths (from 10 to 200), that REML is to be preferred over OLS and PW in estimating slope parameters. Cheang and Reinsel^14^ examined the performance of ML and REML for estimating linear trends in lag(1) time series data of length 60 and 120 (both with and without seasonal components) and concluded that the REML estimator yielded better confidence interval coverage for the slope parameter, and less biased estimates of autocorrelation. Smith and McAleer^9^ examined the performance of the NW estimator for time series of length 100 with lags of 1, 3 and 10, and found that it underestimated the SEs of the slope parameter.

### 7.3 Strengths and Limitations

The strengths of our study include that we have used many combinations of parameter estimates and statistical methods. Our parameter values were informed by characteristics of real world ITS studies^4^. We planned and reported our study using the structured approach of Morris et al^24^ for simulation studies, and we generated a large number of data sets per combination to minimise MCSE.

As with all simulation studies, there are limitations to the applicability of findings. All data series were based on a random number generator and results may change given a different set of series, however, this is unlikely to be problematic given our MCSE was < 0.5% for all potential values of coverage and type I error rate. Our findings are only applicable to the scenarios in which they were generated, and so may not apply to ITS studies with different characteristics, such as unequal numbers of time points in the pre- and post-interruption segments, non-constant variance or different lags of autocorrelation.

### 7.4 Implications for practice

We found that all methods yielded unbiased estimates of the level and slope change, however, the methods differed in their performance in terms of confidence interval coverage and estimation of the autocorrelation parameter. Confidence interval coverage was primarily determined by the length of the time series and the underlying magnitude of autocorrelation. In practice, however, most analysts will only have knowledge of the length of the time series to guide in the choice of method. In rare cases, knowledge of the likely size of the underlying autocorrelation may be available from a previous long time series study in a similar context, which could help inform their choice. In our review of ITS studies investigating public health interruptions or exposures, the magnitude of autocorrelation was almost never explicitly specified (1%, 3/230 time series)^4^. Analysis of data extracted from the ITS studies included in this review using the REML method yielded a median autocorrelation 0.2 (IQR: 0 to 0.6, n=180); however, as shown from the simulation study, the estimates of autocorrelation (on which these summary statistics are based) are likely to be underestimated.

From the statistical methods and scenarios we examined, we found that for small time series (approximately 12 points or under), in the absence of a method that performs well adjusting for autocorrelation in such short series, OLS is the recommended method. For longer time series, REML is recommended. If the analyst has knowledge that the underlying autocorrelation is likely to be large, then using REML with the Satterthwaite adjustment may be advantageous. However, when the Satterthwaite adjustment yields d.f. lower than 2, we recommend replacing these with 2 to mitigate nonsensible confidence intervals. When REML doesn’t converge, ARIMA provides a reasonable alternative. Given most methods will yield confidence intervals that are too small, with type I error rates greater than 5%, borderline findings of statistical significance for the regression parameters should be cautiously interpreted; these may be due to chance rather than as a result of the interruption.

Estimates of autocorrelation from long series can be useful to inform sample size calculations and analytical decisions in future studies. We recommend reporting the REML estimates of the autocorrelation coefficient when possible. We only recommend using the DW test for detecting underlying autocorrelation in long time series (longer than 100 data points) and recommend against its use as part of a two-stage or stepwise approach to determine whether to use a statistical method that adjusts for autocorrelation.

In terms of study design, we recommend using at very minimum 24 points data points. With this number of points, confidence interval coverage close to the nominal 95% level can be achieved using REML with the Satterthwaite adjustment (when underlying autocorrelation is between 0 and 0.6). With fewer data points, poor confidence interval coverage is likely, irrespective of method.

### 7.5 Implications for future research

Although we investigated the statistical methods most commonly observed in reviews of ITS studies^1-4 6^, there is scope for further research examining other statistical methods, such as robust methods^27^ or Bayesian approaches where the uncertainty in the estimate of autocorrelation could be incorporated. We investigated one small-sample adjustment (Satterthwaite) though others, such as Kenward-Roger^28^, which adds a correction to the SE of regression parameter estimates, could also be examined. Further investigation of how the methods perform for scenarios other than those we investigated would be valuable. For example, when there are unequal numbers of points pre- and post-interruption, lags greater than 1, and where the autocorrelation and error variance differ between the pre and post interruption periods.

### 7.6 Conclusion

We undertook a simulation study to examine the performance of a set of statistical methods to analyse ITS data under a range of scenarios that included different level and slope changes, varying lengths of series and magnitudes of autocorrelation. We found that all methods yielded unbiased estimates of the level and slope change, however, the magnitude of autocorrelation was underestimated by all methods. This generally led to SEs that were too small and confidence interval coverage that was less than the nominal level. The DW test for the presence of autocorrelation performed poorly except for long series and large underlying autocorrelation. Care is needed when interpreting results from all methods, given the confidence intervals will generally be too narrow. Further research is required to determine and develop methods that perform well in the presence of autocorrelation, especially for short series.

### 7.7 Author contributions

JEM and ABF conceived the study and all authors contributed to its design. SLT designed and wrote the computer code and ran and analysed the simulations. SLT wrote the first draft of the manuscript, with contributions from JEM. SLT, JEM, AK, ABF, MT contributed to revisions of the manuscript and take public responsibility for its content.

## Data Availability

The data that supports the findings of this study were generated by the simulation code
available in the supplementary material of this article.

## 7.10 Acknowledgements

This work forms part of SLT’s PhD, which is supported by an Australian Postgraduate Award administered through Monash University, Australia.

## 7.11 Data Availability Statement

The data that supports the findings of this study were generated by the simulation code available in the supplementary material of this article.

### Appendix 1 Statistical method details

#### Appendix 1.1 Ordinary Least Squares

Model (1) can be written in a matrix form as:

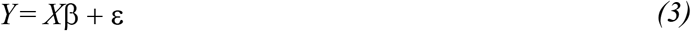

where *Y* and ε are *n × 1* vectors whose *t*^*th*^ element is *y*_*t*_ and *ε*_*t*_ respectively, *X* is the *n × 4* design matrix with *t*^′^*th* row(1,*t,D*_*t*_,*D*_*t*_*I*(*t* − *T*_1_), and *∈*_*t*_ ∼ *N*(0, *σ*^2^). The OLS estimator of *β* is 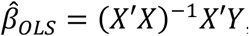, and 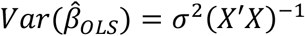.

#### Appendix 1.2 Newey West

The NW estimator (lag-1) of *β* is just the OLS estimator, 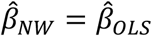, but with a sandwich variance estimator of the form

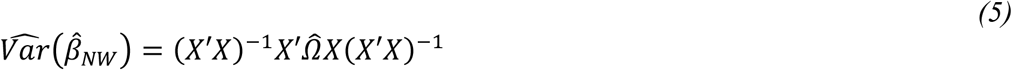

where:

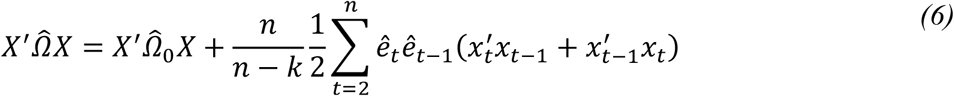

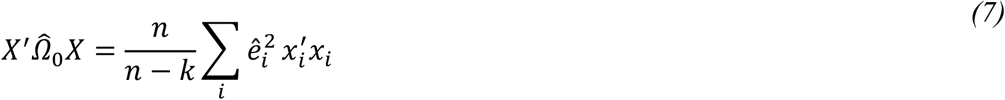

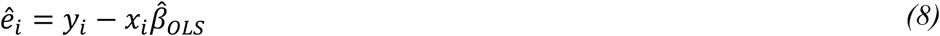

where *X* is the same *n* × 4 design matrix as specified for OLS above. The central term in the variance expression allows for empirical determination of autocorrelation and heteroskedasticity^15^.

#### Appendix 1.3 Generalised Least Squares

In the Cochrane-Orcutt and Prais-Winsten methods, from the equations (1) and (2), the dependent and independent variables are transformed to create a new model in which the error terms are uncorrelated:

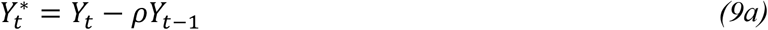

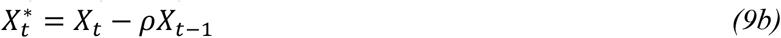

Then fit 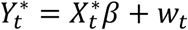, where

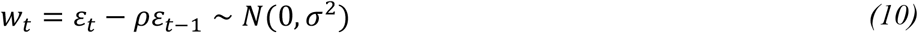

using OLS, and iterate until convergence.

Generally, the correlation is unknown, and must first be estimated. An estimate of autocorrelation at each iteration can be obtained using the OLS residuals *e*_*t*_ from fitting Equation (2) as above:

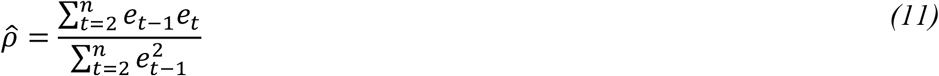

The CO method discards the first observation, while the PW method retains the first observation, but applies the following transformation^15^:

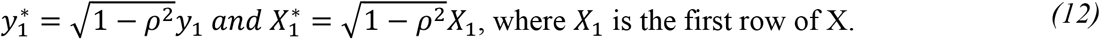

#### Appendix 1.4 Autoregressive integrated moving average

The ARIMA model includes parameters that model observations and error terms from previous time points. In an ARIMA model with first order autocorrelation only, i.e. ARIMA(1,0,0), equations (1) and (2) are fit simultaneously by maximum likelihood.

#### Appendix 1.5 Durbin-Watson test for autocorrelation

The Durbin-Watson test statistic is given by:

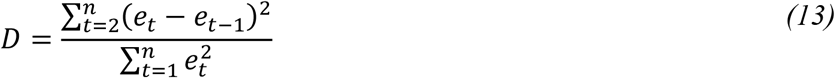

For test statistic values under two, D is compared to lower (*d*_*L*_) and upper (*d*_*U*_) bounds, leading to either a conclusive or inconclusive result. For test statistic values over two, 4-D is compared to the lower and upper bounds and a conclusive *H*_*alternative*_ indicates the presence of negative autocorrelation:

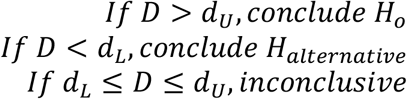

Lower (*d*_*L*_) and upper (*d*_*U*_) bounds can be found in tables online or in textbooks, e.g. Kutner et al^13^.

### Appendix 2 Definitions of performance measures

**Table 3:** Definitions of performance measures. Where θ represents the parameter under investigation, 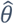being the estimate of that parameter,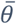 being the mean value of the estimate, n_sim_ being the number of simulations (in this study, 10,000), p_i_ being the p-value of estimate i and α being the significance level^22^.

| Performance measure | Definition | Estimate |
| --- | --- | --- |
| Bias | $E[\hat{\theta}] - \theta$ | $\frac{1}{n_{sim}} \sum_{i=1}^{n_{sim}} \hat{\theta}_i - \theta$ |
| Empirical standard error | $\sqrt{Var(\hat{\theta})}$ | $\sqrt{\frac{1}{n_{sim} - 1} \sum_{i=1}^{n_{sim}} (\hat{\theta}_i - \bar{\theta})^2}$ |
| Mean square error | $E[(\hat{\theta}_i - \theta)^2]$ | $\frac{1}{n_{sim}} \sum_{i=1}^{n_{sim}} (\hat{\theta}_i - \theta)^2$ |
| Coverage | $Pr(\hat{\theta}_{low} \leq \theta \leq \hat{\theta}_{upp})$ | $\frac{1}{n_{sim}} \sum_{i=1}^{n_{sim}} 1(\hat{\theta}_{low,i} \leq \theta \leq \hat{\theta}_{upp,i})$ |
| Power | $Pr(p_i \leq \alpha)$ | $\frac{1}{n_{sim}} \sum_{i=1}^{n_{sim}} 1(p_i \leq \alpha)$ |

