## Supplementary File 1 for "Evaluation of statistical methods used in the analysis of interrupted time series studies: a simulation study"

#### Supplementary 1.1 Interrupted time series graphs

The following, Figure S1, shows an example of a simulated data set using the parameterisation shown in equation 1, section 3.1.

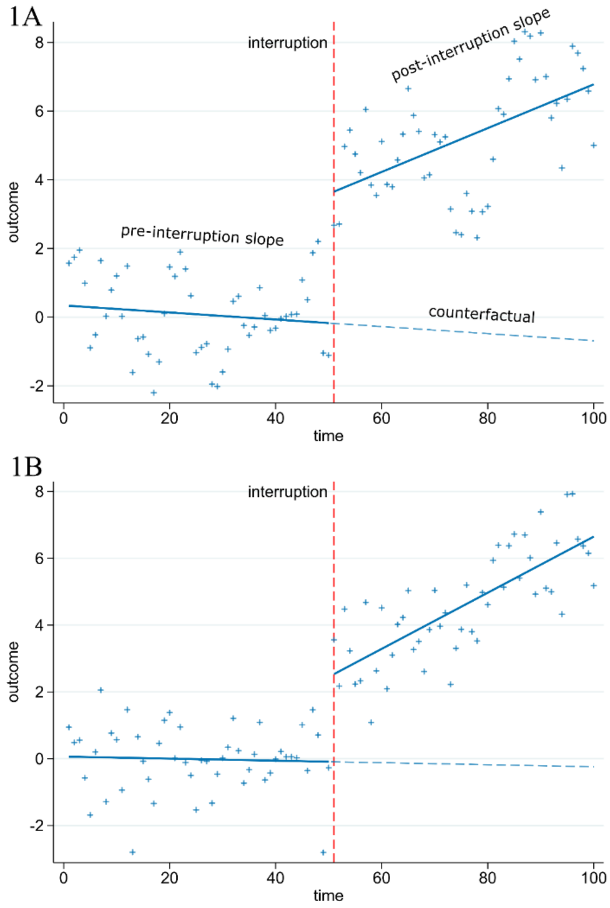


*Figure S1: An example of two simulated data sets. This figure was created using the model ($Y_t = \beta_0 + \beta_1 t + \beta_2 D_t + \beta_3 [t - T_I] D_t + \varepsilon_t$ where $\varepsilon_t = \rho\ \varepsilon_{t-1} + N(0,\sigma^2)$), with parameters $\beta_0 = 0$ (y-intercept), $\beta_1 = 0$ (pre-interruption slope), $\beta_2 = 2$ (level change at time of interruption), $\beta_3 = 0.1$ (slope change after interruption), and σ=1, $D_t$ is an indicator variable that is 0 for pre-interruption times and 1 for post-interruption times. In Figure 1A the autocorrelation (ρ) is 0.8, in Figure 1B it is 0.*

#### Supplementary 1.2 Slope change estimates

The following, Figure S2, shows the distribution of slope change estimates for parameters 2 for level change and 0.1 for slope change.

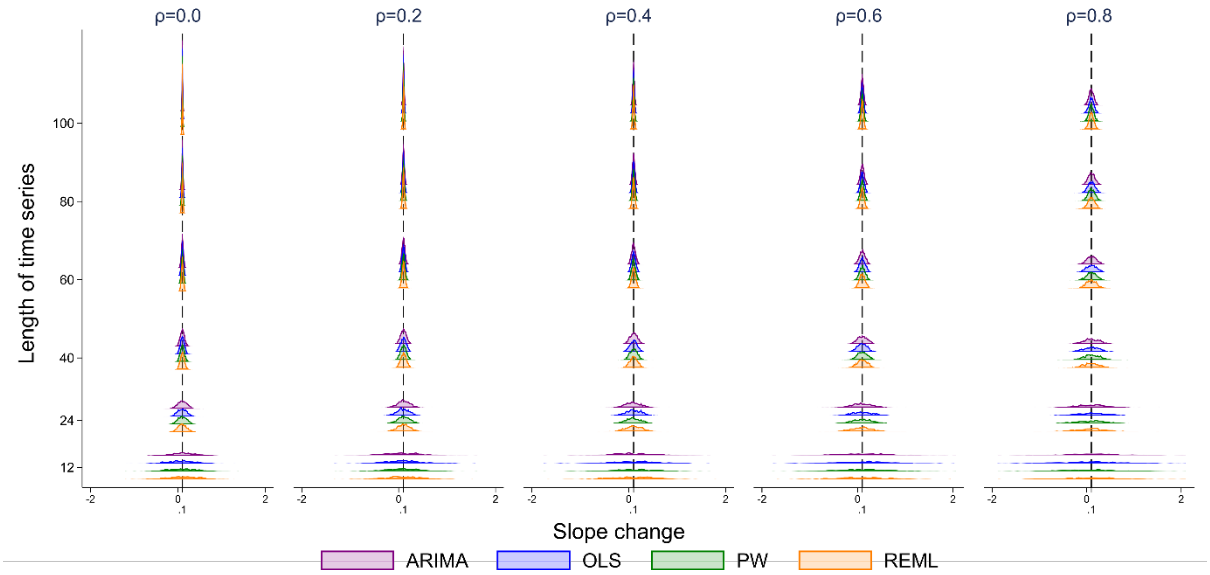


*Figure S2: Distributions of slope change estimates calculated from four statistical methods, from top to bottom: autoregressive integrated moving average (ARIMA) (purple), ordinary least squares regression (OLS) (blue), Prais-Winsten (PW) (green) and restricted maximum likelihood (REML) (orange). The vertical axis shows the length of the time series. The five vertical columns display the results for different values of autocorrelation. The vertical black line represents the true parameter value ($\beta_3$). Each subset of four curves shows the distribution from a different analysis method for a given combination of time series length and autocorrelation. The simulation combination presented is for a level change of 2 and slope change of 0.1; however, other combinations give similar results. The Satterthwaite adjustment to the REML method and the Newey-West adjustment to the OLS method do not impact the estimate of level or slope change, hence these parameter estimates are not shown.*

#### Supplementary 1.3 Differences between slope and level change parameters

The following nested loop plots (Rücker and Schwarzer 2014) show the similarities in estimates and performance measures across the eight different level and slope change parameters. Each statistical method is denoted using different coloured and shaped points across each combination of time series length, level change and slope change.

##### S 1.3.1 Level change bias

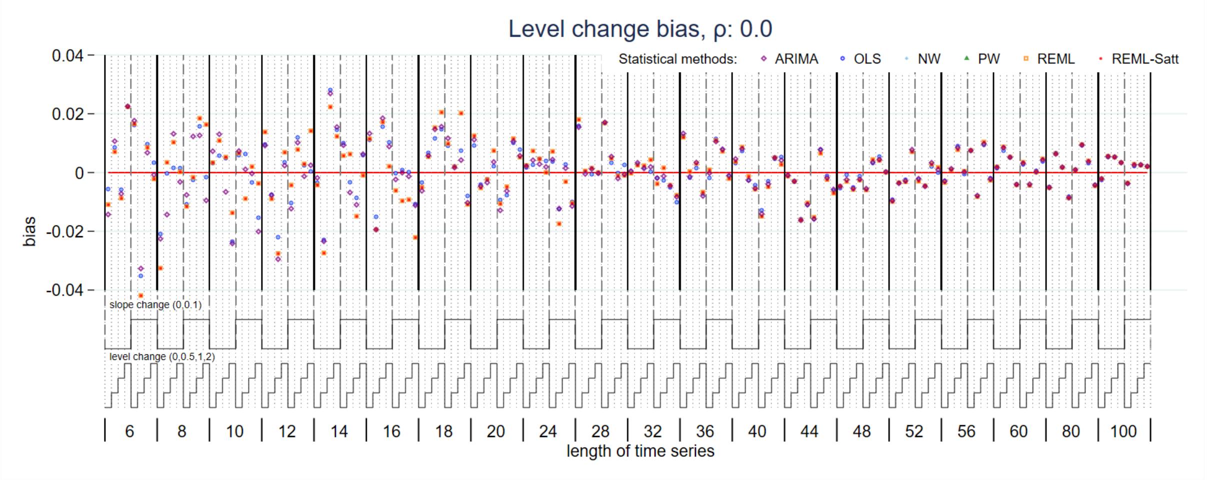


*Figure S3: Bias in level change estimate for magnitude of autocorrelation 0. Each data point shows the mean value from 10,000 simulations for a given combination of slope change, level change and length of time series. Abbreviations: ARIMA, autoregressive integrated moving average; OLS, ordinary least squares; NW, Newey-West; PW, Prais-Winsten; REML, restricted maximum likelihood; Satt, Satterthwaite.*

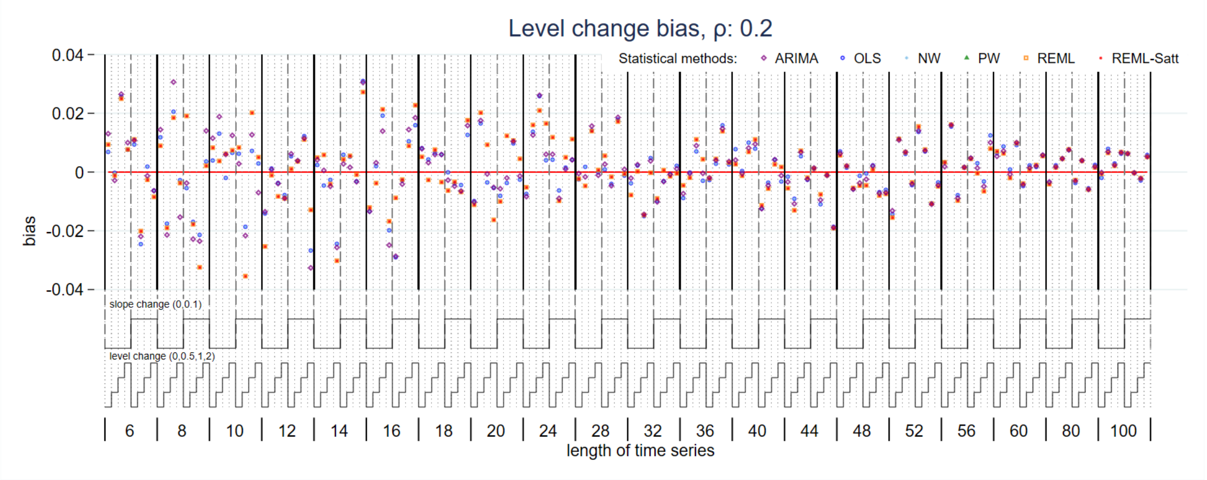


*Figure S4: Bias in level change estimate for magnitude of autocorrelation 0.2. Each data point shows the mean value from 10,000 simulations for a given combination of slope change, level change and length of time series. Abbreviations: ARIMA, autoregressive integrated moving average; OLS, ordinary least squares; NW, Newey-West; PW, Prais-Winsten; REML, restricted maximum likelihood; Satt, Satterthwaite.*

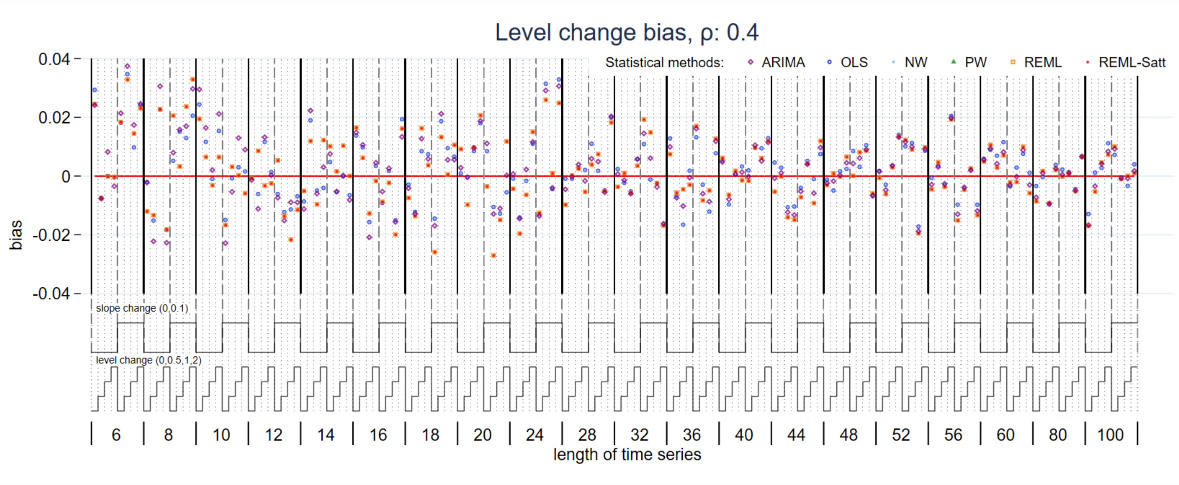


*Figure S5: Bias in level change estimate for magnitude of autocorrelation 0.4. Each data point shows the mean value from 10,000 simulations for a given combination of slope change, level change and length of time series. Abbreviations: ARIMA, autoregressive integrated moving average; OLS, ordinary least squares; NW, Newey-West; PW, Prais-Winsten; REML, restricted maximum likelihood; Satt, Satterthwaite.*

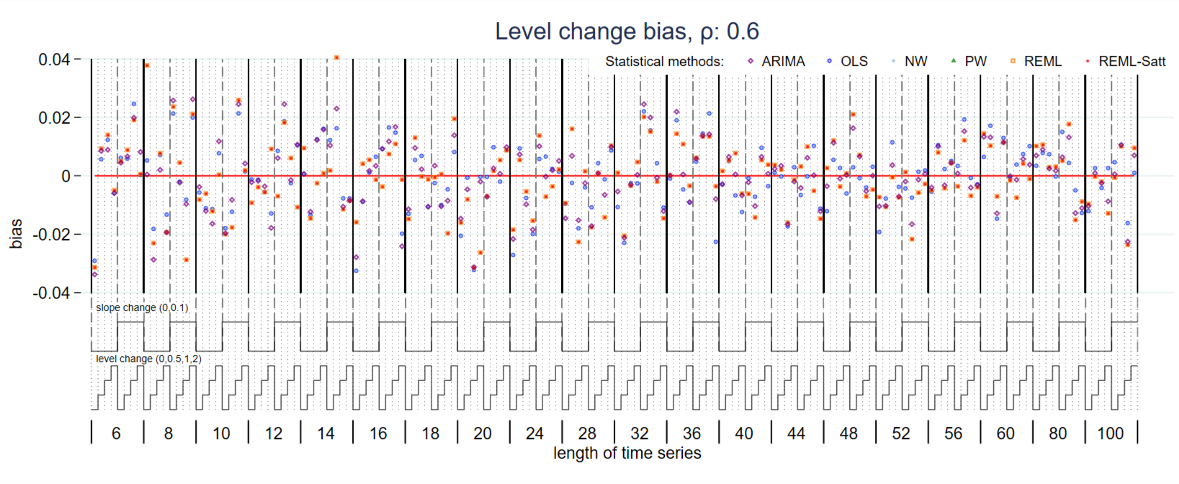


*Figure S6: Bias in level change estimate for magnitude of autocorrelation 0.6. Each data point shows the mean value from 10,000 simulations for a given combination of slope change, level change and length of time series. Abbreviations: ARIMA, autoregressive integrated moving average; OLS, ordinary least squares; NW, Newey-West; PW, Prais-Winsten; REML, restricted maximum likelihood; Satt, Satterthwaite.*

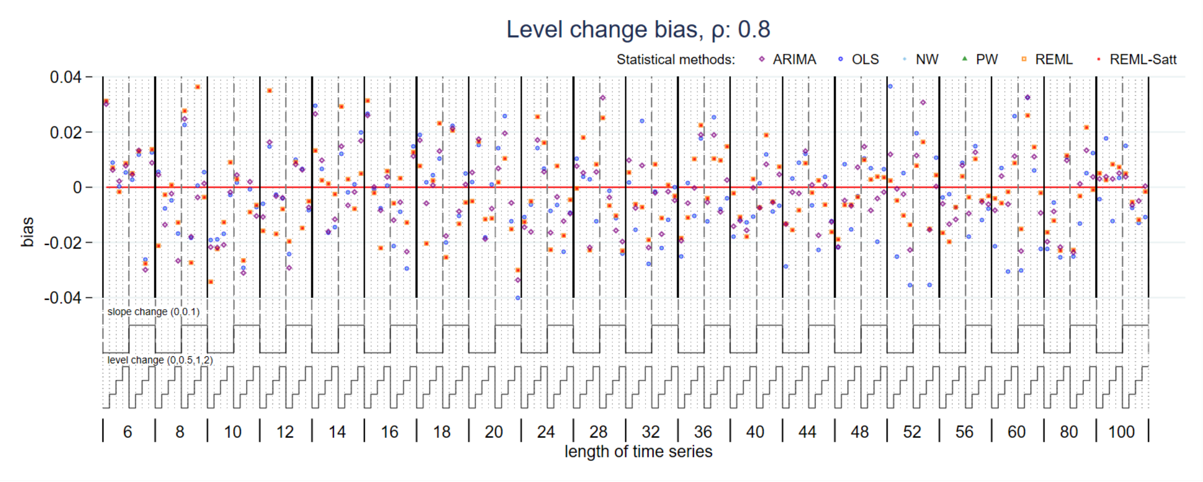


*Figure S7: Bias in level change estimate for magnitude of autocorrelation 0.8. Each data point shows the mean value from 10,000 simulations for a given combination of slope change, level change and length of time series. Abbreviations: ARIMA, autoregressive integrated moving average; OLS, ordinary least squares; NW, Newey-West; PW, Prais-Winsten; REML, restricted maximum likelihood; Satt, Satterthwaite.*

#### S 1.3.2 Slope change bias

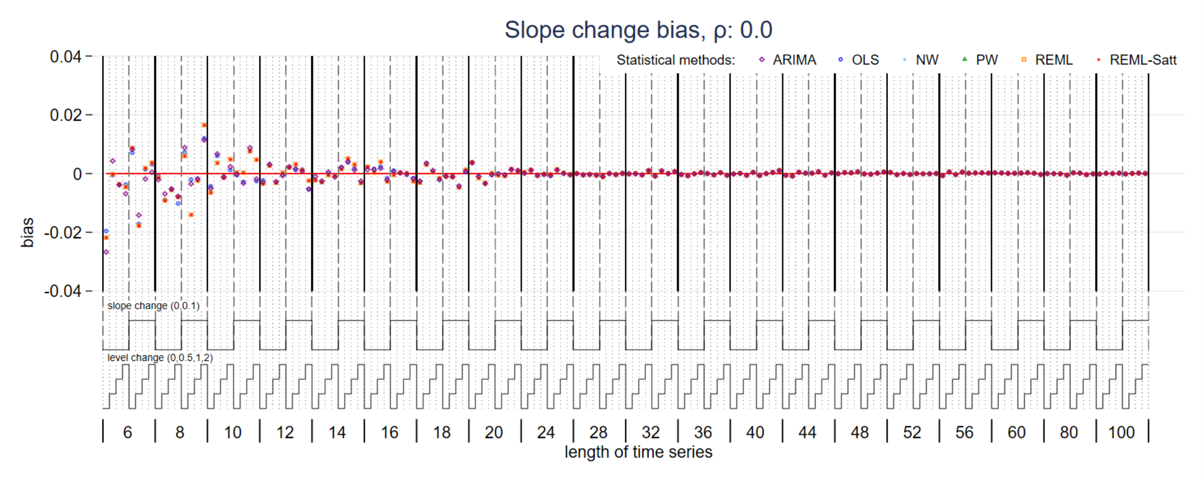


*Figure S8: Bias in slope change estimate for magnitude of autocorrelation 0. Each data point shows the mean value from 10,000 simulations for a given combination of slope change, level change and length of time series. Abbreviations: ARIMA, autoregressive integrated moving average; OLS, ordinary least squares; NW, Newey-West; PW, Prais-Winsten; REML, restricted maximum likelihood; Satt, Satterthwaite.*

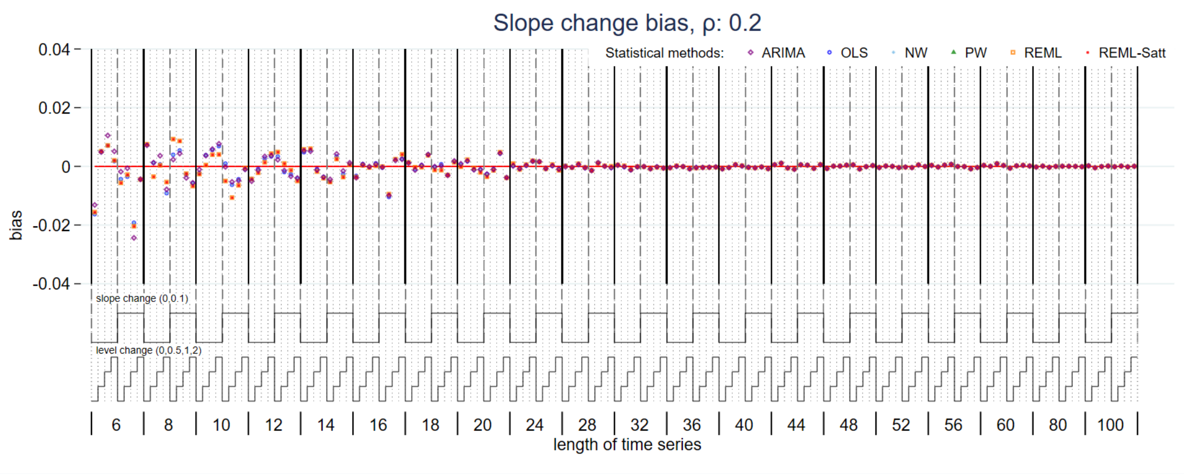


*Figure S9: Bias in slope change estimate for magnitude of autocorrelation 0.2. Each data point shows the mean value from 10,000 simulations for a given combination of slope change, level change and length of time series. Abbreviations: ARIMA, autoregressive integrated moving average; OLS, ordinary least squares; NW, Newey-West; PW, Prais-Winsten; REML, restricted maximum likelihood; Satt, Satterthwaite.*

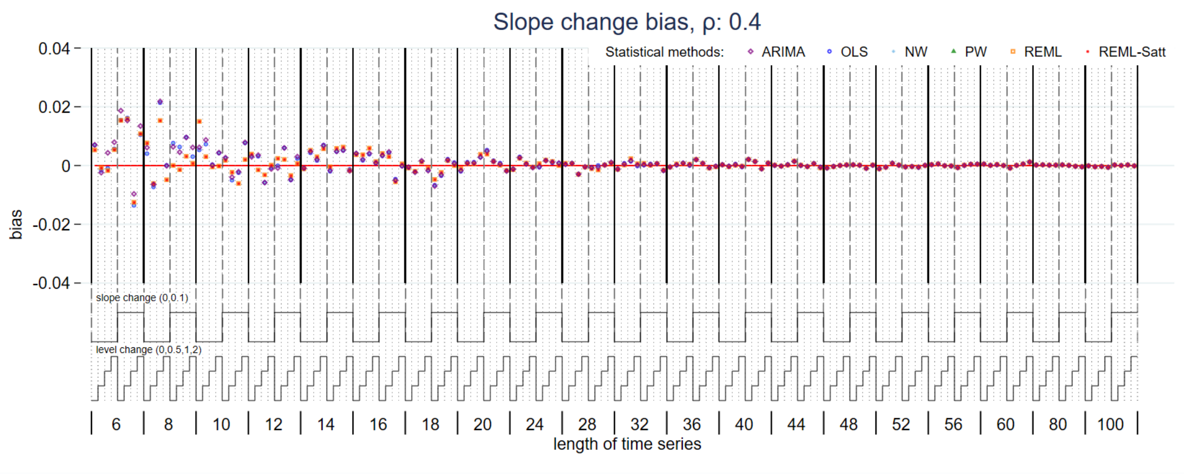


*Figure S10: Bias in slope change estimate for magnitude of autocorrelation 0.4. Each data point shows the mean value from 10,000 simulations for a given combination of slope change, level change and length of time series. Abbreviations: ARIMA, autoregressive integrated moving average; OLS, ordinary least squares; NW, Newey-West; PW, Prais-Winsten; REML, restricted maximum likelihood; Satt, Satterthwaite.*

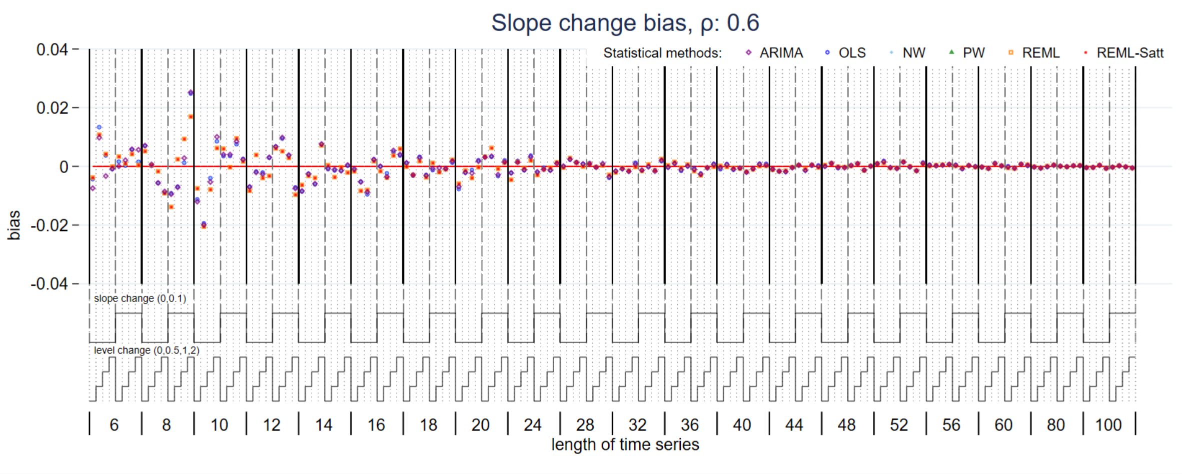


*Figure S11: Bias in slope change estimate for magnitude of autocorrelation 0.6. Each data point shows the mean value from 10,000 simulations for a given combination of slope change, level change and length of time series. Abbreviations: ARIMA, autoregressive integrated moving average; OLS, ordinary least squares; NW, Newey-West; PW, Prais-Winsten; REML, restricted maximum likelihood; Satt, Satterthwaite.*

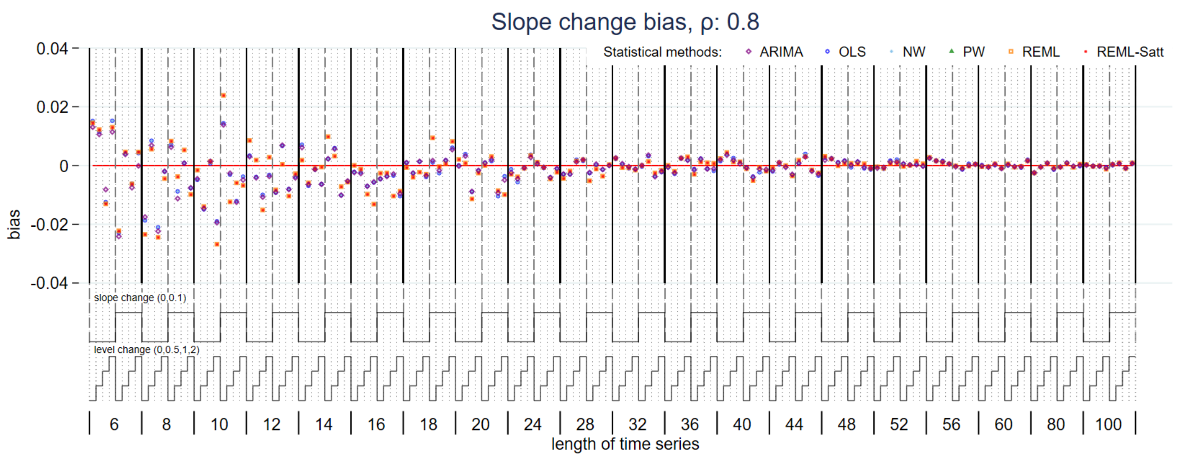


*Figure S12: Bias in slope change estimate for magnitude of autocorrelation 0.8. Each data point shows the mean value from 10,000 simulations for a given combination of slope change, level change and length of time series. Abbreviations: ARIMA, autoregressive integrated moving average; OLS, ordinary least squares; NW, Newey-West; PW, Prais-Winsten; REML, restricted maximum likelihood; Satt, Satterthwaite.*

##### S 1.3.3 Level change empirical standard error

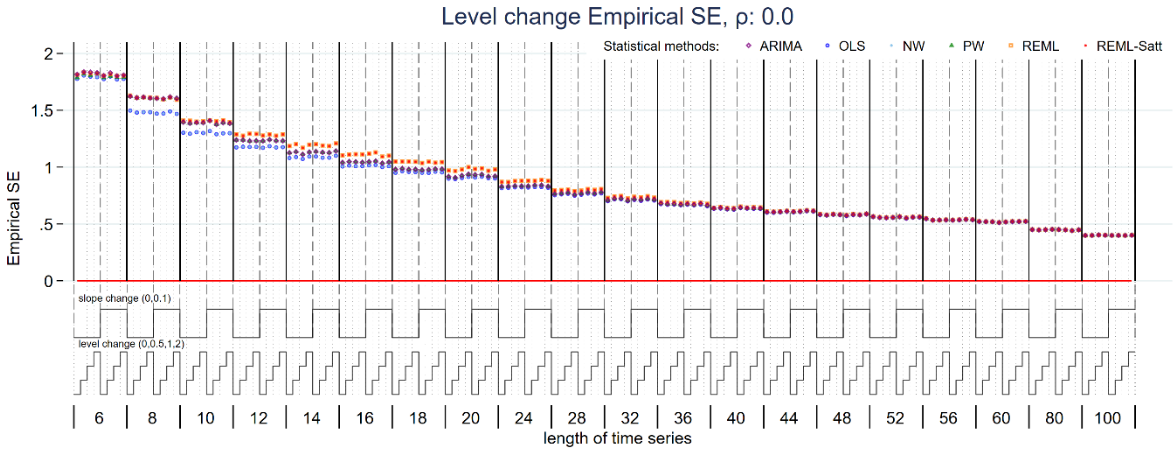


*Figure S13: Empirical standard error of level change estimate for magnitude of autocorrelation 0. Each data point shows the mean value from 10,000 simulations for a given combination of slope change, level change and length of time series. Abbreviations: ARIMA, autoregressive integrated moving average; OLS, ordinary least squares; NW, Newey-West; PW, Prais-Winsten; REML, restricted maximum likelihood; Satt, Satterthwaite.*

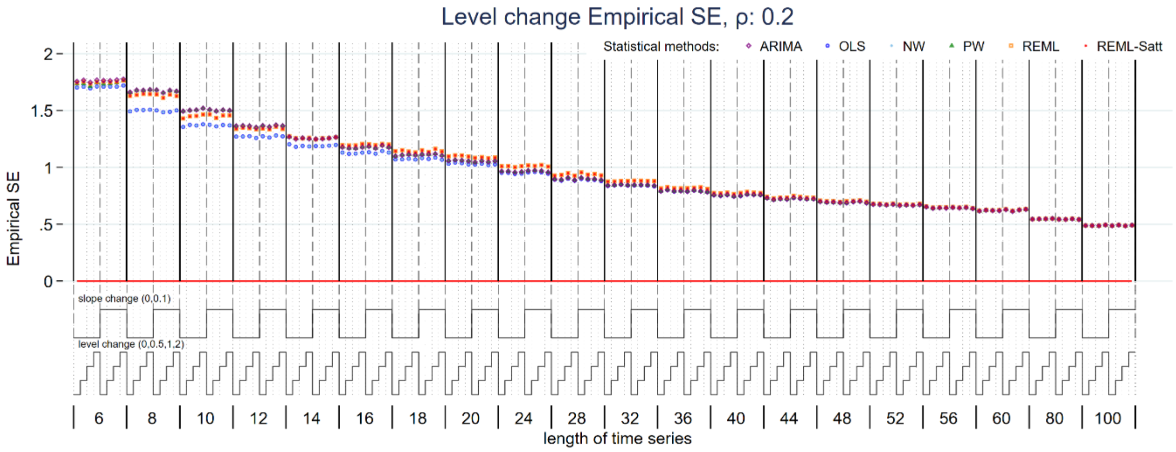


*Figure S14: Empirical standard error of level change estimate for magnitude of autocorrelation 0.2. Each data point shows the mean value from 10,000 simulations for a given combination of slope change, level change and length of time series. Abbreviations: ARIMA, autoregressive integrated moving average; OLS, ordinary least squares; NW, Newey-West; PW, Prais-Winsten; REML, restricted maximum likelihood; Satt, Satterthwaite.*

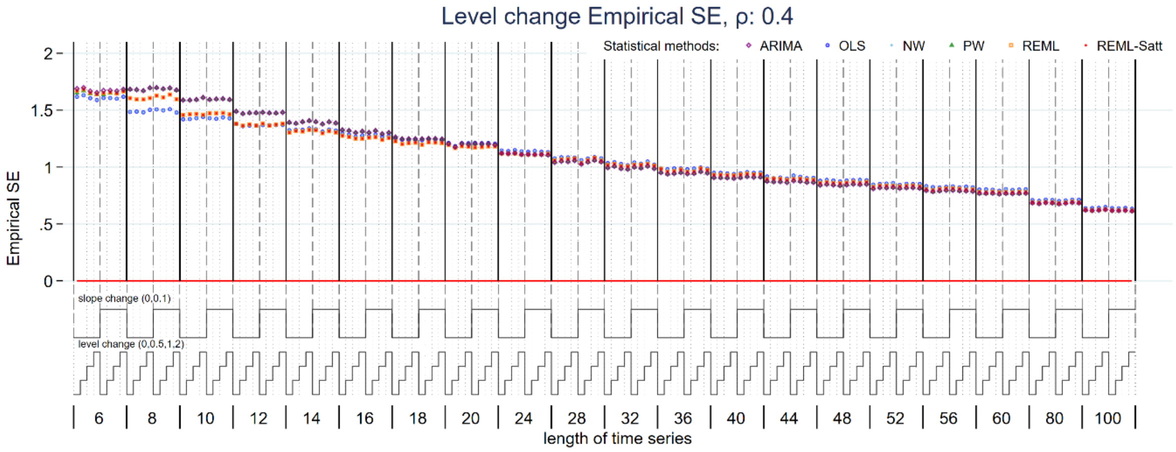


*Figure S15: Empirical standard error of level change estimate for magnitude of autocorrelation 0.4. Each data point shows the mean value from 10,000 simulations for a given combination of slope change, level change and length of time series. Abbreviations: ARIMA, autoregressive integrated moving average; OLS, ordinary least squares; NW, Newey-West; PW, Prais-Winsten; REML, restricted maximum likelihood; Satt, Satterthwaite.*

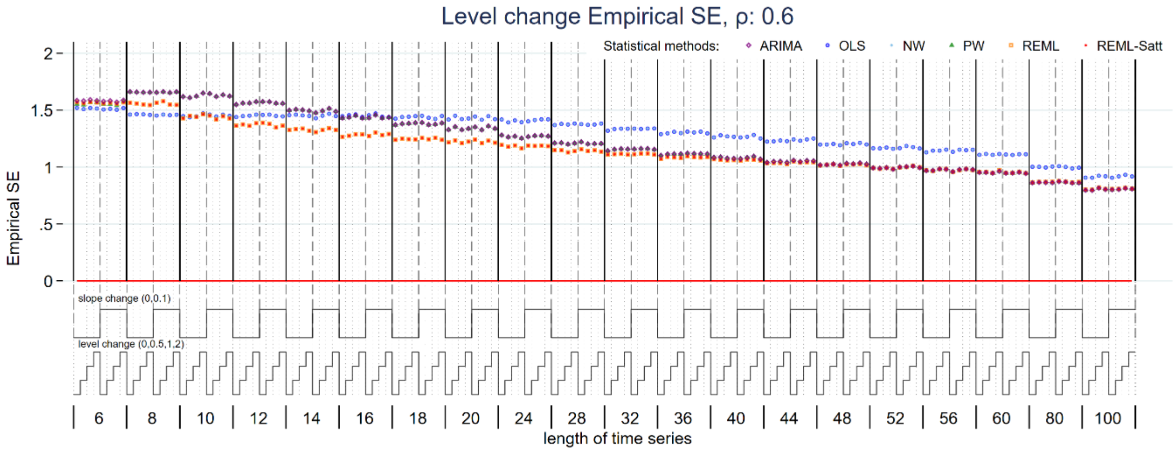


*Figure S16: Empirical standard error of level change estimate for magnitude of autocorrelation 0.6. Each data point shows the mean value from 10,000 simulations for a given combination of slope change, level change and length of time series. Abbreviations: ARIMA, autoregressive integrated moving average; OLS, ordinary least squares; NW, Newey-West; PW, Prais-Winsten; REML, restricted maximum likelihood; Satt, Satterthwaite.*

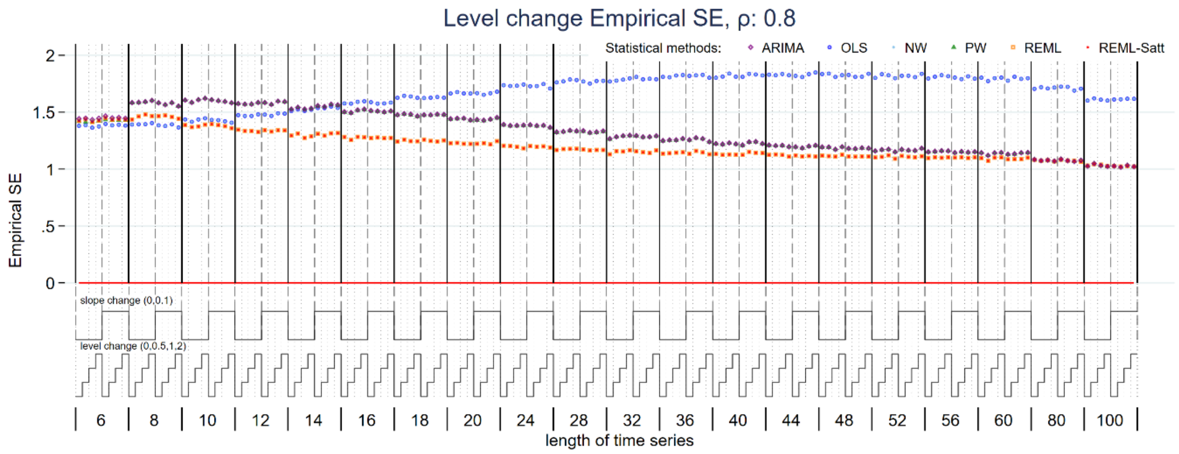


*Figure S17: Empirical standard error of level change estimate for magnitude of autocorrelation 0.8. Each data point shows the mean value from 10,000 simulations for a given combination of slope change, level change and length of time series. Abbreviations: ARIMA, autoregressive integrated moving average; OLS, ordinary least squares; NW, Newey-West; PW, Prais-Winsten; REML, restricted maximum likelihood; Satt, Satterthwaite.*

##### S 1.3.4 Slope change empirical standard error

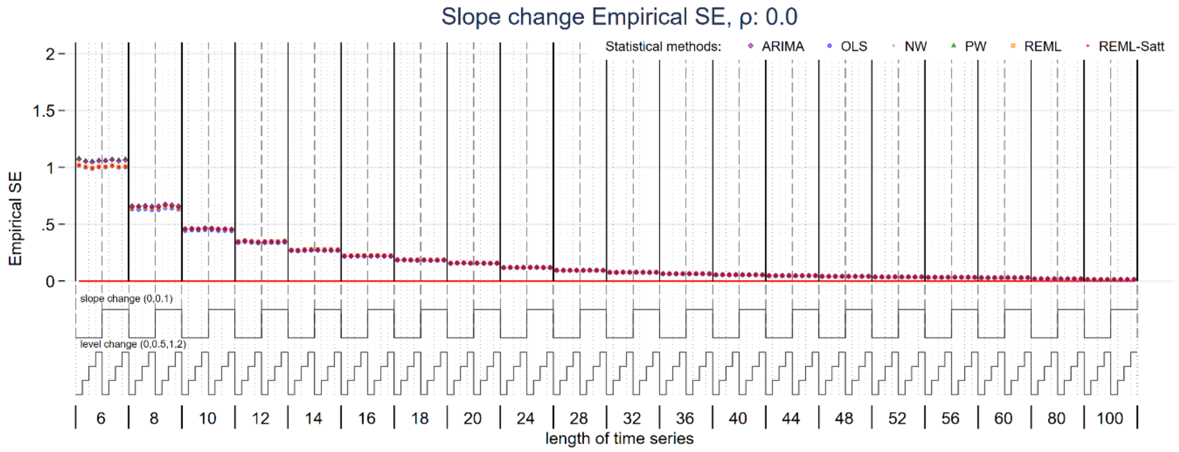


*Figure S18: Empirical standard error of slope change estimate for magnitude of autocorrelation 0. Each data point shows the mean value from 10,000 simulations for a given combination of slope change, level change and length of time series. Abbreviations: ARIMA, autoregressive integrated moving average; OLS, ordinary least squares; NW, Newey-West; PW, Prais-Winsten; REML, restricted maximum likelihood; Satt, Satterthwaite.*

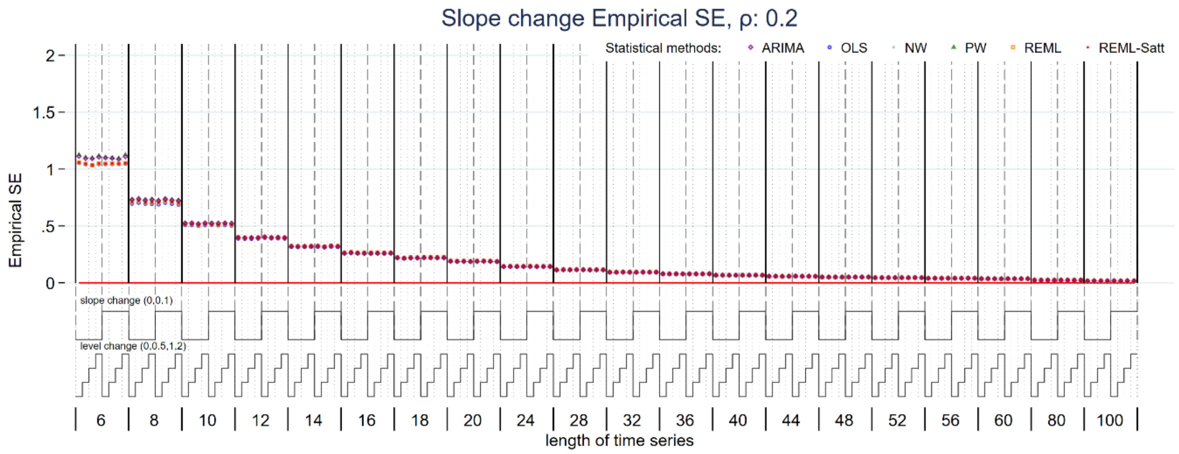


*Figure S19: Empirical standard error of slope change estimate for magnitude of autocorrelation 0.2. Each data point shows the mean value from 10,000 simulations for a given combination of slope change, level change and length of time series. Abbreviations: ARIMA, autoregressive integrated moving average; OLS, ordinary least squares; NW, Newey-West; PW, Prais-Winsten; REML, restricted maximum likelihood; Satt, Satterthwaite.*

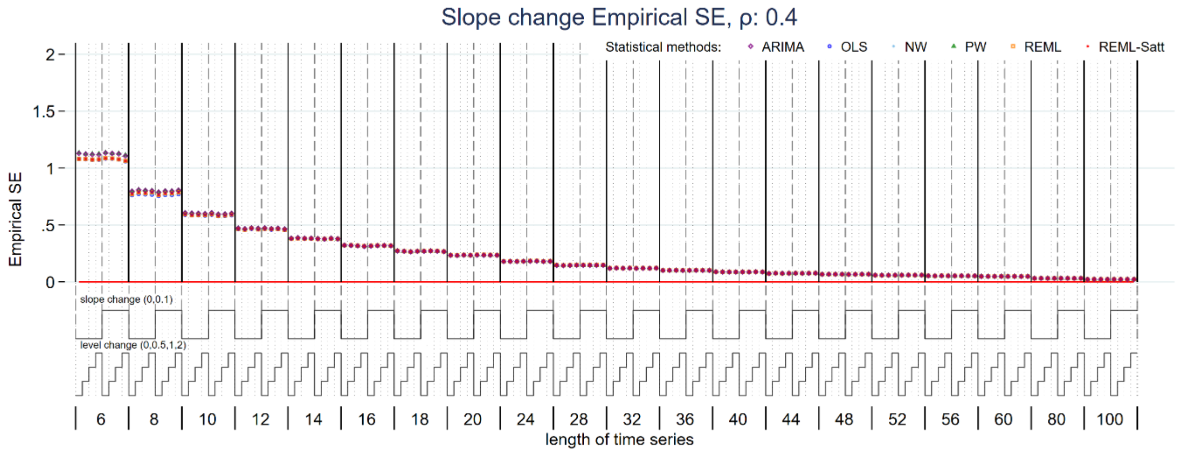


*Figure S20: Empirical standard error of slope change estimate for magnitude of autocorrelation 0.4. Each data point shows the mean value from 10,000 simulations for a given combination of slope change, level change and length of time series. Abbreviations: ARIMA, autoregressive integrated moving average; OLS, ordinary least squares; NW, Newey-West; PW, Prais-Winsten; REML, restricted maximum likelihood; Satt, Satterthwaite.*

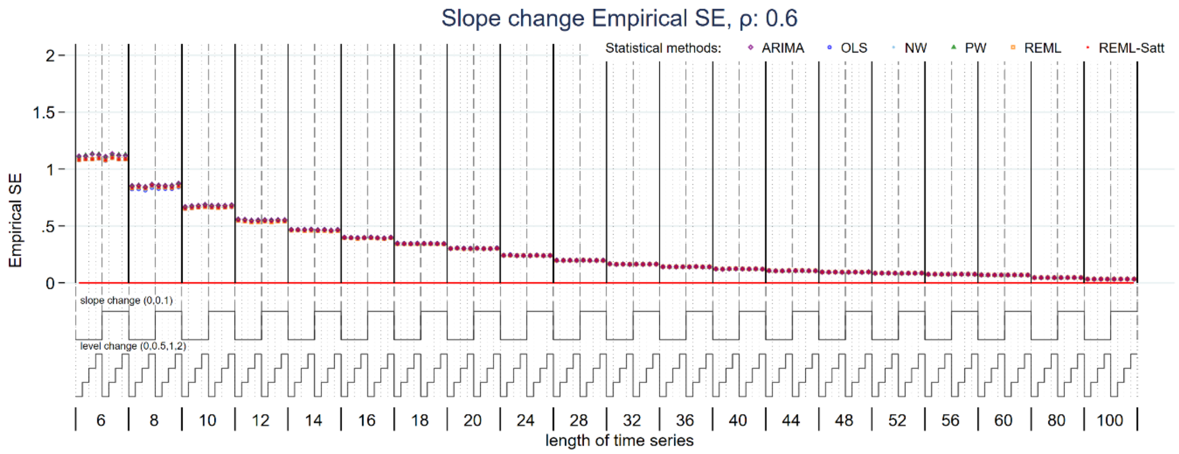


*Figure S21: Empirical standard error of slope change estimate for magnitude of autocorrelation 0.6. Each data point shows the mean value from 10,000 simulations for a given combination of slope change, level change and length of time series. Abbreviations: ARIMA, autoregressive integrated moving average; OLS, ordinary least squares; NW, Newey-West; PW, Prais-Winsten; REML, restricted maximum likelihood; Satt, Satterthwaite.*

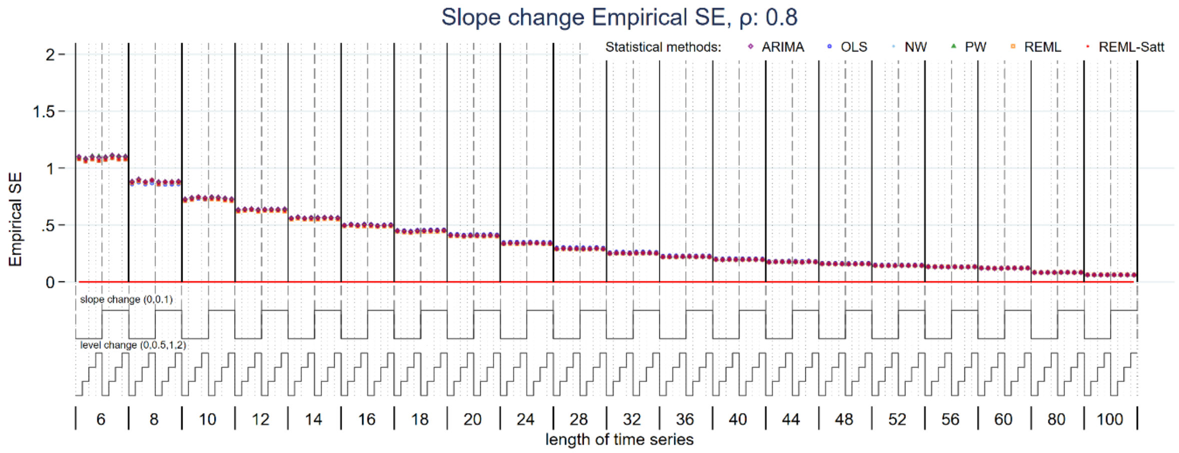


*Figure S22: Empirical standard error of slope change estimate for magnitude of autocorrelation 0.8. Each data point shows the mean value from 10,000 simulations for a given combination of slope change, level change and length of time series. Abbreviations: ARIMA, autoregressive integrated moving average; OLS, ordinary least squares; NW, Newey-West; PW, Prais-Winsten; REML, restricted maximum likelihood; Satt, Satterthwaite.*

#### S 1.3.5 Level change model-based standard error

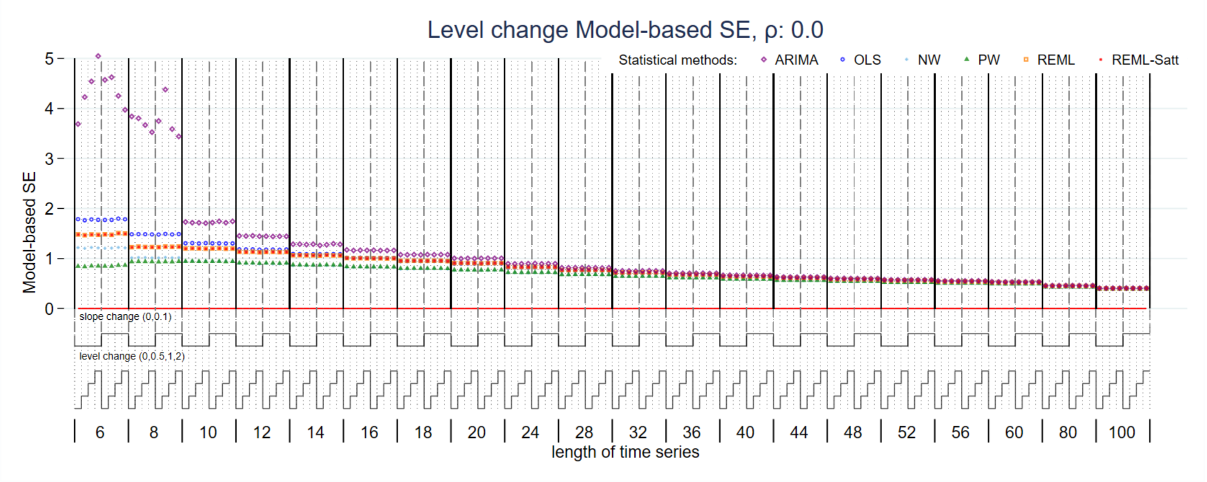


*Figure S23: Model-based standard error of level change estimate for magnitude of autocorrelation 0. Each data point shows the mean value from 10,000 simulations for a given combination of slope change, level change and length of time series. Abbreviations: ARIMA, autoregressive integrated moving average; OLS, ordinary least squares; NW, Newey-West; PW, Prais-Winsten; REML, restricted maximum likelihood; Satt, Satterthwaite.*

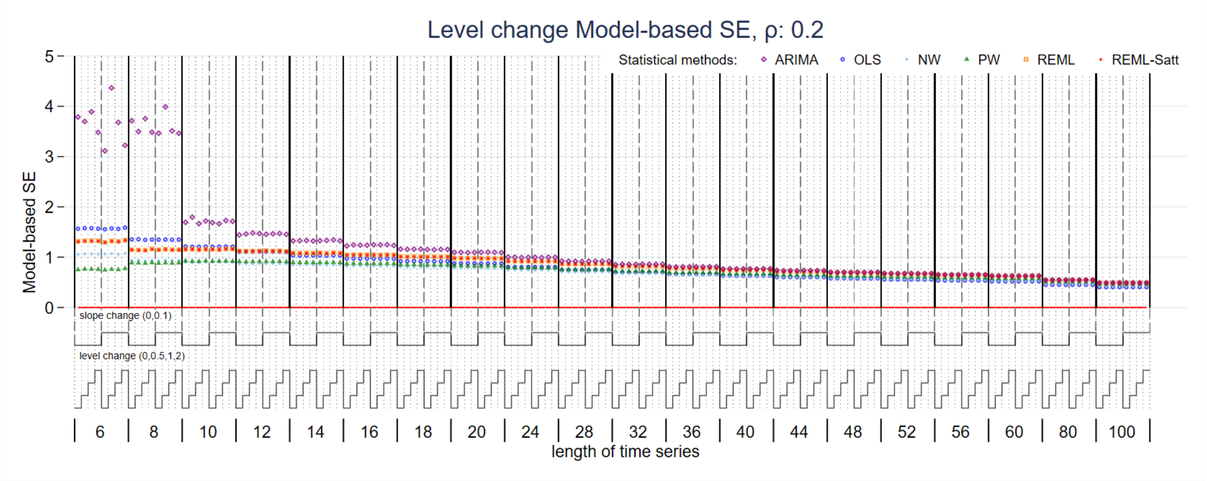


*Figure S24: Model-based standard error of level change estimate for magnitude of autocorrelation 0.2. Each data point shows the mean value from 10,000 simulations for a given combination of slope change, level change and length of time series. Abbreviations: ARIMA, autoregressive integrated moving average; OLS, ordinary least squares; NW, Newey-West; PW, Prais-Winsten; REML, restricted maximum likelihood; Satt, Satterthwaite.*

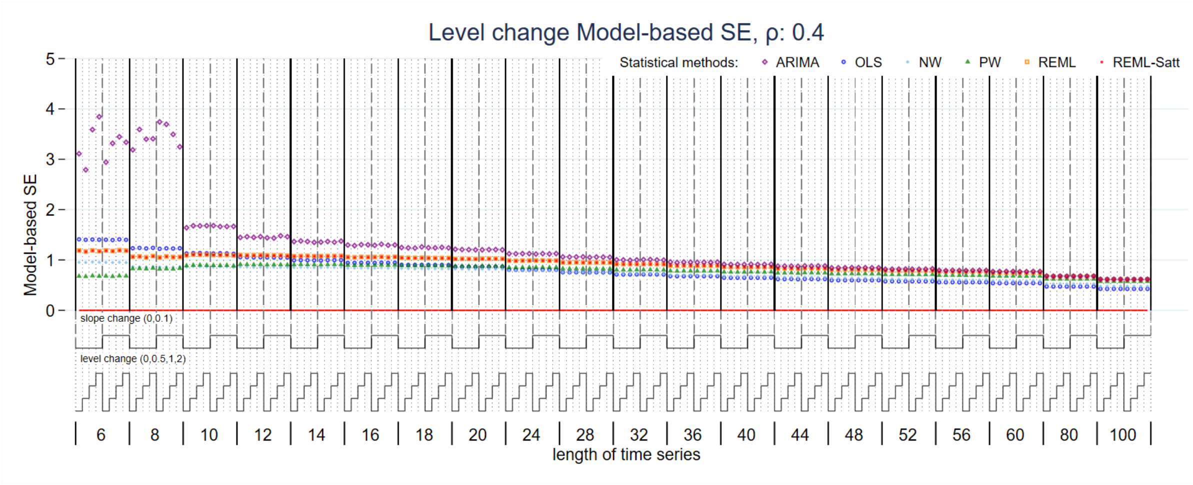


*Figure S25: Model-based standard error of level change estimate for magnitude of autocorrelation 0.4. Each data point shows the mean value from 10,000 simulations for a given combination of slope change, level change and length of time series. Abbreviations: ARIMA, autoregressive integrated moving average; OLS, ordinary least squares; NW, Newey-West; PW, Prais-Winsten; REML, restricted maximum likelihood; Satt, Satterthwaite.*

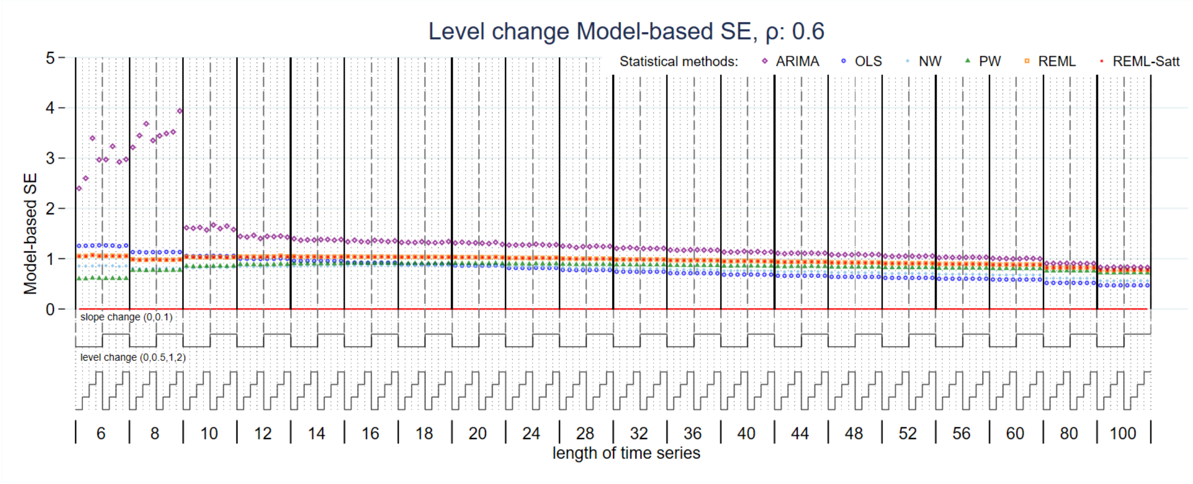


*Figure S26: Model-based standard error of level change estimate for magnitude of autocorrelation 0.6. Each data point shows the mean value from 10,000 simulations for a given combination of slope change, level change and length of time series. Abbreviations: ARIMA, autoregressive integrated moving average; OLS, ordinary least squares; NW, Newey-West; PW, Prais-Winsten; REML, restricted maximum likelihood; Satt, Satterthwaite.*

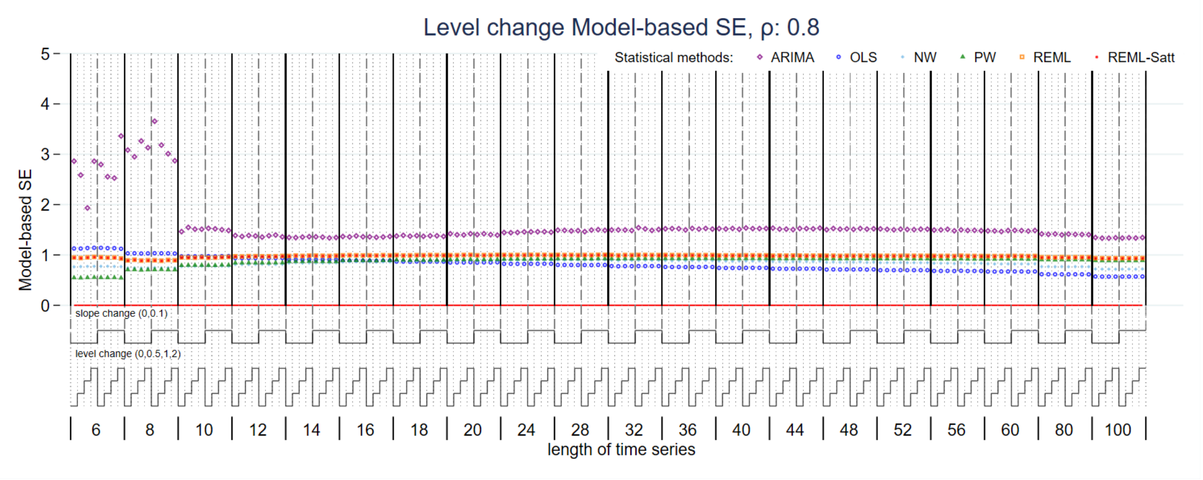


*Figure S27: Model-based standard error of level change estimate for magnitude of autocorrelation 0.8. Each data point shows the mean value from 10,000 simulations for a given combination of slope change, level change and length of time series. Abbreviations: ARIMA, autoregressive integrated moving average; OLS, ordinary least squares; NW, Newey-West; PW, Prais-Winsten; REML, restricted maximum likelihood; Satt, Satterthwaite.*

##### S 1.3.6 Slope change model-based standard error

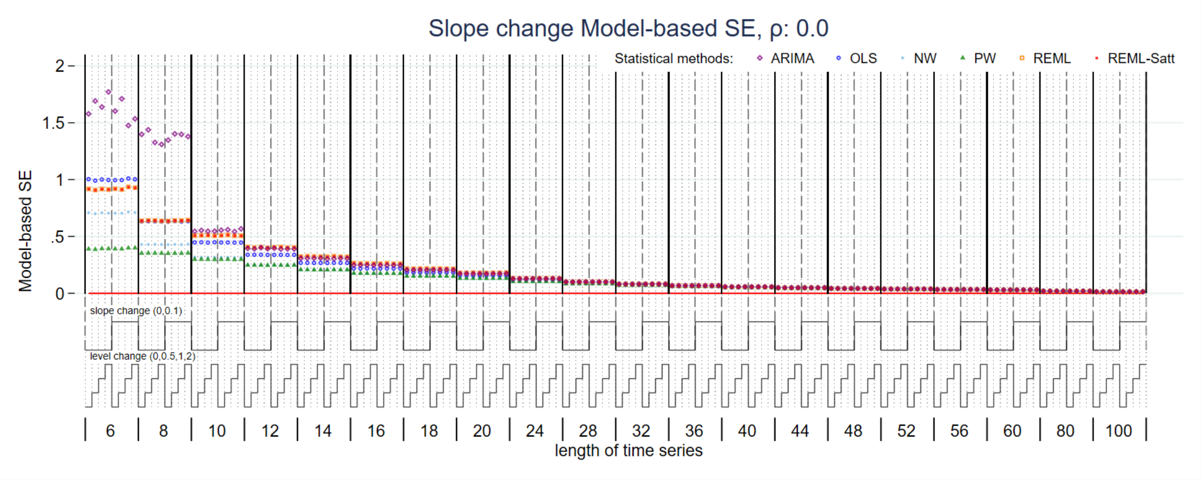


*Figure S28: Model-based standard error of slope change estimate for magnitude of autocorrelation 0. Each data point shows the mean value from 10,000 simulations for a given combination of slope change, level change and length of time series. Abbreviations: ARIMA, autoregressive integrated moving average; OLS, ordinary least squares; NW, Newey-West; PW, Prais-Winsten; REML, restricted maximum likelihood; Satt, Satterthwaite.*

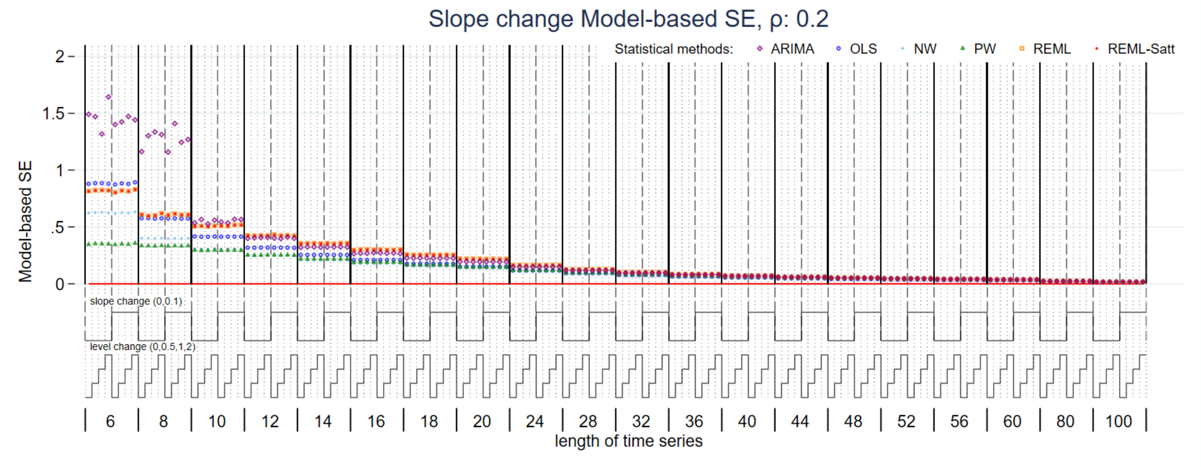


*Figure S29: Model-based standard error of slope change estimate for magnitude of autocorrelation 0.2. Each data point shows the mean value from 10,000 simulations for a given combination of slope change, level change and length of time series. Abbreviations: ARIMA, autoregressive integrated moving average; OLS, ordinary least squares; NW, Newey-West; PW, Prais-Winsten; REML, restricted maximum likelihood; Satt, Satterthwaite.*

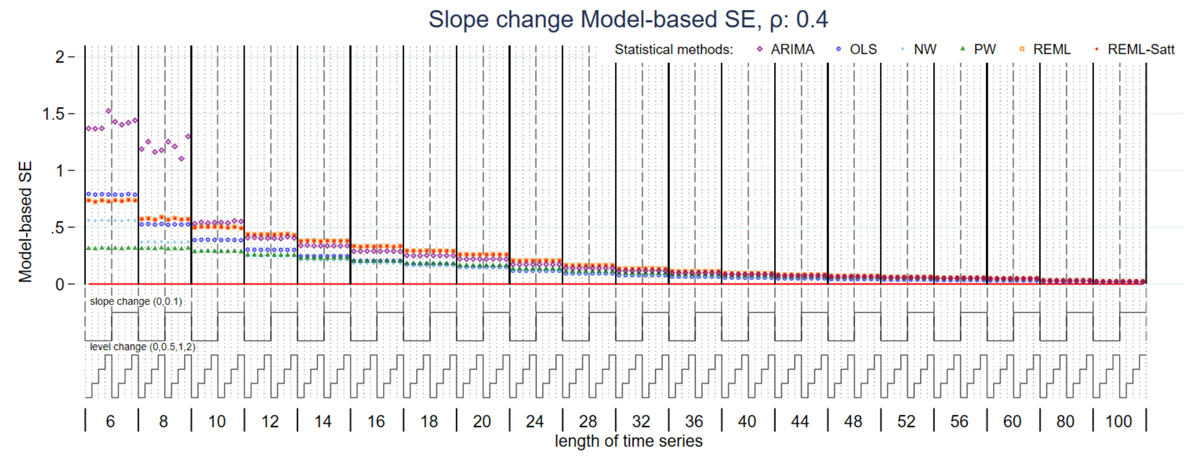


*Figure S30: Model-based standard error of slope change estimate for magnitude of autocorrelation 0.4. Each data point shows the mean value from 10,000 simulations for a given combination of slope change, level change and length of time series. Abbreviations: ARIMA, autoregressive integrated moving average; OLS, ordinary least squares; NW, Newey-West; PW, Prais-Winsten; REML, restricted maximum likelihood; Satt, Satterthwaite.*

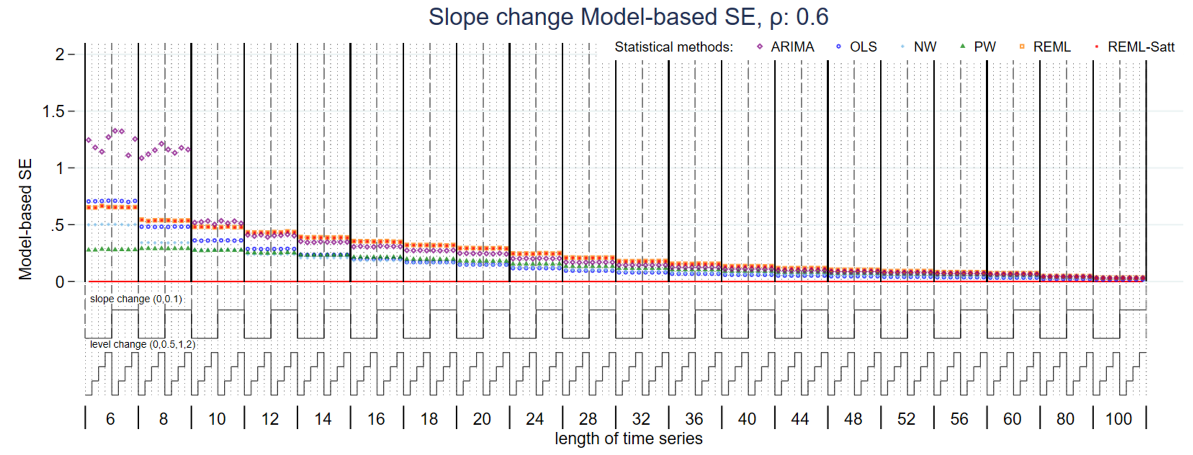


*Figure S31: Model-based standard error of slope change estimate for magnitude of autocorrelation 0.6. Each data point shows the mean value from 10,000 simulations for a given combination of slope change, level change and length of time series. Abbreviations: ARIMA, autoregressive integrated moving average; OLS, ordinary least squares; NW, Newey-West; PW, Prais-Winsten; REML, restricted maximum likelihood; Satt, Satterthwaite.*

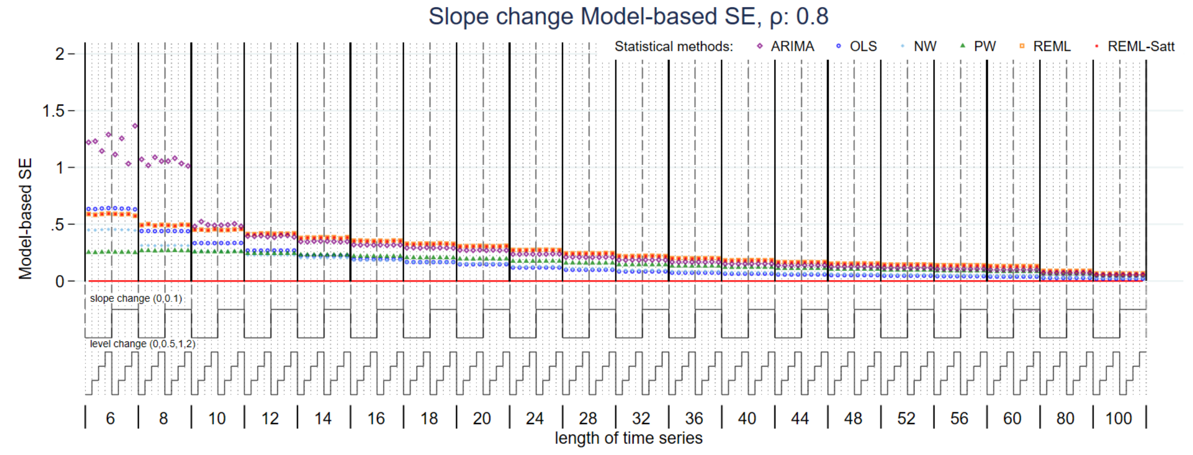


*Figure S32: Model-based standard error of slope change estimate for magnitude of autocorrelation 0.8. Each data point shows the mean value from 10,000 simulations for a given combination of slope change, level change and length of time series. Abbreviations: ARIMA, autoregressive integrated moving average; OLS, ordinary least squares; NW, Newey-West; PW, Prais-Winsten; REML, restricted maximum likelihood; Satt, Satterthwaite.*

#### S 1.3.7 Level change coverage

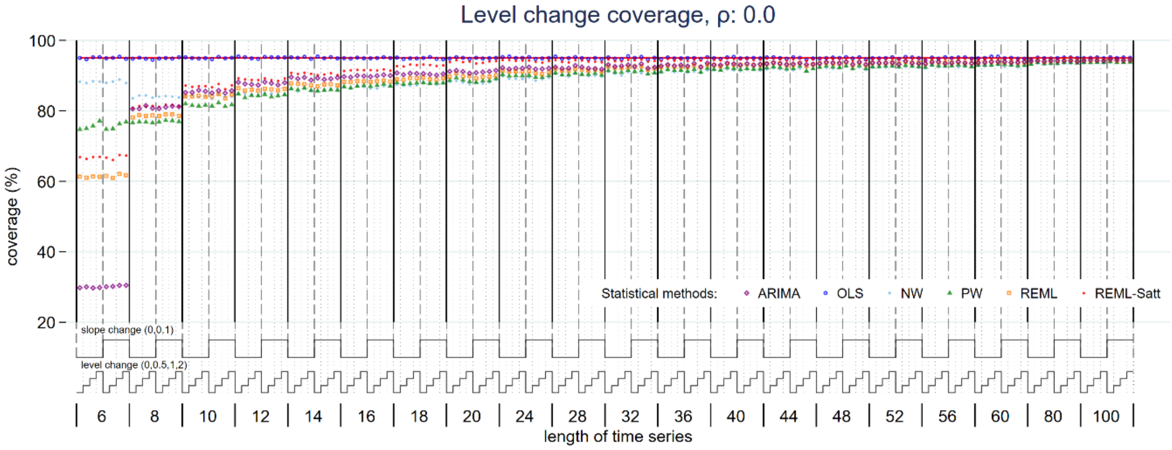


*Figure S33: Coverage of level change estimate for magnitude of autocorrelation 0. Each data point shows the mean value from 10,000 simulations for a given combination of slope change, level change and length of time series. Abbreviations: ARIMA, autoregressive integrated moving average; OLS, ordinary least squares; NW, Newey-West; PW, Prais-Winsten; REML, restricted maximum likelihood; Satt, Satterthwaite.*

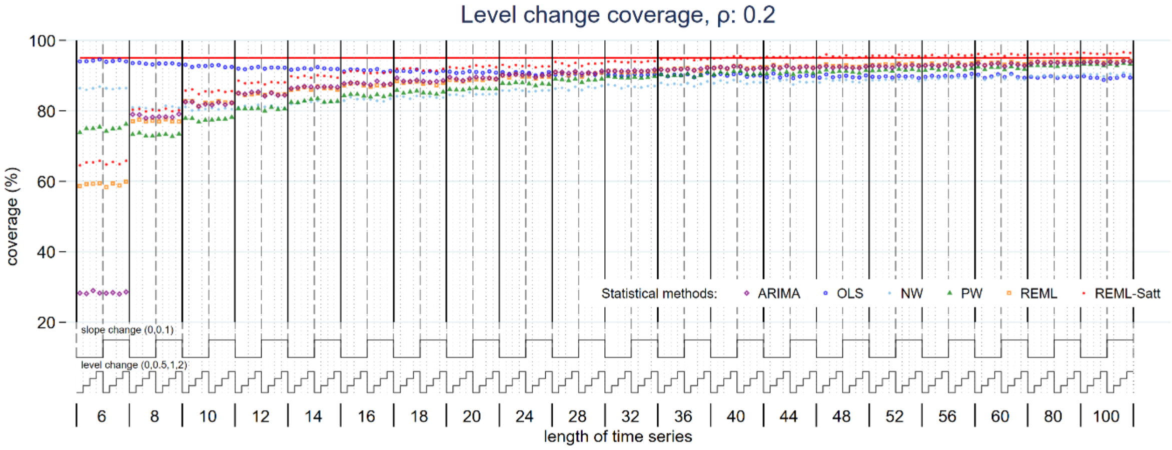


*Figure S34: Coverage of level change estimate for magnitude of autocorrelation 0.2. Each data point shows the mean value from 10,000 simulations for a given combination of slope change, level change and length of time series. Abbreviations: ARIMA, autoregressive integrated moving average; OLS, ordinary least squares; NW, Newey-West; PW, Prais-Winsten; REML, restricted maximum likelihood; Satt, Satterthwaite.*

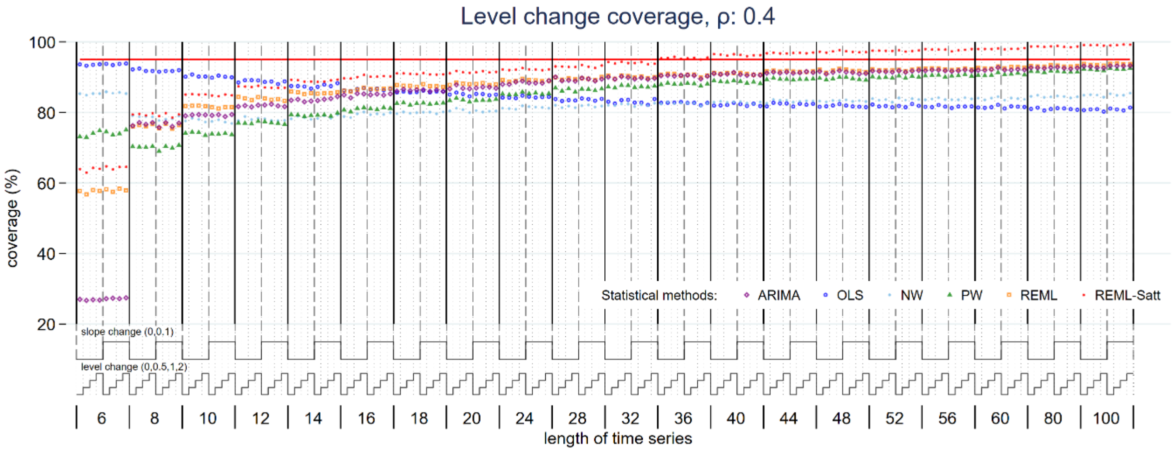


*Figure S35: Coverage of level change estimate for magnitude of autocorrelation 0.4. Each data point shows the mean value from 10,000 simulations for a given combination of slope change, level change and length of time series. Abbreviations: ARIMA, autoregressive integrated moving average; OLS, ordinary least squares; NW, Newey-West; PW, Prais-Winsten; REML, restricted maximum likelihood; Satt, Satterthwaite.*

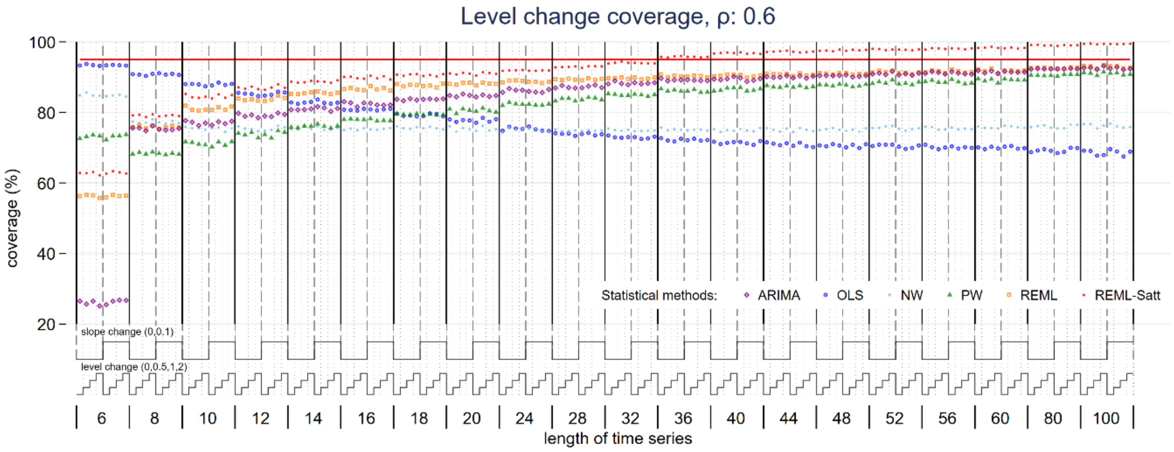


*Figure S36: Coverage of level change estimate for magnitude of autocorrelation 0.6. Each data point shows the mean value from 10,000 simulations for a given combination of slope change, level change and length of time series. Abbreviations: ARIMA, autoregressive integrated moving average; OLS, ordinary least squares; NW, Newey-West; PW, Prais-Winsten; REML, restricted maximum likelihood; Satt, Satterthwaite.*

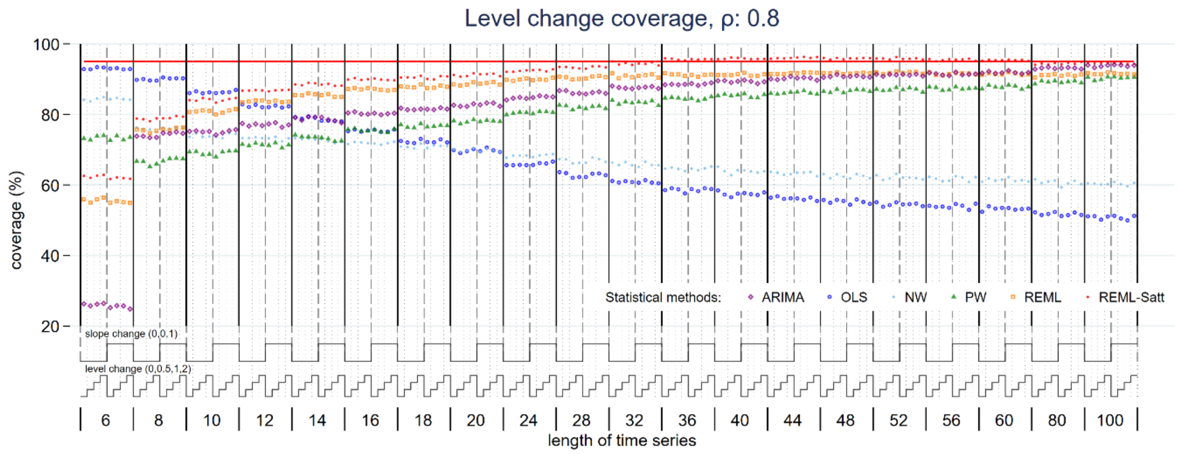


*Figure S37: Coverage of level change estimate for magnitude of autocorrelation 0.8. Each data point shows the mean value from 10,000 simulations for a given combination of slope change, level change and length of time series. Abbreviations: ARIMA, autoregressive integrated moving average; OLS, ordinary least squares; NW, Newey-West; PW, Prais-Winsten; REML, restricted maximum likelihood; Satt, Satterthwaite.*

#### S 1.3.8 Slope change coverage

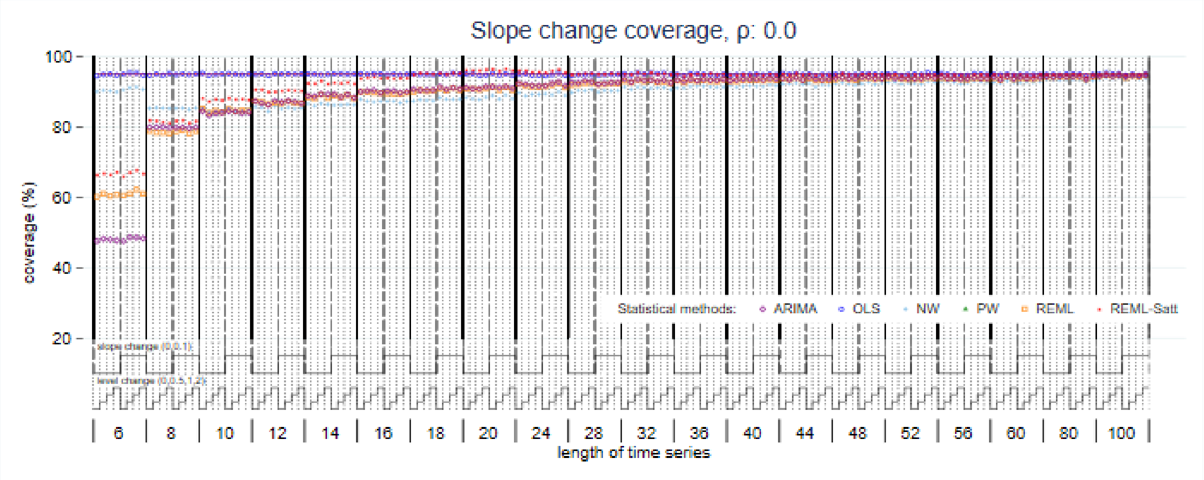


*Figure S38: Coverage of slope change estimate for magnitude of autocorrelation 0. Each data point shows the mean value from 10,000 simulations for a given combination of slope change, level change and length of time series. Abbreviations: ARIMA, autoregressive integrated moving average; OLS, ordinary least squares; NW, Newey-West; PW, Prais-Winsten; REML, restricted maximum likelihood; Satt, Satterthwaite.*

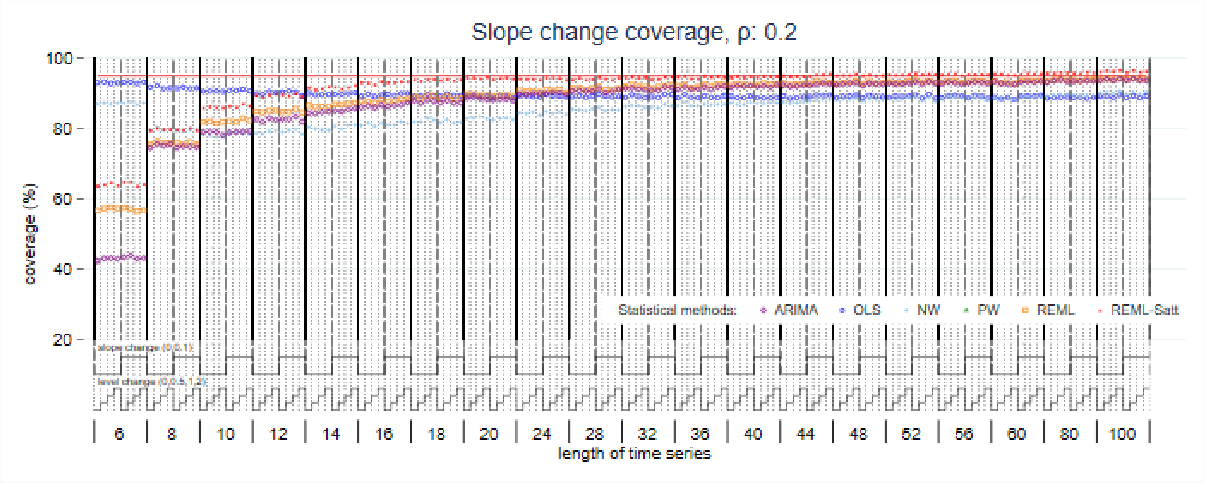


*Figure S39: Coverage of slope change estimate for magnitude of autocorrelation 0.2. Each data point shows the mean value from 10,000 simulations for a given combination of slope change, level change and length of time series. Abbreviations: ARIMA, autoregressive integrated moving average; OLS, ordinary least squares; NW, Newey-West; PW, Prais-Winsten; REML, restricted maximum likelihood; Satt, Satterthwaite.*

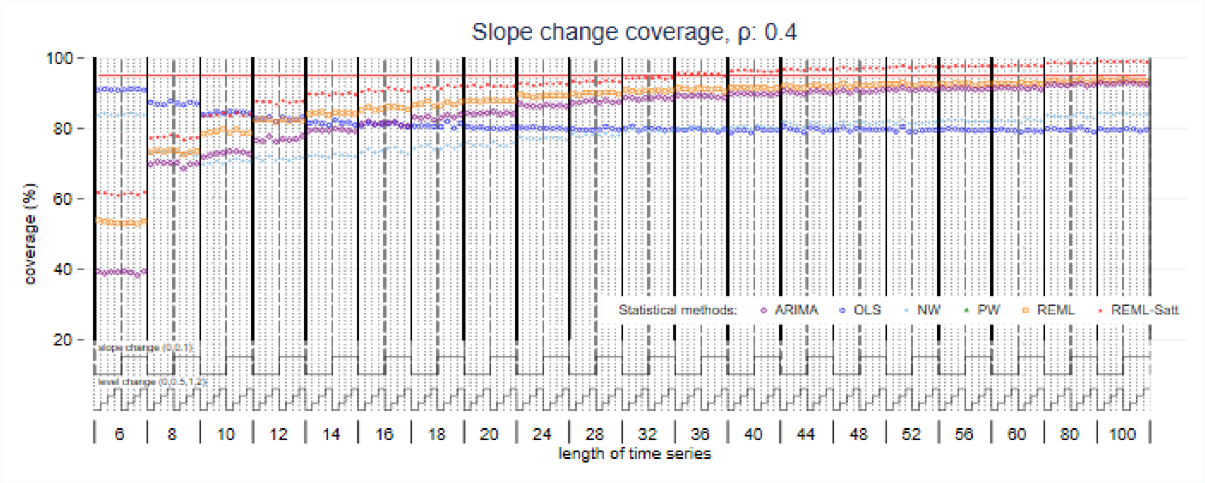


*Figure S40: Coverage of slope change estimate for magnitude of autocorrelation 0.4. Each data point shows the mean value from 10,000 simulations for a given combination of slope change, level change and length of time series. Abbreviations: ARIMA, autoregressive integrated moving average; OLS, ordinary least squares; NW, Newey-West; PW, Prais-Winsten; REML, restricted maximum likelihood; Satt, Satterthwaite.*

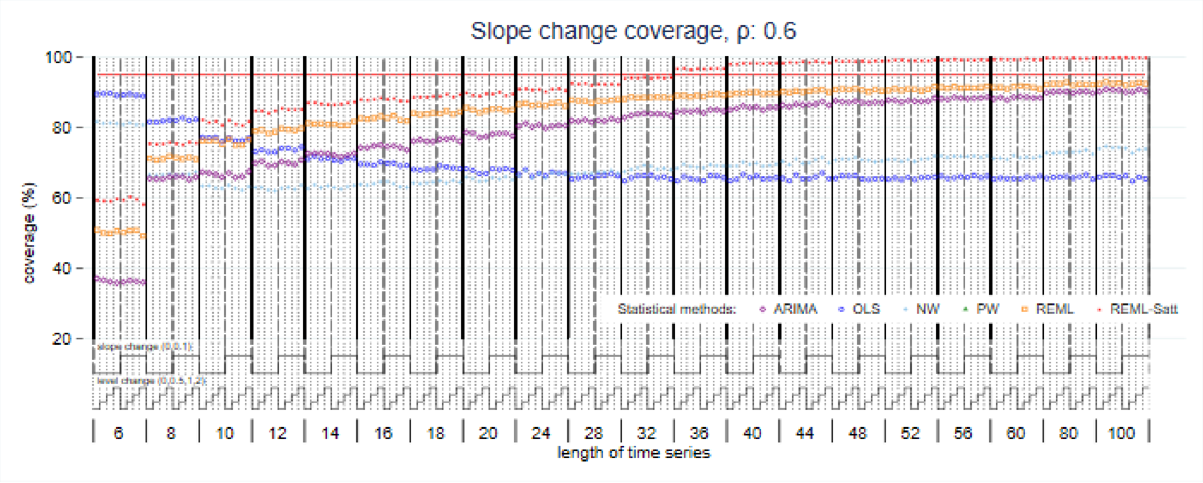


*Figure S41: Coverage of slope change estimate for magnitude of autocorrelation 0.6. Each data point shows the mean value from 10,000 simulations for a given combination of slope change, level change and length of time series. Abbreviations: ARIMA, autoregressive integrated moving average; OLS, ordinary least squares; NW, Newey-West; PW, Prais-Winsten; REML, restricted maximum likelihood; Satt, Satterthwaite.*

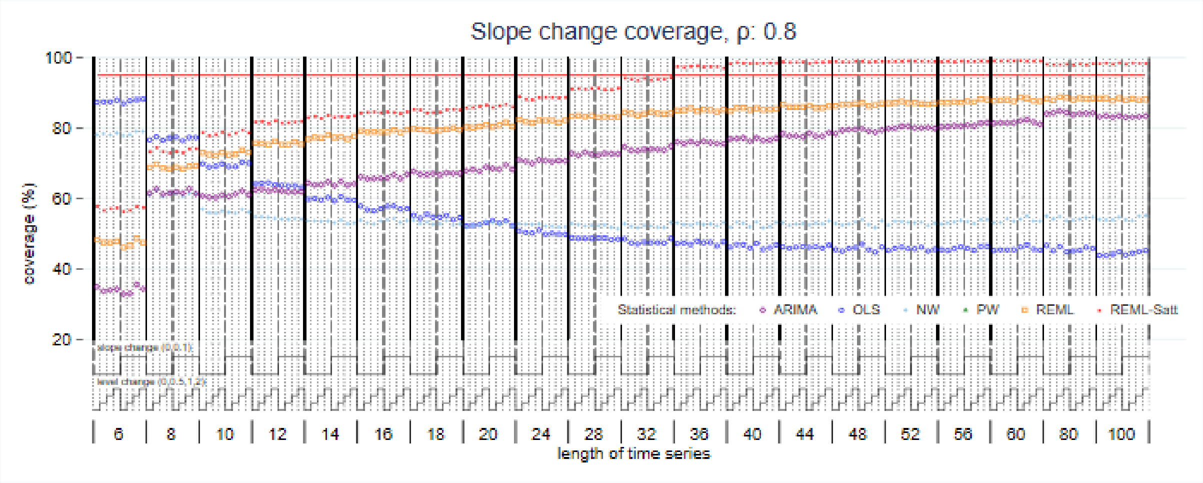


*Figure S42: Coverage of slope change estimate for magnitude of autocorrelation 0.8. Each data point shows the mean value from 10,000 simulations for a given combination of slope change, level change and length of time series. Abbreviations: ARIMA, autoregressive integrated moving average; OLS, ordinary least squares; NW, Newey-West; PW, Prais-Winsten; REML, restricted maximum likelihood; Satt, Satterthwaite.*

#### S 1.3.9 Estimate of autocorrelation

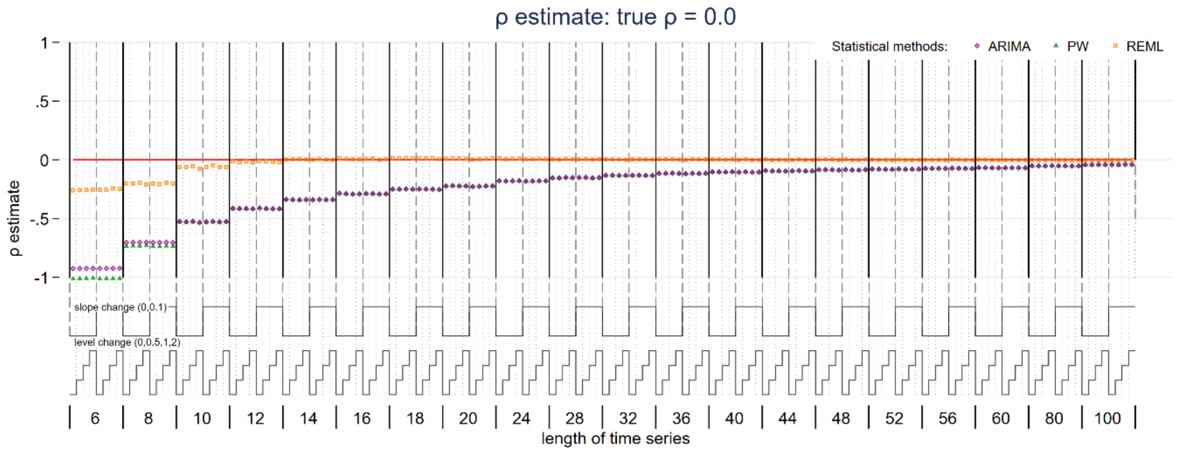


*Figure S43: Autocorrelation estimate for true magnitude of autocorrelation 0. Each data point shows the mean value from 10,000 simulations for a given combination of slope change, level change and length of time series. Abbreviations: ARIMA, autoregressive integrated moving average; PW, Prais-Winsten; REML, restricted maximum likelihood.*

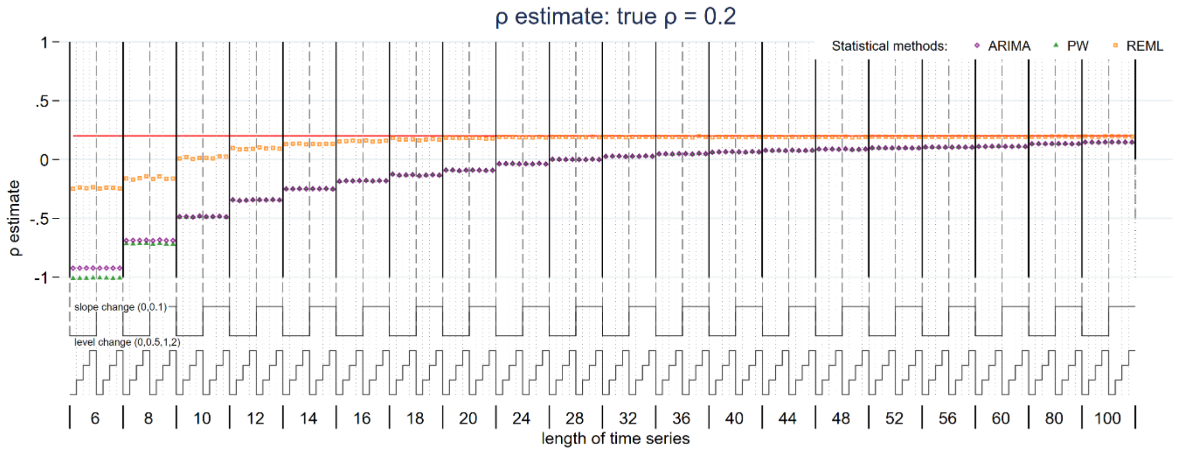


*Figure S44: Autocorrelation estimate for true magnitude of autocorrelation 0.2. Each data point shows the mean value from 10,000 simulations for a given combination of slope change, level change and length of time series. Abbreviations: ARIMA, autoregressive integrated moving average; PW, Prais-Winsten; REML, restricted maximum likelihood.*

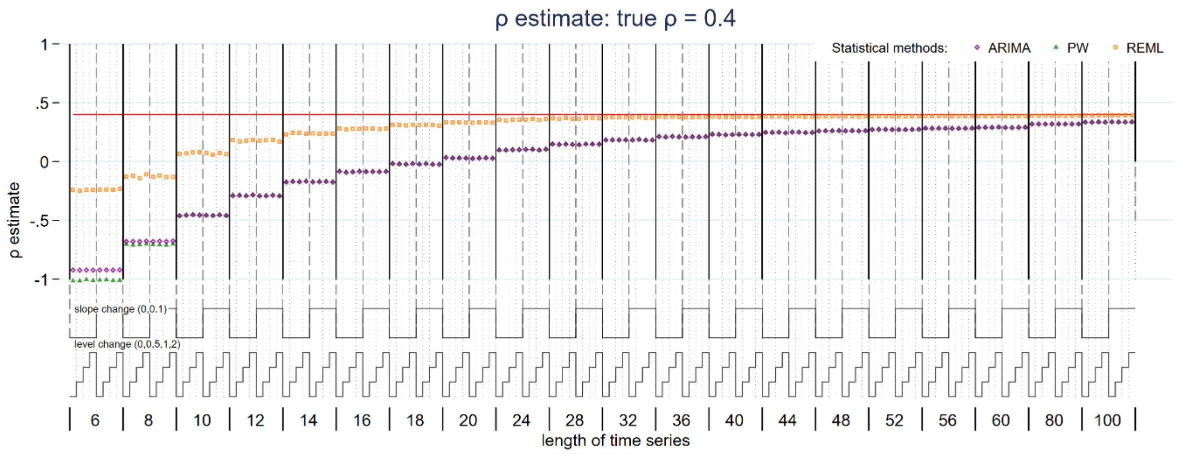


*Figure S45: Autocorrelation estimate for true magnitude of autocorrelation 0.4. Each data point shows the mean value from 10,000 simulations for a given combination of slope change, level change and length of time series. Abbreviations: ARIMA, autoregressive integrated moving average; PW, Prais-Winsten; REML, restricted maximum likelihood.*

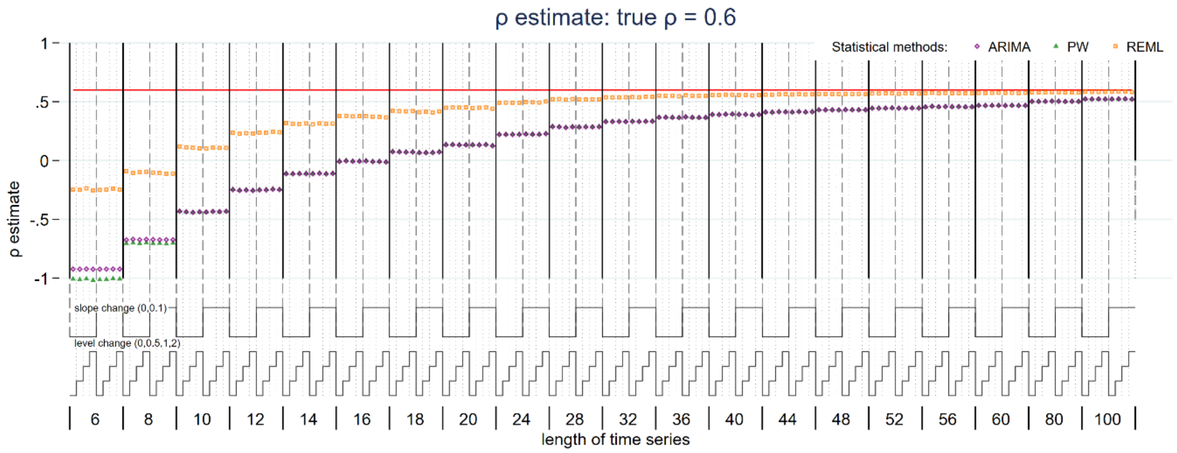


*Figure S46: Autocorrelation estimate for true magnitude of autocorrelation 0.6. Each data point shows the mean value from 10,000 simulations for a given combination of slope change, level change and length of time series. Abbreviations: ARIMA, autoregressive integrated moving average; PW, Prais-Winsten; REML, restricted maximum likelihood.*

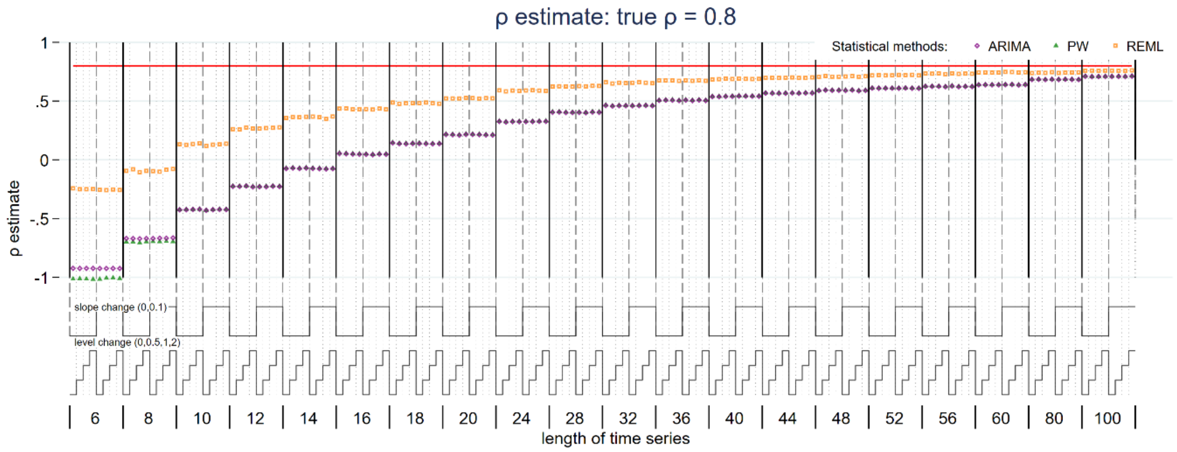


*Figure S47: Autocorrelation estimate for true magnitude of autocorrelation 0.8. Each data point shows the mean value from 10,000 simulations for a given combination of slope change, level change and length of time series. Abbreviations: ARIMA, autoregressive integrated moving average; PW, Prais-Winsten; REML, restricted maximum likelihood.*

#### Supplementary 1.4 Standard error of level change parameter for OLS

Additional simulations were undertaken to examine the behaviour of the OLS standard error estimator for the level change for additional autocorrelations (0.7 and 0.9). This showed a similar pattern of initially increasing SEs with an increasing number of points, which we had observed in the main simulation study for an underlying autocorrelation of 0.8.

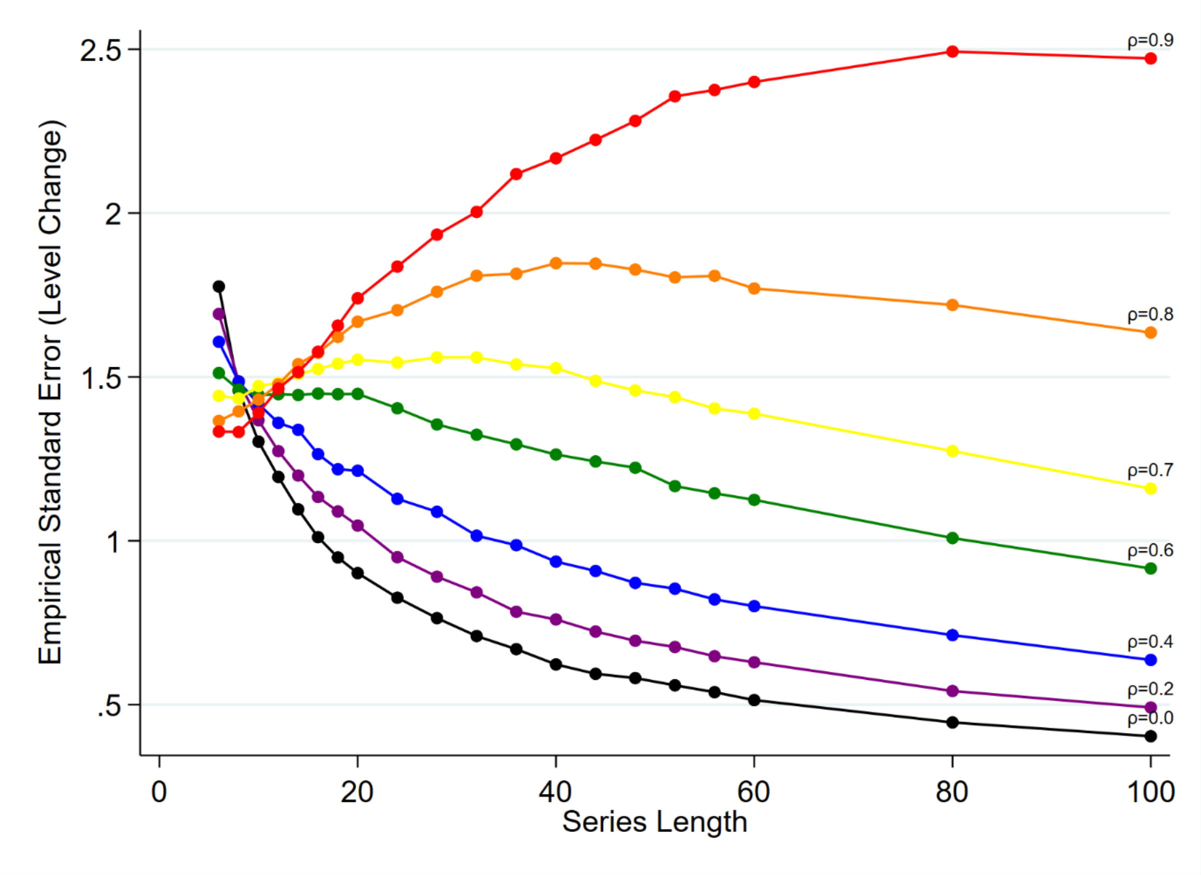


*Figure S48: Empirical standard error (SE) of level change parameter for ordinary least squares regression. The horizontal axis shows the length of the time series, the vertical axis shows the empirical SE. Each coloured line shows the results for a different magnitude of autocorrelation ranging from 0 (bottom right, black) to 0.8 (top right, red). The simulation combination presented represents a level change of 2 and slope change of 0.1; however, other combinations give similar results.*

#### Supplementary 1.5 Model-based standard errors

Generally, similar patterns were observed for the model-based SEs (of level and slope change) as compared with the empirical SEs, across all model combinations as would be expected (Appendix 1.5 for level change, Appendix 1.6 for slope change). The exception was for the OLS SEs for the level change in which the empirical SE showed divergent behaviour for higher levels of autocorrelation compared to the other statistical methods but not for the model-based SE, where it behaved similarly to the other statistical methods.

The model-based SEs of the level change differed by the statistical method, with those estimated by ARIMA being the largest compared to all other methods (Figure S47). The model-based errors for slope change estimated by ARIMA were substantially larger than the other methods for short series (< 10 data points) (Figure 48).

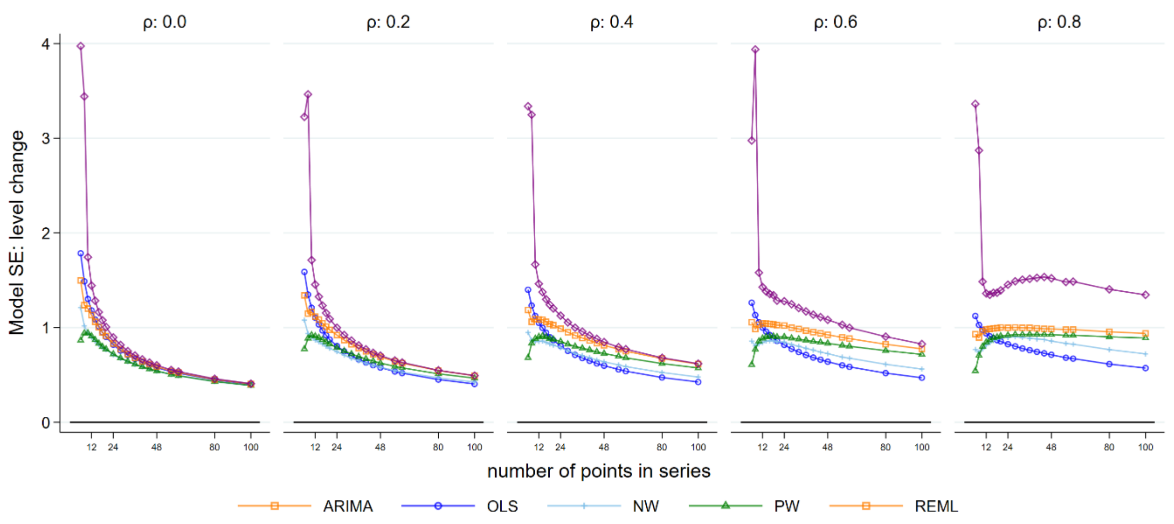


*Figure 49: Model-based standard errors (SE) for the level change parameter. The horizontal axis shows the length of the time series, the vertical axis shows the model-based SE. The five vertical columns display the results for different values of autocorrelation. The simulation combination presented represent a level change of 2 and slope change of 0.1; however, other combinations give similar results. Abbreviations: ARIMA, autoregressive integrated moving average; OLS, ordinary least squares; NW, Newey-West; PW, Prais-Winsten; REML, restricted maximum likelihood.*

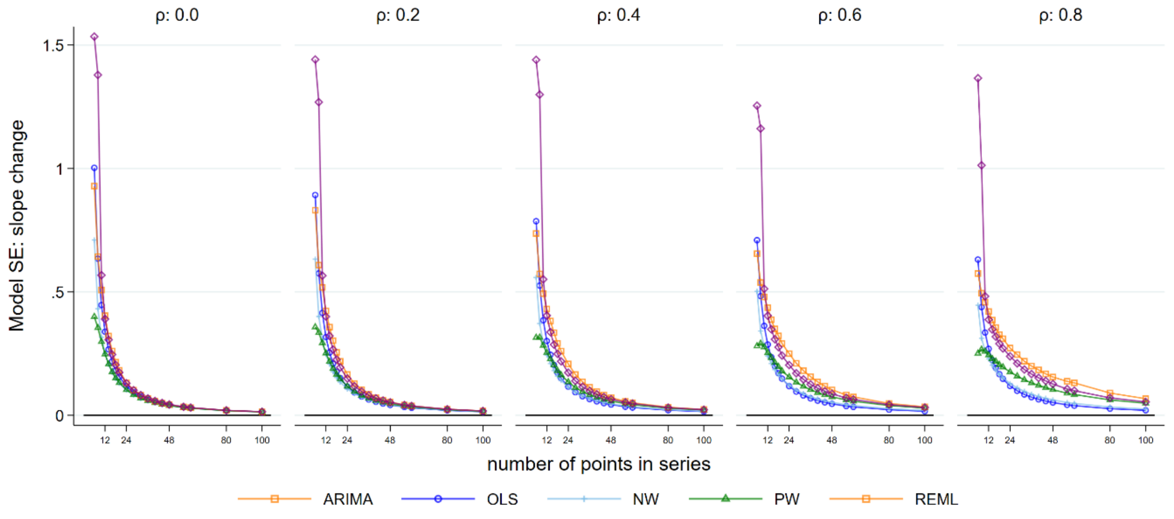


*Figure 50: Model-based standard errors (SE) for the level change parameter. The horizontal axis shows the length of the time series, the vertical axis shows the model-based SE. The five vertical columns display the results for different values of autocorrelation The simulation combination presented represent a level change of 2 and slope change of 0.1; however, other combinations give similar results. Abbreviations: ARIMA, autoregressive integrated moving average; OLS, ordinary least squares; NW, Newey-West; PW, Prais-Winsten; REML, restricted maximum likelihood.*

#### Supplementary 1.6 Comparison between empirical and model-based standard errors: slope change

The ratio of model to empirical-based standard errors are shown in Figure S51. The accompanying text is in section 5.2.2.

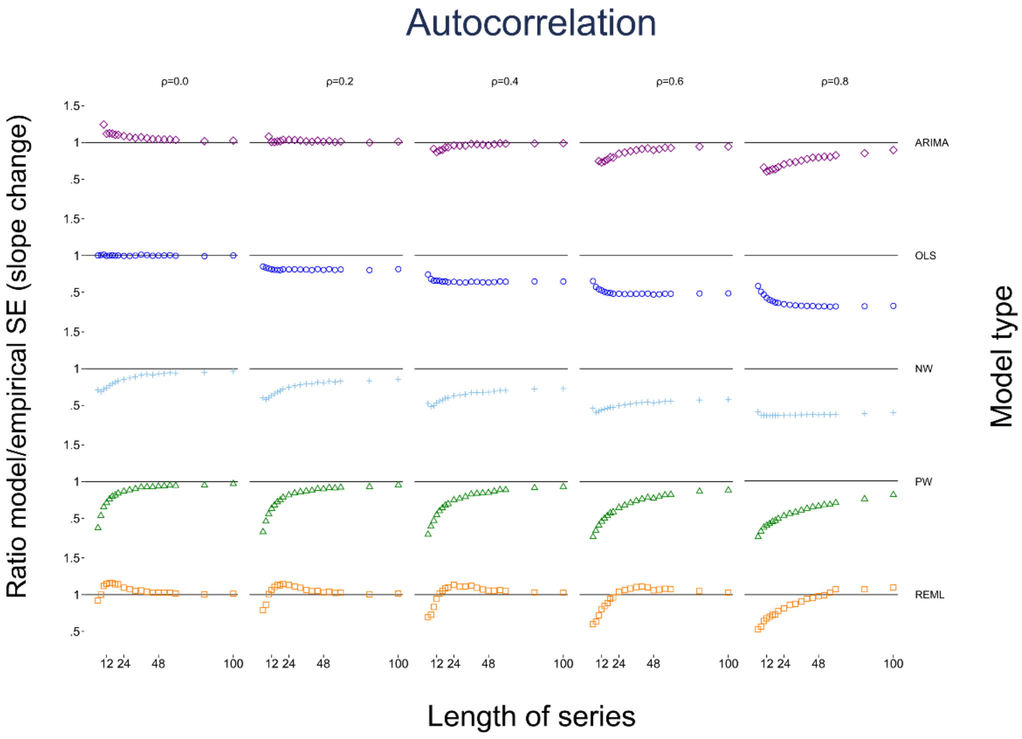


*Figure S51: Scatter plots of the ratio of model-based standard error (SE) to the empirical SE for the slope change parameter with different levels of autocorrelation and series length. The horizontal axis represents the number of points in the time series, the vertical axis shows the ratio of model-based to empirical SE. The five vertical columns display the results for different values of autocorrelation. The simulation combination presented is for a level change of 2 and slope change of 0.1; however, other combinations give similar results. The first two series lengths are not shown for the ARIMA method due to extreme values. The Satterthwaite adjustment to the REML does not impact the estimate of SE, hence details of this method are not shown. Abbreviations: ARIMA, autoregressive integrated moving average; OLS, ordinary least squares; NW, Newey-West; PW, Prais-Winsten; REML, restricted maximum likelihood.*

#### Supplementary 1.7 Confidence interval coverage: slope change

Figure S52 shows the 95% coverage of the slope change estimates. Accompanying text is in Section 5.3.

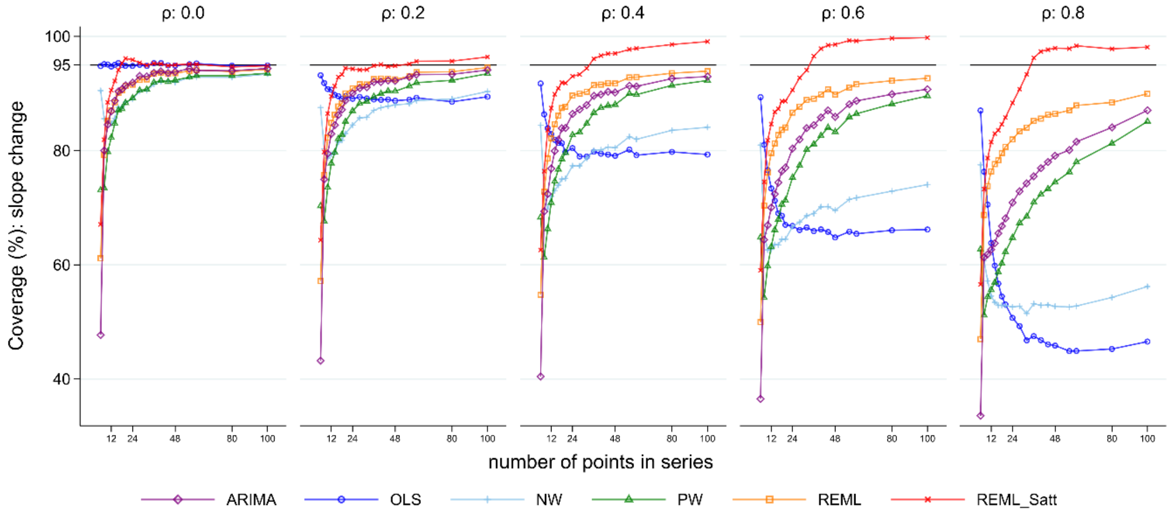


*Figure S52: Coverage for the slope change parameter. Each point is the proportion of the 10,000 simulations in which the 95% confidence interval included the true value of the parameter. The solid black line depicts the nominal 95% coverage level. The simulation combination presented is for a level change of 2 and slope change of 0.1; however, other combinations give similar results. Abbreviations: ARIMA, autoregressive integrated moving average; OLS, ordinary least squares; NW, Newey-West; PW, Prais-Winsten; REML, restricted maximum likelihood; Satt, Satterthwaite.*

### Supplementary 1.8 Power

The following graphs show the estimated power for additional level and slope change parameters. The results presented below should be viewed as approximate power only and will generally be lower than the value observed if coverage was at least 95%.

#### S 1.8.1 Level change

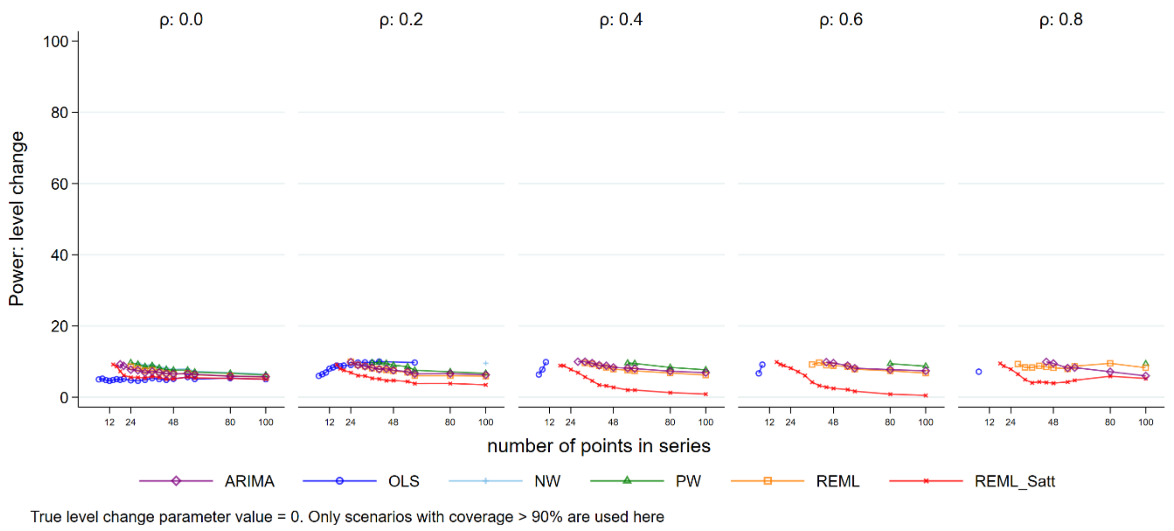


*Figure S53: Power for level change, true value 0. Each point is the mean number of times the 95% confidence interval of the estimate did not include zero from 10,000 simulations. Abbreviations: ARIMA, autoregressive integrated moving average; OLS, ordinary least squares; NW, Newey-West; PW, Prais-Winsten; REML, restricted maximum likelihood; Satt, Satterthwaite; NW, Newey-West.*

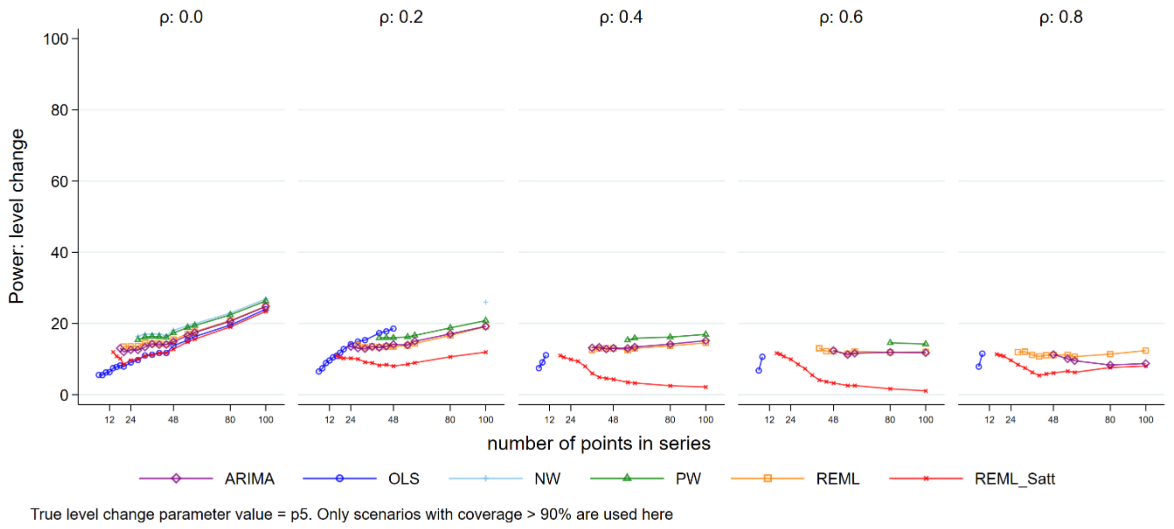


*Figure S54: Power for level change, true value .5. Each point is the mean number of times the 95% confidence interval of the estimate did not include zero from 10,000 simulations. Abbreviations: ARIMA, autoregressive integrated moving average; OLS, ordinary least squares; NW, Newey-West; PW, Prais-Winsten; REML, restricted maximum likelihood; Satt, Satterthwaite; NW, Newey-West.*

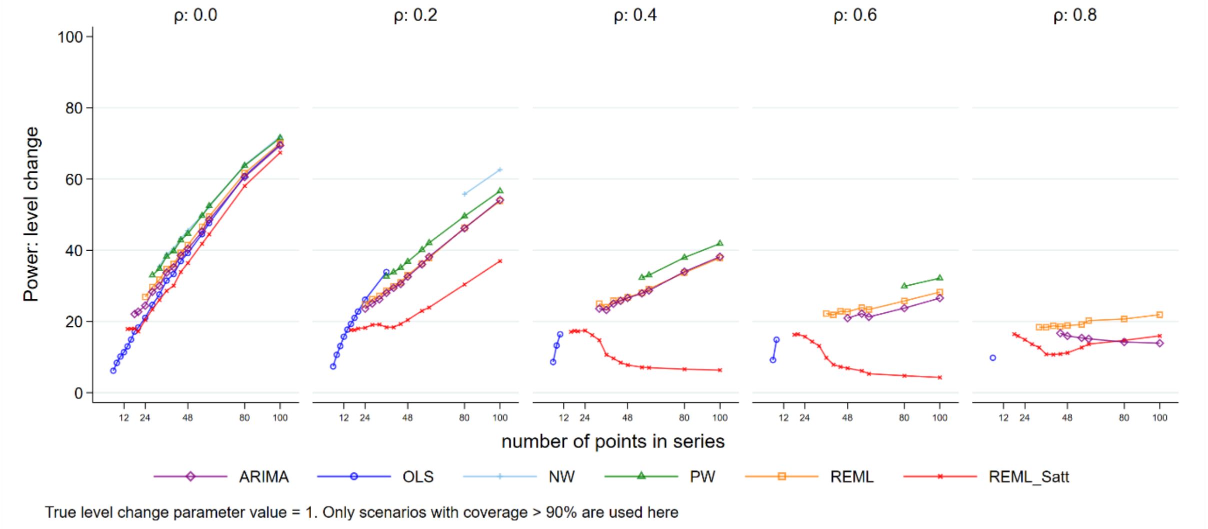


*Figure S55: Power for level change, true value 1. Each point is the mean number of times the 95% confidence interval of the estimate did not include zero from 10,000 simulations. Abbreviations: ARIMA, autoregressive integrated moving average; OLS, ordinary least squares; NW, Newey-West; PW, Prais-Winsten; REML, restricted maximum likelihood; Satt, Satterthwaite; NW, Newey-West.*

#### S 1.8.2 Slope change

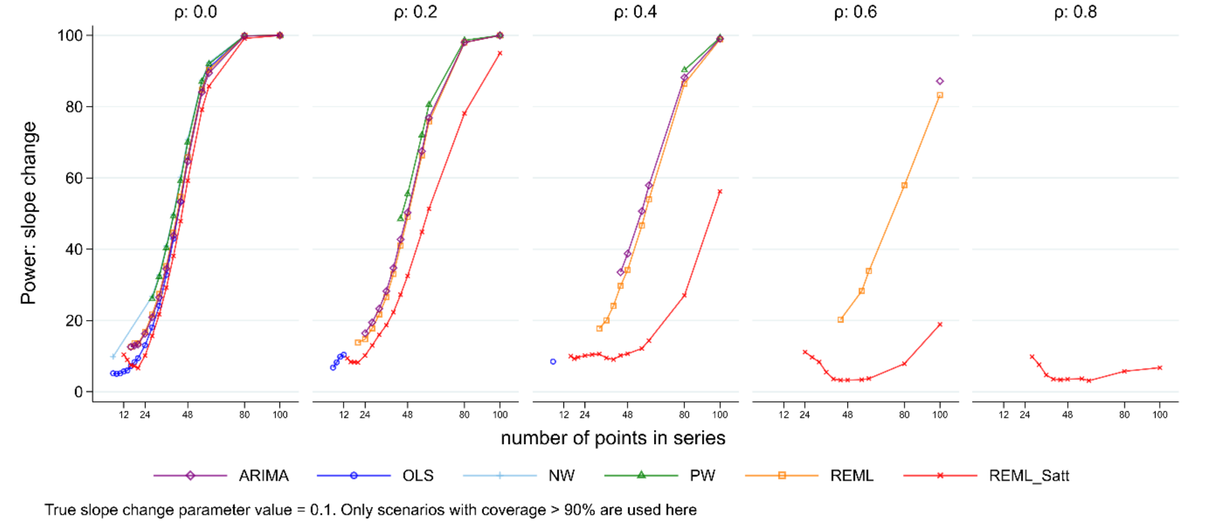


*Figure S56: Power for slope change, true value 0.1. Each point is the mean number of times the 95% confidence interval of the estimate did not include zero from 10,000 simulations. Abbreviations: ARIMA, autoregressive integrated moving average; OLS, ordinary least squares; NW, Newey-West; PW, Prais-Winsten; REML, restricted maximum likelihood; Satt, Satterthwaite; NW, Newey-West.*

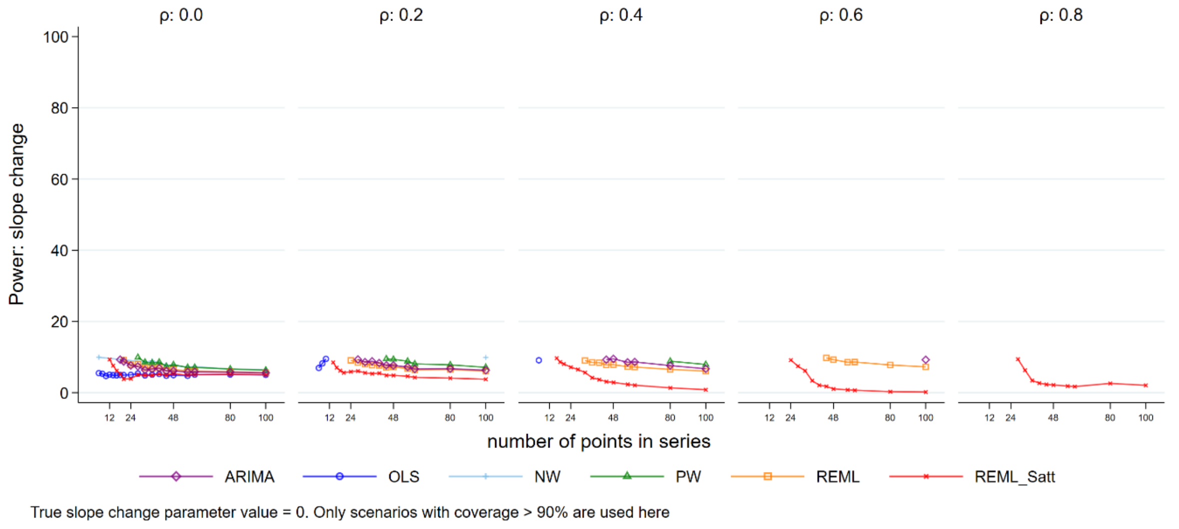


*Figure S57: Power for slope change, true value 0. Each point is the mean number of times the 95% confidence interval of the estimate did not include zero from 10,000 simulations. Abbreviations: ARIMA, autoregressive integrated moving average; OLS, ordinary least squares; NW, Newey-West; PW, Prais-Winsten; REML, restricted maximum likelihood; Satt, Satterthwaite; NW, Newey-West.*

#### Supplementary 1.9 Standard error of autocorrelation coefficient estimates.

The following graph shows the empirical SE of the estimates of the magnitude of autocorrelation.

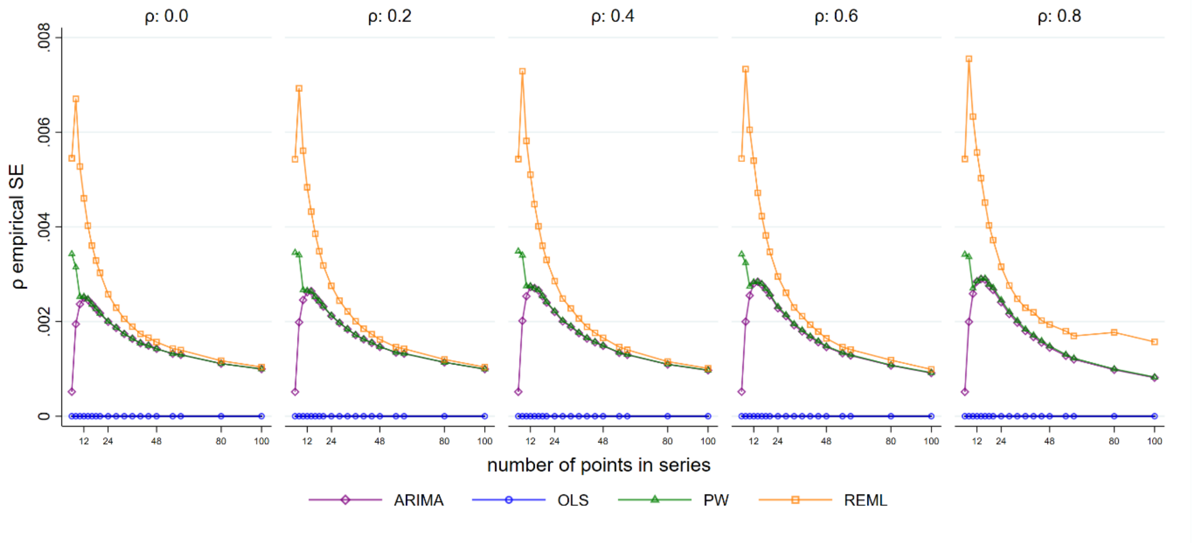


*Figure S58: Empirical standard error (SE) for autocorrelation coefficient estimates. The horizontal axis shows the length of the time series. The vertical axis shows the empirical SE of the autocorrelation coefficient estimates. The five plots display the results for different values of autocorrelation ranging from 0 to 0.8. Each coloured point shows the mean value of the SE of the autocorrelation coefficient estimates from 10,000 simulations for a given combination of autocorrelation coefficient and number of points in the data series The simulation combinations presented represent a model structure with a level change of 2 and slope change of 0.1; however, other combinations give similar results. Abbreviations: ARIMA, autoregressive integrated moving average; OLS, ordinary least squares; PW, Prais-Winsten; REML, restricted maximum likelihood.*

#### Supplementary 1.10 Convergence of estimation methods

Figure S54 shows the number of times the estimation methods converged. Accompanying text is in Section 5.6

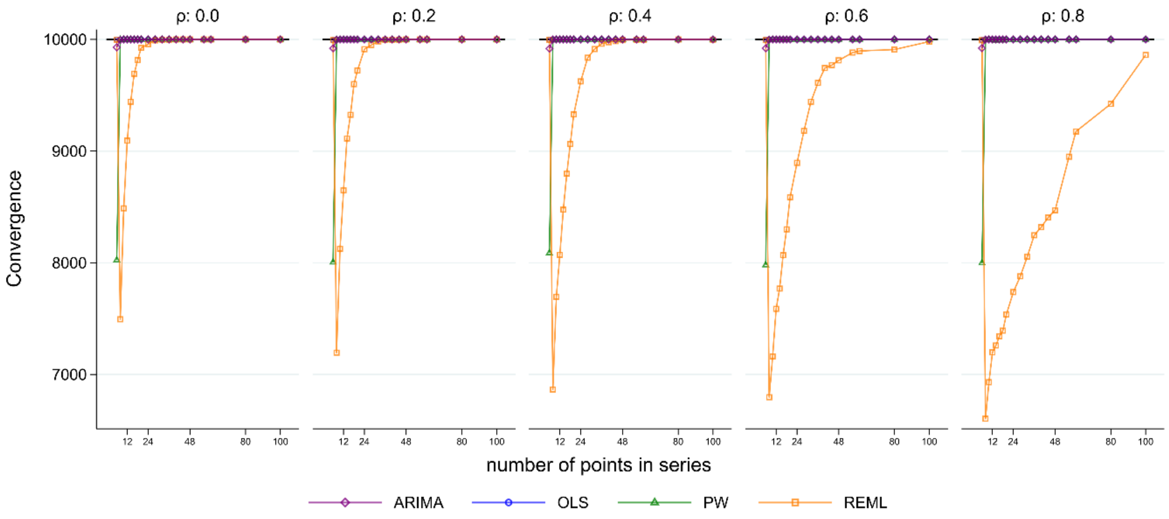


*Figure S59: Model convergence. The horizontal axis shows the length of the time series. The vertical axis shows the number of times each method converged out of 10,000 simulation replications. The five plots display the results for different values of autocorrelation ranging from 0 to 0.8. The simulation combination presented is for a level change of 2 and slope change of 0.1; however, other combinations give similar results. Abbreviations: ARIMA, autoregressive integrated moving average; OLS, ordinary least squares; PW, Prais-Winsten; REML, restricted maximum likelihood.*

#### Supplementary 1.11 Coverage by autocorrelation bias: slope change

Figure S60 shows the coverage for slope change versus the bias in autocorrelation estimate. Accompanying text is in section 7.1.

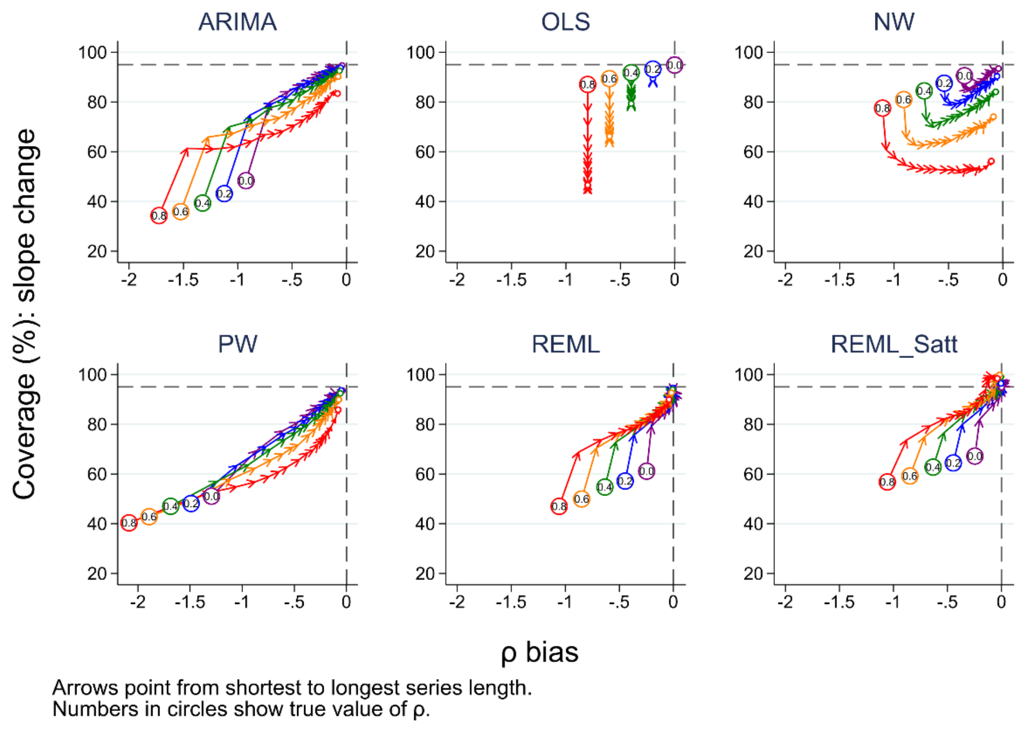


*Figure S60: Bias in autocorrelation estimate versus coverage for slope change. The horizontal axis shows the bias in the autocorrelation estimate. The vertical axis shows the percentage coverage. The horizontal dashed line indicates 95% coverage, the vertical dashed line indicates no bias in the estimate of autocorrelation. Each colour represents a different value of underlying autocorrelation, ranging from zero (purple) to 0.8 (red), with each value displayed in a circle at the smallest series length (six points). The arrows point from shortest to longest series length, with the small circles at the end of each line showing coverage at a series length of 100 data points. Each data point shows the mean value from 10,000 simulations for a given combination of autocorrelation coefficient and number of points in the series. The simulation combination presented is for a level change of 2 and slope change of 0.1; however, other combinations give similar results. Abbreviations: ARIMA, autoregressive integrated moving average; OLS, ordinary least squares; NW, Newey-West; PW, Prais-Winsten; REML, restricted maximum likelihood; Satt, Satterthwaite.*
